# Sleep and circadian health in the UK Biobank: Report on the 2023 sleep questionnaire enhancement

**DOI:** 10.1101/2025.09.24.25336551

**Authors:** Katrina Y. K. Tse, Hang Yuan, Charilaos Zisou, Jo Holliday, Colin A. Espie, Derk-Jan Dijk, Angus Burns, Aiden Doherty, Jacqueline Lane, Hanna Ollila, Allan Pack, David Ray, Susan Redline, Rebecca Richmond, Richa Saxena, Eva S. Schernhammer, Barbara Schormair, Kai Spiegelhalder, Heming Wang, Julianne Winkelmann, Andrew R. Wood, Martin K. Rutter, Emmanuel Mignot, Simon D. Kyle

## Abstract

**Study Objectives:** Our study introduced the 2023 UK Biobank sleep questionnaire and described variation in sleep health dimensions and prevalence of disordered sleep.

**Methods:** A questionnaire comprising validated measures and bespoke items was developed to capture key self-reported domains of sleep health and symptoms of sleep disorders. We quantified cohort prevalence of operationally defined sleep disorders and assessed patterning of sleep health dimensions across key sociodemographic and clinically relevant variables.

**Results:** 327,752 individuals were invited of whom 185,056 (56.5%) completed at least one module and were included in the analysis. Respondents were predominately from a White ethnic background (96.8%), had a mean age of 69.9 (SD, 7.5) years, 57.9% were female, and 25.5% were in employment. Compared to non-respondents, respondents were more likely to be female, tended to be better educated, healthier, and exhibit lower levels of socioeconomic deprivation, although baseline sleep variables were similar between respondents and non-respondents. Around 40% of respondents reported sleep duration less than 7 hours and 49% reported poor sleep quality (Pittsburgh Sleep Quality Index > 5). Approximately one-quarter (25.2%) met criteria for at least one operationally defined sleep disorder, with insomnia being the most common (14.4%) followed by obstructive sleep apnoea (8.0%), restless legs syndrome (4.1%), and frequent nightmares (3.7%). Sleep disorders were associated with higher levels of anxiety, depression, fatigue, and cognitive complaints.

**Conclusions:** Poor sleep quality and operationally defined sleep disorders are common in the UK Biobank cohort. Sleep questionnaire data can now be integrated with a range of biomedical information to advance understanding of sleep.

**Statement of significance:** A comprehensive sleep questionnaire was introduced to the UK Biobank, with over 185,000 participants providing data. Overall, respondents reported relatively poor sleep quality; 40% reported sleep duration less than 7 hours, and 25% met criteria for at least one sleep disorder. Enhanced assessment of sleep in UK Biobank now enables integration with extensive biomedical data, including genetic, wearable, imaging, lifestyle, biomarker, and electronic health record data, offering opportunities to investigate the biological and environmental factors that influence sleep and circadian systems, and their impact on health.

## Introduction

Sleep disorders are highly prevalent and burdensome ^1,2^. Despite major advances in understanding and classifying sleep disorders, sleep complaints often remain under-assessed, under-diagnosed, and by extension poorly managed in clinical practice ^3,4^. In addition to disordered sleep, population variation in sleep duration, timing, continuity, regularity, and quality are consistently associated with a range of adverse mental and physical health outcomes ^5^; while optimising sleep dimensions in clinical and non-clinical populations delivers health benefit ^6–8^.

The emergence of large-scale biobank studies with embedded sleep measurement has facilitated novel discoveries; for example, on the genetic architecture of sleep and circadian traits ^9–11^, associations with morbidity and mortality ^12,13^, and potential causality between sleep-circadian parameters and disease through application of Mendelian randomisation ^14,15^. Indeed, to date^1^, over 700 sleep and circadian related studies have been published utilising data from UK Biobank, a cohort study of ∼500,000 people aged 40 to 69 years at enrolment, which integrates clinical, genetic, imaging and other biomedical data with assessments of lifestyle and sociodemographic factors. However, a recurrent criticism levelled at such large-scale studies is the low-resolution measurement of sleep, typically reflecting just single item questions ^16^, as well as limited consideration of the range and co-occurrence of sleep and circadian disorders ^17^.

Ideally, large-scale biobank studies would include both polysomnographic and clinical assessment of sleep and sleep disorders, but currently this is neither practical nor feasible at scale (although recent developments in measurement and analytics may change the future landscape; ^18^). Enhanced capture of probable sleep disorders at scale through comprehensive questionnaire measures would represent a key advance relative to previous work. For example, improved phenotyping would reduce misclassification between related sleep disorders in epidemiological analysis, and more precisely characterise heterogeneity within disorder categories, paving the way for precision medicine approaches. Because sleep disorders often go undiagnosed, and are therefore under-reported in electronic health records, we need measures that capture probable cases to permit comparison with non-cases on genetics, biomarkers, structural and functional brain health, and other disease indices. Doing so will provide significant opportunity to define the patterning, underpinning biology, and putative consequences of a range of sleep and circadian disorders.

Recognising the fundamental importance of sleep and circadian rhythms for health, UK Biobank introduced a dedicated web-based sleep questionnaire in 2023 (with data released in March 2025). The purpose of this paper is to describe the instruments and items included in the questionnaire, introduce suggested phenotype coding for a range of sleep and circadian disorders, and present descriptive statistics on the patterning of sleep disorder cases and sleep/circadian traits.

Our objectives were thus:

1. to describe the characteristics of individuals who completed the sleep questionnaire;
2. to describe variation in sleep health dimensions and the association with sociodemographic factors and disordered sleep;
3. to describe the association between reporting sleep difficulties at the baseline assessment visit using single items (2006-2010) and meeting criteria for disordered sleep on the sleep questionnaire (2023)
4. to describe cohort-wide prevalence of disordered sleep, co-occurrence between different sleep disorders, and the sociodemographic, environmental, clinical and lifestyle correlates of sleep disorders.

## Methods

### UK Biobank

UK Biobank is a prospective cohort study of over half a million adults aged 40-69 years when recruited from England, Scotland and Wales between 2006 and 2010 (response rate 5.5%) (more information can be found in Sudlow et al., 2015 ^19^). At the baseline assessment visit, participants gave informed consent, completed a touchscreen questionnaire, provided anthropometric measures and biological samples, and underwent an interview with a nurse. Sleep and sleep disruption were captured using single-item questions probing sleep duration, difficulties getting up in the morning, napping, dozing, snoring, insomnia, and chronotype.

The baseline assessment visit included consent to be recontacted to take part in further voluntary assessments. All the procedures were approved by the NHS Research Ethics Service (Ref. 11/NW/0382).

### Development of the sleep questionnaire

Between 2017 and 2021, a group of sleep researchers and clinicians worked with the UK Biobank team to design a sleep questionnaire with the aim of capturing key domains of self-reported sleep health and sleep disorders. The online questionnaire comprised 138 items across 11 modules (see Figure 1). When developing the content of the questionnaire, we considered scientific value, the need to balance breadth and depth of assessment, participant acceptability (e.g. time to complete, ease of use, limited item duplication), and licencing considerations. At the time of planning, we judged that there was no existing single questionnaire with adequate psychometric properties to permit identification of all the main sleep and circadian disorder types. We selected published scales commonly used in research and clinical practice to assess common sleep disorders, supplemented by amended or additional questionnaire items (Appendix 1). These comprised the Sleep Condition Indicator ^20^ (insomnia), Berlin Questionnaire ^19^ (obstructive sleep apnoea), Brief Screen for Sleep Disorders ^21^ (delayed and advanced sleep-wake phase disorder), Shift Work Sleep Disorder Questionnaire ^22^ (shift work disorder), Alliance Sleep Questionnaire ^23^ (narcolepsy, parasomnias), REM Sleep Behaviour Disorder Single-Question Screen ^24^ (REM sleep behaviour disorder) and Cambridge-Hopkins Restless Legs Syndrome Questionnaire ^25^ (restless legs syndrome).

**Figure 1.**
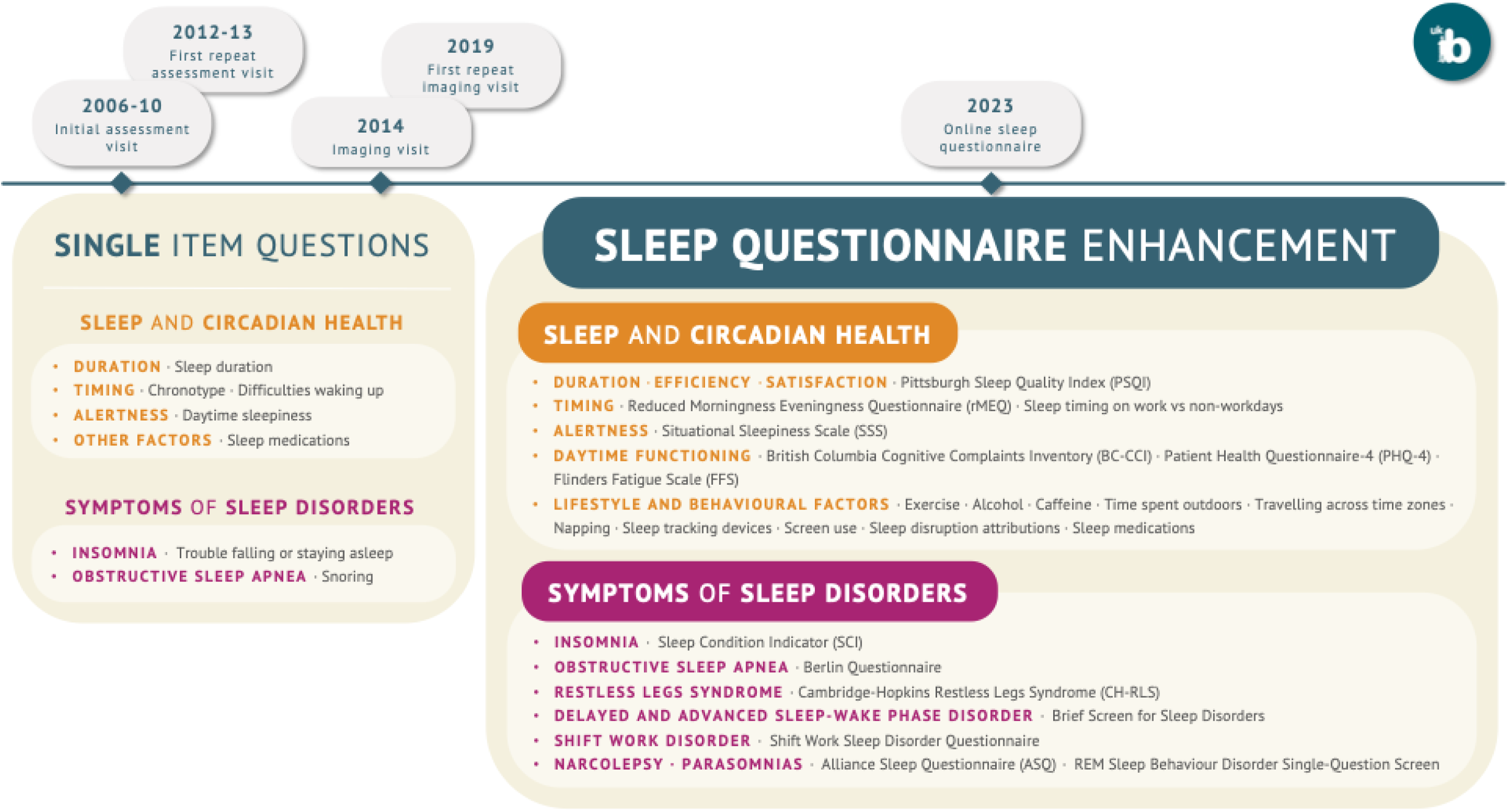
UK Biobank Sleep Questionnaire (2023).

We also assessed sleep and circadian health dimensions and selective potential correlates of sleep, including Pittsburgh Sleep Quality Index ^26^ (PSQI; global sleep quality), the reduced Morningness-Eveningness Questionnaire ^27^ (rMEQ; chronotype), the situational sleepiness scale and bespoke questionnaire items ^28^ (sleepiness), Flinders Fatigue Scale ^29^ (FFS; fatigue), British Columbia Cognitive Complaints Inventory ^30^ (BC-CCI; cognitive impairment), Patient Health Questionnaire-4 ^31^ (PHQ-4; depression and anxiety symptoms) and family history of sleep disorders ^23^. A number of additional bespoke items were added to capture domains where there was no existing questionnaire, or to provide context to included questionnaire items (e.g. duration of early morning awakenings, napping, changes in sleep following the COVID-19 pandemic, shift work pattern, timing of sleep periods on work and non-work days, and key lifestyle and behavioural factors, such as exercise, use of sleep tracking devices, alcohol, and caffeine).

### Questionnaire administration

Participants with an active email address and had not withdrawn from further contact (n = 335,587), received an invitation email containing a hyperlink to complete the online sleep questionnaire. Reminder emails were sent to non-respondents two weeks and four months after the initial invitation, and to partial respondents two weeks after they started the questionnaire.

Participants for whom UK Biobank did not have an email address were encouraged via information on the UK Biobank website, and in the UK Biobank participant newsletter, to complete the online questionnaire by logging-on directly to the participant website.

The questionnaire was piloted in December 2022, administered initially to around 13,000 participants, and resultant data were incorporated into the final dataset. The aim was to ensure that the online platform and procedures were adequately robust and that the questionnaire was acceptable in terms of content and length. Based on pilot feedback, several questions in the ‘work and sleep’ and ‘sleep consequences’ modules were identified as needing revised wording to improve clarity, although the response options remained unchanged. Changes in wording (Appendix 2) between item versions was judged to be sufficiently minor and, for the purposes of the present analysis, responses were grouped. The main phase of questionnaire administration began in late January 2023. Data were accessed in March 2025.

### Defining outcomes/phenotypes from the sleep questionnaire

Using the available questionnaire items, we developed putative case definitions for insomnia disorder, obstructive sleep apnoea, restless legs syndrome, delayed and advanced sleep-wake phase disorder, shift work disorder, and REM and NREM parasomnias. To do this, we drew upon diagnostic criteria (DSM-5 ^32^ or ICSD-3-TR ^33^), original scale scoring, and clinical expertise to arrive at consensus agreement. An initial coding framework was sent to a broader working group of sleep-circadian researchers, who provided comment and refinement, prior to finalising definitions. Given that comprehensive clinical assessment (often involving polysomnography) is needed to appropriately diagnose sleep disorders, our case definitions should be considered suggestive of disorder rather than definitive.

Sleep disorder coding was applied based on overall case definitions. Respondents who met these definitions were classified as likely cases, even if responses to sub-fields were missing. Those who did not meet the case definitions were classified as likely non-case. Respondents with insufficient information to determine case or non-case status were assigned missing values for that phenotype. Changes made to phenotypes during the analysis phase are described in Appendix 3.

Missing data were defined as those who responded “prefer not to answer”. Responses including “do not know” and “not applicable” were considered as missing unless it was appropriate to assign a default value (see Appendix 4 for framework). All “varies significantly” responses were considered as missing when the outcomes required precise clock timings (e.g. calculating sleep efficiency, mid-point of sleep period).

### Analysis

Phenotype coding and descriptive analyses were independently conducted by two authors (HY and CZ), with any discrepancies resolved through discussion. All operationally defined phenotypes were cross-checked at the respondent level, and resulting tabulations were compared by visual inspection.

Baseline characteristics and phenotypes were summarised using descriptive statistics (see Appendix 6 for data fields for all single item variables). Continuous variables were approximately normally distributed and reported as mean (SD), while categorical variables were summarised as frequencies (percentages). Cross-tabulations were used to examine patterns in sleep health dimensions and operationally defined sleep disorders overall and across participant subgroups. Descriptive statistics involving sleep phenotypes were reported as row-wise or column-wise variables in tables. For the statistic in a row-wise or column-wise calculation, the denominator is the same across all columns or rows.

All analyses were performed on the UK Biobank Research Analysis Platform (RAP: https://www.ukbiobank.ac.uk/enable-your-research/research-analysis-platform) using Python (version 3.10) and R (version 4.4). To support transparency and reproducibility, all scripts used in the analysis are publicly available on GitHub (https://github.com/UK-Biobank-Sleep-consortium/ukb_sleep_enhancement). The operationally defined phenotypes will be returned to the UK Biobank Showcase for use by other researchers.

For composite timing phenotypes that required calculations from responses in the form of 24-hour timestamps, a cut-point based inclusion criterion was applied to filter out incorrect responses due to timestamp confusion. If computed habitual time in bed or sleep duration was less than three hours or greater than 16 hours responses used for calculations were set to missing (Appendix 5). Results were reported in accordance with the Strengthening the Reporting of Observational Studies in Epidemiology (STROBE) guidelines (Appendix 7).

## Results

### Respondent characteristics

A total of 185,056 respondents completed at least one module of the sleep questionnaire and were included in the analysis, representing approximately 56% of those invited (Figure 2). Respondents were predominately female (57.9%), from a White ethnic background (96.8%), and had a mean (SD) age of 69.9 (7.5) years (Table 1).

**Figure 2.**
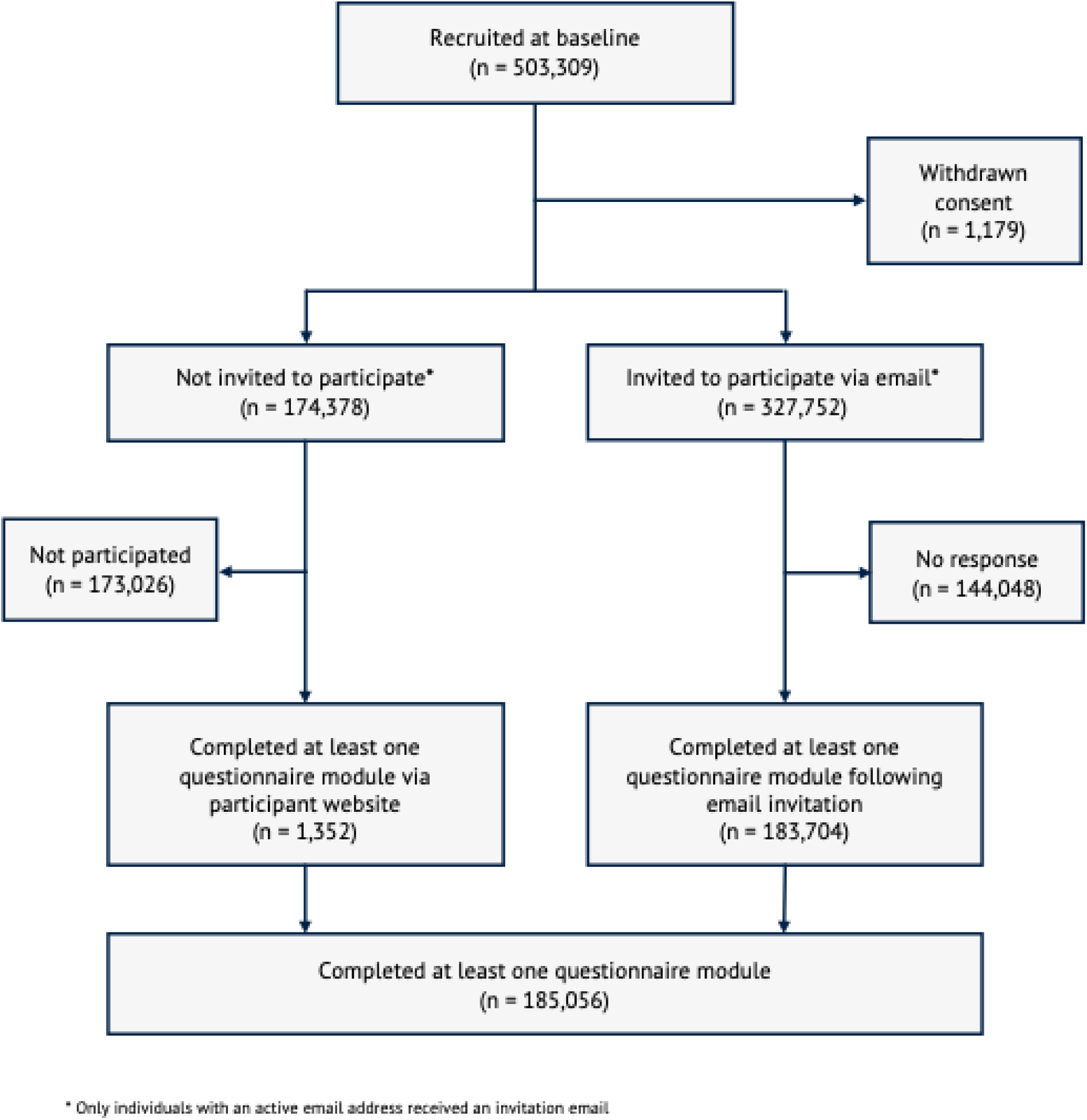
Flowchart of the UK Biobank Sleep Questionnaire.

**Table 1.** Population characteristics at baseline (2006-2010) for UK Biobank Sleep Cohort: participant count and proportions by subgroup.

| Characteristic | Baseline<br>assessment (2006-2010)<br>(n = 502,128) | Sleep<br>questionnaire<br>(2023)<br>(n = 185,056) | Invited but did not<br>participate in sleep<br>questionnaire<br>(n = 144,047) |
| --- | --- | --- | --- |
| **Age at baseline (years)** |  |  |  |
| Mean (SD) | 57.0 (8.1) | 55.8 (7.6) | 56.4 (8.3) |
| **Age at sleep questionnaire**<br>**completion (years)** |  |  |  |
| Mean (SD) | NA (NA) | 69.9 (7.5) | NA (NA) |
| **Sex** |  |  |  |
| Female | 273,155 (54.4%) | 107,071 (57.9%) | 75,303 (52.3%) |
| Male | 228,973 (45.6%) | 77,985 (42.1%) | 68,744 (47.7%) |
| **Ethnicity** |  |  |  |
| Asian or Asian British | 11,443 (2.3%) | 2,063 (1.1%) | 4,167 (2.9%) |
| Black or Black British | 8,048 (1.6%) | 1,294 (0.7%) | 2,804 (1.9%) |
| Mixed Race or Other | 7,502 (1.5%) | 1,996 (1.1%) | 2,476 (1.7%) |
| White | 472,360 (94.1%) | 179,047 (96.8%) | 133,975 (93.0%) |
| Missing | 2,775 (0.6%) | 656 (0.4%) | 625 (0.4%) |
| **Highest qualification** |  |  |  |
| College or University degree | 161,001 (32.1%) | 81,005 (43.8%) | 46,088 (32.0%) |
| Vocational qualifications | 135,324 (27.0%) | 50,578 (27.3%) | 40,855 (28.4%) |
| National exams at ages 16-18 | 110,453 (22.0%) | 38,692 (20.9%) | 34,325 (23.8%) |
| None of the above | 85,228 (17.0%) | 12,886 (7.0%) | 20,471 (14.2%) |
| Missing | 10,122 (2.0%) | 1,895 (1.0%) | 2,308 (1.6%) |
| **Employment status** |  |  |  |
| Employed | 286,640 (57.1%) | 123,177 (66.6%) | 85,899 (59.6%) |
| Not employed | 212,641 (42.3%) | 61,453 (33.2%) | 57,398 (39.8%) |
| Missing | 2,847 (0.6%) | 426 (0.2%) | 750 (0.5%) |
| **Townsend deprivation index** |  |  |  |
| Least Deprived | 186,226 (37.1%) | 76,751 (41.5%) | 56,263 (39.1%) |
| 2nd Quintile | 102,013 (20.3%) | 39,309 (21.2%) | 29,994 (20.8%) |
| 3rd Quintile | 74,880 (14.9%) | 27,699 (15.0%) | 21,327 (14.8%) |
| 4th Quintile | 67,107 (13.4%) | 22,441 (12.1%) | 18,669 (13.0%) |
| Most Deprived | 71,278 (14.2%) | 18,637 (10.1%) | 17,600 (12.2%) |
| Missing | 624 (0.1%) | 219 (0.1%) | 194 (0.1%) |
| **Body mass index** |  |  |  |
| Underweight (<18.5 kg/m<sup>2</sup>) | 2,625 (0.5%) | 1,005 (0.5%) | 600 (0.4%) |
| Normal weight (18.5-24.9 kg/m<sup>2</sup>) | 162,260 (32.3%) | 70,668 (38.2%) | 45,424 (31.5%) |
| Overweight (25-29.9 kg/m<sup>2</sup>) | 211,968 (42.2%) | 76,487 (41.3%) | 62,952 (43.7%) |
| Obese (30+ kg/m<sup>2</sup>) | 122,172 (24.3%) | 36,454 (19.7%) | 34,339 (23.8%) |
| Missing | 3,103 (0.6%) | 442 (0.2%) | 732 (0.5%) |
| **Smoking status** |  |  |  |
| Never | 273,323 (54.4%) | 108,409 (58.6%) | 79,339 (55.1%) |
| Previous | 172,920 (34.4%) | 63,168 (34.1%) | 50,360 (35.0%) |
| Current | 52,937 (10.5%) | 13,017 (7.0%) | 13,686 (9.5%) |
| Missing | 2,948 (0.6%) | 462 (0.2%) | 662 (0.5%) |
| **Alcohol consumption frequency** |  |  |  |
| <1 times/week | 154,373 (30.7%) | 47,328 (25.6%) | 41,920 (29.1%) |
| 1-2 times/week | 129,184 (25.7%) | 47,529 (25.7%) | 37,632 (26.1%) |
| 3-4 times/week | 115,356 (23.0%) | 48,711 (26.3%) | 34,132 (23.7%) |
| Daily | 101,715 (20.3%) | 41,334 (22.3%) | 30,076 (20.9%) |
| Missing | 1,500 (0.3%) | 154 (0.1%) | 287 (0.2%) |
| **Self-reported overall health rating** |  |  |  |
| Excellent | 81,787 (16.3%) | 40,056 (21.6%) | 22,482 (15.6%) |
| Good | 288,816 (57.5%) | 111,924 (60.5%) | 85,562 (59.4%) |
| Fair | 105,281 (21.0%) | 28,549 (15.4%) | 29,990 (20.8%) |
| Poor | 22,759 (4.5%) | 4,092 (2.2%) | 5,269 (3.7%) |
| Missing | 3,485 (0.7%) | 435 (0.2%) | 744 (0.5%) |
| **Self-reported prescribed sleep medication** |  |  |  |
| Yes | 4,829 (1.0%) | 1,167 (0.6%) | 1,204 (0.8%) |
| No | 496,437 (98.9%) | 183,814 (99.3%) | 142,677 (99.0%) |
| Missing | 862 (0.2%) | 75 (0.0%) | 166 (0.1%) |
| **Participation in UK Biobank Mental Health Questionnaire (2016-2017)** |  |  |  |
| Yes | 157,255 (31.3%) | 117,550 (63.5%) | 30,173 (20.9%) |
| No | 344,873 (68.7%) | 67,506 (36.5%) | 113,874 (79.1%) |
| **Participation in UK Biobank Accelerometry (2013-2015)** |  |  |  |
| Yes | 103,610 (20.6%) | 71,900 (38.9%) | 23,643 (16.4%) |
| No | 398,518 (79.4%) | 113,156 (61.1%) | 120,404 (83.6%) |
| **Participation in UK Biobank Imaging (2014-ongoing)** |  |  |  |
| Yes | 92,031 (18.3%) | 71,801 (38.8%) | 15,978 (11.1%) |
| No | 410,097 (81.7%) | 113,255 (61.2%) | 128,069 (88.9%) |

Compared to individuals who received an invitation but did not participate, sleep questionnaire respondents were more likely to be female (57.9% vs 52.3%), from a White ethnic background (96.8% vs 93.0%), and have a College or University degree (43.8% vs 32.0%, Table 1), Moreover, sleep questionnaire respondents were more likely to be in the ‘healthy’ BMI range (38.2% vs 31.5%), have never smoked (58.6% vs 55.1%) and report ‘excellent’ overall health (21.6% vs 15.6%) at the baseline assessment (Table 1). A similar pattern was also observed when comparing sleep questionnaire respondents to the broader UK Biobank cohort (Table 1). Sleep parameters collected at baseline were broadly similar between respondents and non-respondents (eTable 1). Respondents were more likely to have participated in the UK Biobank mental health questionnaire (63.5% vs 20.9%), accelerometery (38.9% vs 16.4%) and imaging assessment (38.8% vs 11.1%).

For brevity, below we provide an illustrative summary of descriptive data patterns but see Tables 1-5 and eTables 1-7 for complete information on all examined variables and outcomes.

### Sleep health dimensions

Mean sleep duration (SD) was 6.9 hours (1.2 hours) with 53.5% of respondents reporting ‘optimal’ sleep duration (7-9 hours), while 39.5% reported ‘short’ (<7hrs) and 4.6% reported ‘long’ (>9 hrs) sleep duration (eTable 2). 74.6% reported ‘fairly good’ or ‘good’ sleep quality in the past month. Mean [SD] global sleep quality score (total PSQI) was 6.65 [3.46], with 48.8% scoring in the sleep disturbance range (>5), and mean sleep efficiency was 79.1% (SD, 13.3%; eTable 2). The majority (73.6%) of respondents did not work, but those engaged in employment (and worked typical hours of 9am-5pm, n = 35,632, 19.3%) woke 53 minutes later on free-days compared to work-days in the past month (eTable 3). For shift-workers (n = 10,908, 5.9%), sleep timing on work-days in the past month followed a predictable pattern (e.g. morning shift workers had advanced sleep onset and offset times, while night-shift workers had markedly delayed sleep onset and offset times).

Sleep efficiency was lower in participants with advancing age, lower levels of education, and poorer self-rated health (eTable 2). Short sleep duration was more common amongst individuals with Asian, Black or a Mixed ethnic background; with lower levels of education; those living in the most deprived geographical areas; those in the ‘obese’ BMI range; and those with ‘poor’ or ‘fairly poor’ health (eTable 2). Global sleep quality score (PSQI) was also higher (indicating worse sleep quality) in participants with lower levels of educational qualification and less frequent alcohol consumption, higher levels of deprivation, BMI, smoking status and poorer self-rated health status (eTable 2). Females were more likely to have shorter sleep duration, lower sleep efficiency, and poorer global sleep quality than males. Individuals who were not employed or taking sleep medications at baseline (2006-2010) had poorer global PSQI sleep quality score (eTable 2) compared to those who worked and did not take sleep medications.

### Operationally-defined sleep disorders – prevalence, baseline characteristics and co-occurrence

Around one-quarter (25.2%) of respondents met criteria for at least one sleep disorder, while 7.3% met criteria for at least two sleep disorders (Table 2). Chronic insomnia disorder was the most common sleep disorder (14.4%) followed by obstructive sleep apnoea (OSA; 8.0%), restless legs syndrome (RLS; 4.1%), nightmare disorder (3.7%), REM sleep behaviour disorder (RBD; 1.2%), sleepwalking (0.9%), shift work disorder (SWD; 0.9% of respondents/ 15.7% of shift workers), and delayed and advanced sleep-wake phase disorder (DSWPD, 0.7% and ASWPD, 0.5%) (Table 2).

**Table 2.** Prevalence of operationally defined sleep disorders by sex and current age.

|  | n (%) | Overall |  | Age, n (%) |  | Sex, n (%) |  |
| --- | --- | --- | --- | --- | --- | --- | --- |
|  |  | n (%)<sup>a</sup> | Age in years (SD) | <65 years | ≥65 years | Female | Male |
| **Chronic Insomnia Disorder** |  | 26,729 (14.4%) | 69.1 (7.7) | 8,823 (33.0) | 17,906 (67.0) | 18,342 (68.6) | 8,387 (31.4) |
| **Obstructive Sleep Apnoea** |  | 14,796 (8.0%) | 67.7 (7.6) | 5,887 (39.8) | 8,909 (60.2) | 8,513 (57.5) | 6,283 (42.5) |
| **Restless Legs Syndrome** |  | 7,506 (4.1%) | 69.7 (7.5) | 2,193 (29.2) | 5,313 (70.8) | 5,456 (72.7) | 2,050 (27.3) |
| **Delayed Sleep-Wake Phase Disorder** |  | 1,207 (0.7%) | 67.9 (7.6) | 452 (37.4) | 755 (62.6) | 765 (63.4) | 442 (36.6) |
| **Advanced Sleep-Wake Phase Disorder** |  | 903 (0.5%) | 69.6 (7.5) | 274 (30.3) | 629 (69.7) | 502 (55.6) | 401 (44.4) |
| **Shift Work Disorder** |  | 1,717 (0.9%) | 62.2 (5.9) | 1,289 (75.1) | 428 (24.9) | 968 (56.4) | 749 (43.6) |
| **Sleepwalking** |  | 1,609 (0.9%) | 70.3 (7.7) | 448 (27.8) | 1,161 (72.2) | 820 (51.0) | 789 (49.0) |
| **REM Sleep Behaviour Disorder** |  | 2,157 (1.2%) | 69.6 (7.5) | 657 (30.5) | 1,500 (69.5) | 604 (28.0) | 1,553 (72.0) |
| **Nightmares** |  | 6,805 (3.7%) | 68.9 (7.6) | 2,272 (33.4) | 4,533 (66.6) | 3,721 (54.7) | 3,084 (45.3) |
| **Summary** |  |  |  |  |  |  |  |
| None of the above |  | 138,445 (74.8) | 70.2 (7.5) | 36,903 (26.7) | 101,542 (73.3) | 77,658 (56.1) | 60,787 (43.9) |
| ≥1 of above |  | 46,611 (25.2) | 69.1 (7.7) | 15,341 (32.9) | 31,270 (67.1) | 29,413 (63.1) | 17,198 (36.9) |
| ≥2 of above |  | 13,462 (7.3) | 67.8 (7.6) | 5,327 (39.6) | 8,135 (60.4) | 8,371 (62.2) | 5,091 (37.8) |
| ≥3 of above |  | 2,842 (1.5) | 66.4 (7.4) | 1,344 (47.3) | 1,498 (52.7) | 1,643 (57.8) | 1,199 (42.2) |

<sup>a</sup> The percentage (%) is calculated by dividing n by the total sleep questionnaire respondents (n = 185,056).

There was a particularly high proportion of female (vs male) cases for insomnia (68.6%), RLS (72.7%) and DSWPD (63.4%) (Table 2), while conversely possible RBD was more frequently observed in males (72.0%) (Table 2).

In terms of co-occurrence of sleep disorders, around a quarter of insomnia cases (26.1%) met criteria for OSA (Table 3). Insomnia was also common in participants meeting criteria for OSA (47.2%), SWD (54.5%), RLS (30.2%) and nightmares (38.8%) (Table 3).

**Table 3.** Cooccurrence between operationally defined sleep disorders.

| n (%) | Overall prevalence<sup>a</sup> | Comorbidity |  |  |  |  |  |  |  |  |
| --- | --- | --- | --- | --- | --- | --- | --- | --- | --- | --- |
|  |  | Chronic Insomnia Disorder | Obstructive Sleep Apnoea | Restless Legs Syndrome | Delayed Sleep-Wake Phase Disorder | Advanced Sleep-Wake Phase Disorder | Shift Work Disorder | Sleepwalking | REM Sleep Behaviour Disorder | Nightmares |
| **Chronic Insomnia Disorder** | 26,729 (14.4%) |  | 6,986 (26.1%) | 2,268 (8.5%) | 310 (1.2%) | 141 (0.5%) | 935 (3.5%) | 372 (1.4%) | 574 (2.1%) | 2,638 (9.9%) |
| **Obstructive Sleep Apnoea** | 14,796 (8.0%) | 6,986 (47.2%) |  | 1,228 (8.3%) | 333 (2.3%) | 43 (0.3%) | 546 (3.7%) | 202 (1.4%) | 474 (3.2%) | 1,480 (10.0%) |
| **Restless Legs Syndrome** | 7,506 (4.1%) | 2,268 (30.2%) | 1,228 (16.4%) |  | 90 (1.2%) | 27 (0.4%) | 110 (1.5%) | 72 (1.0%) | 158 (2.1%) | 451 (6.0%) |
| **Delayed Sleep-Wake Phase Disorder** | 1,207 (0.7%) | 310 (25.7%) | 333 (27.6%) | 90 (7.5%) |  | 0 (0.0%) | 40 (3.3%) | 14 (1.2%) | 29 (2.4%) | 106 (8.8%) |
| **Advanced Sleep-Wake Phase Disorder** | 903 (0.5%) | 141 (15.6%) | 43 (4.8%) | 27 (3.0%) | 0 (0.0%) |  | 11 (1.2%) | 18 (2.0%) | 7 (0.8%) | 48 (5.3%) |
| **Shift Work Disorder** | 1,717 (0.9%) | 935 (54.5%) | 546 (31.8%) | 110 (6.4%) | 40 (2.3%) | 11 (0.6%) |  | 36 (2.1%) | 50 (2.9%) | 191 (11.1%) |
| **Sleepwalking** | 1,609 (0.9%) | 372 (23.1%) | 202 (12.6%) | 72 (4.5%) | 14 (0.9%) | 18 (1.1%) | 36 (2.2%) |  | 92 (5.7%) | 135 (8.4%) |
| **REM Sleep Behaviour Disorder** | 2,157 (1.2%) | 574 (26.6%) | 474 (22.0%) | 158 (7.3%) | 29 (1.3%) | 7 (0.3%) | 50 (2.3%) | 92 (4.3%) |  | 539 (25.0%) |
| **Nightmares** | 6,805 (3.7%) | 2,638 (38.8%) | 1,480 (21.7%) | 451 (6.6%) | 106 (1.6%) | 48 (0.7%) | 191 (2.8%) | 135 (2.0%) | 539 (7.9%) |  |

<sup>a</sup> Prevalence (%) is calculated by dividing the number of cases by the total sleep questionnaire respondents (n = 185,056).

Across all possible sleep disorders, compared to the overall sample, there was a higher prevalence of individuals who, at baseline (2006-2010), were in the obese BMI range (21.4-35.7% vs. 19.7%, eTable 4), reported having frequent insomnia (29.8-50.5% vs. 26.6%), reported frequent daytime sleepiness (2.7-5.2% vs. 2.1%) and had sleep duration <7 hrs (24.1-38.5% vs 22.3%, eTable 5). With the exception of ASWPD, individuals meeting criteria for any operationally defined sleep disorder, compared to the overall sample, were more likely to report baseline evening chronotype (8.6-57.2% vs. 8.3%; ASWPD 0.4%) and difficulties getting up in the morning (not at all easy, 4.6-25.5% vs. 3.4%; ASWPD 0.6%, eTable 5).

### Operationally defined sleep disorders – sleep health dimensions and lifestyle factors

Consistent with case criteria, those with operationally defined sleep disorders were characterised by poorer sleep quality and higher levels of daytime sleepiness relative to the overall sample (Table 4). Short sleep duration (< 7hrs) was most prominent in those with possible insomnia (76.4%) and shift work disorder (70.5%, Table 4). Sleep timing was earliest in those with possible ASWPD (midpoint of sleep period, 02:31am) and latest in those with DSWPD (midpoint of sleep period, 05:31, Table 4). Of all possible sleep disorders, insomnia had the lowest sleep efficiency (mean [SD], 67.2% [13.8]), while DSWPD had the highest (mean [SD], 79.2% [14.3]; Table 4). Respondents with insomnia also had the highest PSQI score (mean [SD], 11.1 [3.0]), while those with ASWPD had the lowest (6.9 [2.8]) (Table 4).

**Table 4.**
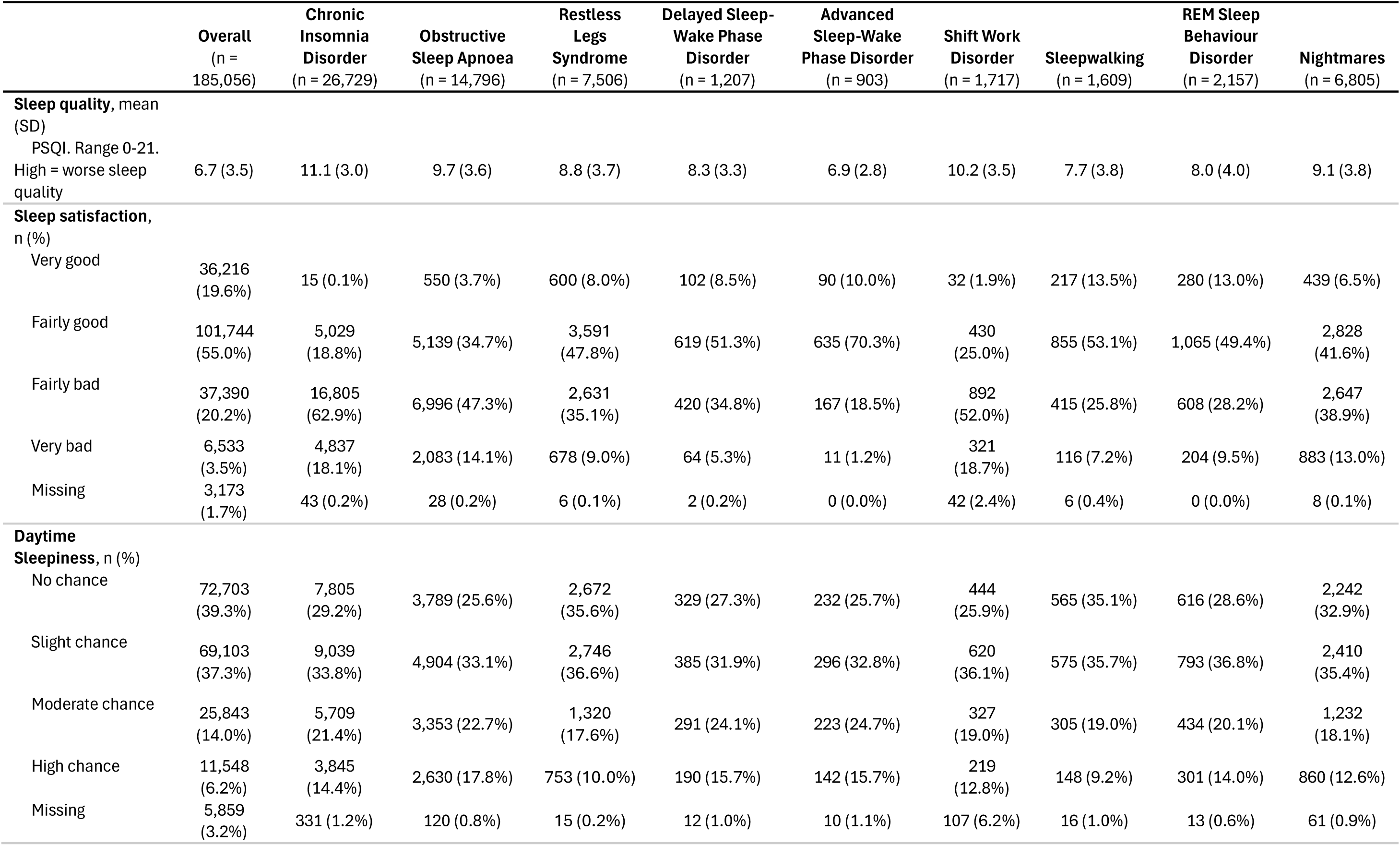
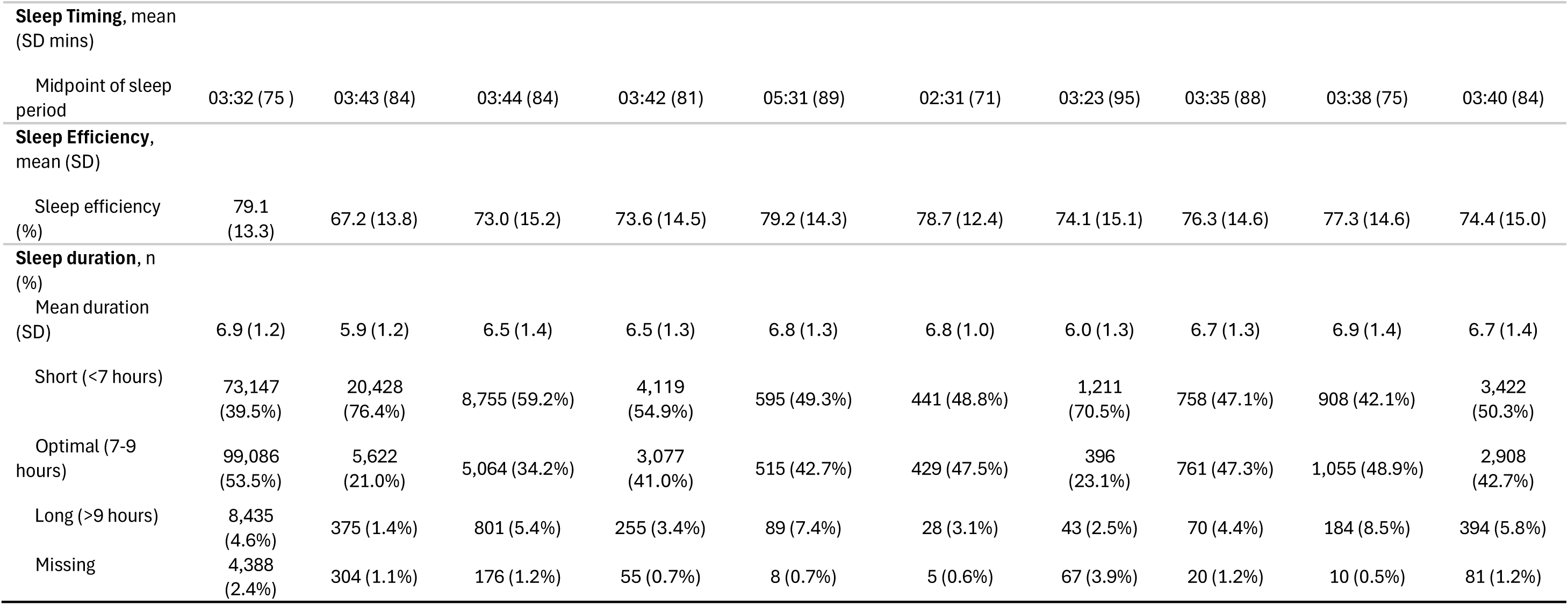
Sleep health dimensions by operationally defined sleep disorders.

In terms of lifestyle factors, compared to overall respondents, a greater proportion of individuals with sleep disorders napped daily (7.1-15.0% vs. 6.7%), did not exercise in the past month (16.5-25.0% vs. 14.4%), and used electronic devices in bed (24.0-35.2% vs. 22.6%, Table 5). Those with DSWPD, compared to overall respondents and other sleep disorders, were more likely to spend less than 1 hour per day outdoors in summer (14.7% vs. 4.5% vs. 5.5-10.3%) and in winter (47.8% vs. 23.4% vs. 22.6-34.9%, Table 5). Individuals with operationally defined sleep disorders tended to report greater cognitive difficulties (range of mean BC-CCI values: 4.5-7.0 vs. 3.8), fatigue (range of mean FFS values: 6.3-14.0 vs. 5.8), depression (range of mean PHQ-2 values: 0.8-2.3 vs. 0.7) and anxiety (range of mean GAD-2 values: 1.2-2.5 vs. 1.0, eTable 6). Individuals with SWD were more likely to report being involved in ≥1 accident or near-miss due to sleepiness in the past year compared to people with other operationally defined sleep disorders (10.0% vs. 4.0-7.0%, eTable 6).

**Table 5.** Chronotype and lifestyle related factors by operationally defined sleep disorders.

|  | Overall<br>(n =<br>185,056) | Chronic<br>Insomnia<br>Disorder<br>(n = 26,729) | Obstructive<br>Sleep Apnoea<br>(n = 14,796) | Restless<br>Legs<br>Syndrome<br>(n = 7,506) | Delayed<br>Sleep-Wake<br>Phase<br>Disorder<br>(n = 1,207) | Advanced<br>Sleep-Wake<br>Phase Disorder<br>(n = 903) | Shift Work<br>Disorder<br>(n = 1,717) | Sleepwalking<br>(n = 1,609) | REM Sleep<br>Behaviour<br>Disorder<br>(n = 2,157) | Nightmare<br>Disorder<br>(n = 6,805) |
| --- | --- | --- | --- | --- | --- | --- | --- | --- | --- | --- |
| **Chronotype** |  |  |  |  |  |  |  |  |  |  |
| Definitely morning<br>type | 7,193<br>(3.9%) | 770 (2.9%) | 235 (1.6%) | 264 (3.5%) | 0 (0.0%) | 258 (28.6%) | 65 (3.8%) | 63 (3.9%) | 56 (2.6%) | 215 (3.2%) |
| Moderately morning<br>type | 57,340<br>(31.0%) | 7,297<br>(27.3%) | 3,169 (21.4%) | 2,197<br>(29.3%) | 0 (0.0%) | 645 (71.4%) | 407<br>(23.7%) | 513 (31.9%) | 565 (26.2%) | 1,837<br>(27.0%) |
| Neither type | 104,714<br>(56.6%) | 14,823<br>(55.5%) | 8,586 (58.0%) | 4,199<br>(55.9%) | 0 (0.0%) | 0 (0.0%) | 872<br>(50.8%) | 847 (52.6%) | 1,229 (57.0%) | 3,699<br>(54.4%) |
| Moderately evening<br>type | 10,415<br>(5.6%) | 2,277 (8.5%) | 1,815 (12.3%) | 574 (7.6%) | 1,026 (85.0%) | 0 (0.0%) | 223<br>(13.0%) | 117 (7.3%) | 182 (8.4%) | 665 (9.8%) |
| Definitely evening<br>type | 566<br>(0.3%) | 219 (0.8%) | 168 (1.1%) | 52 (0.7%) | 181 (15.0%) | 0 (0.0%) | 32 (1.9%) | 6 (0.4%) | 20 (0.9%) | 81 (1.2%) |
| Missing | 4,828<br>(2.6%) | 1,343 (5.0%) | 823 (5.6%) | 220 (2.9%) | 0 (0.0%) | 0 (0.0%) | 118 (6.9%) | 63 (3.9%) | 105 (4.9%) | 308 (4.5%) |
| **Time spent outdoors**<br>**per day in summer** |  |  |  |  |  |  |  |  |  |  |
| Less than 1 hour | 8,239<br>(4.5%) | 1,971 (7.4%) | 1,379 (9.3%) | 415 (5.5%) | 178 (14.7%) | 63 (7.0%) | 176<br>(10.3%) | 102 (6.3%) | 137 (6.4%) | 532 (7.8%) |
| 1-<2 hours | 4,971<br>(2.7%) | 818 (3.1%) | 520 (3.5%) | 165 (2.2%) | 60 (5.0%) | 19 (2.1%) | 81 (4.7%) | 36 (2.2%) | 55 (2.5%) | 223 (3.3%) |
| 2-<3 hours | 24,270<br>(13.1%) | 3,776<br>(14.1%) | 2,283 (15.4%) | 975 (13.0%) | 205 (17.0%) | 92 (10.2%) | 289<br>(16.8%) | 208 (12.9%) | 264 (12.2%) | 913 (13.4%) |
| 3+ hours | 124,895<br>(67.5%) | 17,012<br>(63.6%) | 8,861 (59.9%) | 5,322<br>(70.9%) | 625 (51.8%) | 662 (73.3%) | 872<br>(50.8%) | 1,104<br>(68.6%) | 1,530 (70.9%) | 4,427<br>(65.1%) |
| Missing | 22,681<br>(12.3%) | 3,152<br>(11.8%) | 1,753 (11.8%) | 629 (8.4%) | 139 (11.5%) | 67 (7.4%) | 299<br>(17.4%) | 159 (9.9%) | 171 (7.9%) | 710 (10.4%) |
| **Time spent outdoors**<br>**per day in winter** |  |  |  |  |  |  |  |  |  |  |
| Less than 1 hour | 43,325<br>(23.4%) | 8,229<br>(30.8%) | 5,171 (34.9%) | 2,126<br>(28.3%) | 577 (47.8%) | 217 (24.0%) | 501<br>(29.2%) | 364 (22.6%) | 573 (26.6%) | 2,004<br>(29.4%) |
| 1-<2 hours | 41,181<br>(22.3%) | 5,781<br>(21.6%) | 3,081 (20.8%) | 1,728<br>(23.0%) | 229 (19.0%) | 165 (18.3%) | 317<br>(18.5%) | 345 (21.4%) | 455 (21.1%) | 1,456<br>(21.4%) |
| 2-<3 hours | 46,164<br>(24.9%) | 6,013<br>(22.5%) | 3,040 (20.5%) | 1,868<br>(24.9%) | 189 (15.7%) | 227 (25.1%) | 317<br>(18.5%) | 402 (25.0%) | 536 (24.8%) | 1,557<br>(22.9%) |
| 3+ hours | 35,288<br>(19.1%) | 4,324<br>(16.2%) | 2,220 (15.0%) | 1,314<br>(17.5%) | 122 (10.1%) | 237 (26.2%) | 320<br>(18.6%) | 372 (23.1%) | 465 (21.6%) | 1,256<br>(18.5%) |
| Missing | 19,098<br>(10.3%) | 2,382 (8.9%) | 1,284 (8.7%) | 470 (6.3%) | 90 (7.5%) | 57 (6.3%) | 262<br>(15.3%) | 126 (7.8%) | 128 (5.9%) | 532 (7.8%) |
| **Use of electronic devices before bedtime** |  |  |  |  |  |  |  |  |  |  |
| I use them in bed | 41,798<br>(22.6%) | 7,348<br>(27.5%) | 4,483 (30.3%) | 1,874<br>(25.0%) | 425 (35.2%) | 253 (28.0%) | 546<br>(31.8%) | 392 (24.4%) | 518 (24.0%) | 1,805<br>(26.5%) |
| Less than 1 hour | 103,937<br>(56.2%) | 13,961<br>(52.2%) | 7,678 (51.9%) | 4,222<br>(56.2%) | 608 (50.4%) | 436 (48.3%) | 800<br>(46.6%) | 796 (49.5%) | 1,214 (56.3%) | 3,664<br>(53.8%) |
| 1-2 hours | 21,400<br>(11.6%) | 3,433<br>(12.8%) | 1,576 (10.7%) | 969 (12.9%) | 100 (8.3%) | 108 (12.0%) | 183<br>(10.7%) | 232 (14.4%) | 258 (12.0%) | 831 (12.2%) |
| 2-3 hours | 5,217<br>(2.8%) | 729 (2.7%) | 399 (2.7%) | 206 (2.7%) | 28 (2.3%) | 43 (4.8%) | 38 (2.2%) | 74 (4.6%) | 76 (3.5%) | 189 (2.8%) |
| 3 hours or longer | 4,848<br>(2.6%) | 672 (2.5%) | 360 (2.4%) | 165 (2.2%) | 24 (2.0%) | 34 (3.8%) | 26 (1.5%) | 64 (4.0%) | 48 (2.2%) | 183 (2.7%) |
| Not applicable | 1,769<br>(1.0%) | 212 (0.8%) | 151 (1.0%) | 49 (0.7%) | 8 (0.7%) | 19 (2.1%) | 14 (0.8%) | 27 (1.7%) | 24 (1.1%) | 62 (0.9%) |
| Missing | 6,087<br>(3.3%) | 374 (1.4%) | 149 (1.0%) | 21 (0.3%) | 14 (1.2%) | 10 (1.1%) | 110 (6.4%) | 24 (1.5%) | 19 (0.9%) | 71 (1.0%) |
| **Exercise frequency in the past month** |  |  |  |  |  |  |  |  |  |  |
| Daily | 35,980<br>(19.4%) | 3,984<br>(14.9%) | 2,066 (14.0%) | 1,328<br>(17.7%) | 146 (12.1%) | 224 (24.8%) | 208<br>(12.1%) | 313 (19.5%) | 398 (18.5%) | 1,290<br>(19.0%) |
| More than once a week | 44,552<br>(24.1%) | 5,411<br>(20.2%) | 2,724 (18.4%) | 1,631<br>(21.7%) | 249 (20.6%) | 168 (18.6%) | 305<br>(17.8%) | 349 (21.7%) | 467 (21.7%) | 1,389<br>(20.4%) |
| 3-4 times | 39,386<br>(21.3%) | 5,878<br>(22.0%) | 2,982 (20.2%) | 1,639<br>(21.8%) | 200 (16.6%) | 193 (21.4%) | 346<br>(20.2%) | 339 (21.1%) | 462 (21.4%) | 1,505<br>(22.1%) |
| 1-2 times | 27,683<br>(15.0%) | 4,760<br>(17.8%) | 2,613 (17.7%) | 1,343<br>(17.9%) | 212 (17.6%) | 124 (13.7%) | 323<br>(18.8%) | 258 (16.0%) | 336 (15.6%) | 1,098<br>(16.1%) |
| Not at all | 26,612<br>(14.4%) | 4,878<br>(18.2%) | 3,337 (22.6%) | 1,244<br>(16.6%) | 302 (25.0%) | 167 (18.5%) | 368<br>(21.4%) | 265 (16.5%) | 364 (16.9%) | 1,136<br>(16.7%) |
| Unable to exercise | 4,383<br>(2.4%) | 1,383 (5.2%) | 899 (6.1%) | 292 (3.9%) | 82 (6.8%) | 15 (1.7%) | 51 (3.0%) | 58 (3.6%) | 109 (5.1%) | 294 (4.3%) |
| Missing | 6,460<br>(3.5%) | 435 (1.6%) | 175 (1.2%) | 29 (0.4%) | 16 (1.3%) | 12 (1.3%) | 116 (6.8%) | 27 (1.7%) | 21 (1.0%) | 93 (1.4%) |
| **Nap frequency in the past month** |  |  |  |  |  |  |  |  |  |  |
| Daily | 12,317<br>(6.7%) | 2,932<br>(11.0%) | 2,220 (15.0%) | 655 (8.7%) | 164 (13.6%) | 117 (13.0%) | 122 (7.1%) | 149 (9.3%) | 259 (12.0%) | 873 (12.8%) |
| More than once a week | 25,368<br>(13.7%) | 4,268<br>(16.0%) | 2,685 (18.1%) | 1,095<br>(14.6%) | 233 (19.3%) | 176 (19.5%) | 285<br>(16.6%) | 240 (14.9%) | 358 (16.6%) | 1,089<br>(16.0%) |
| 3-4 times | 23,506<br>(12.7%) | 4,927<br>(18.4%) | 2,979 (20.1%) | 1,138<br>(15.2%) | 223 (18.5%) | 168 (18.6%) | 316<br>(18.4%) | 251 (15.6%) | 413 (19.1%) | 1,230<br>(18.1%) |
| 1-2 times | 56,108<br>(30.3%) | 7,577<br>(28.3%) | 3,845 (26.0%) | 2,301<br>(30.7%) | 312 (25.8%) | 240 (26.6%) | 475<br>(27.7%) | 478 (29.7%) | 617 (28.6%) | 1,860<br>(27.3%) |
| Not at all | 61,587<br>(33.3%) | 6,639<br>(24.8%) | 2,916 (19.7%) | 2,293<br>(30.5%) | 260 (21.5%) | 191 (21.2%) | 405<br>(23.6%) | 467 (29.0%) | 495 (22.9%) | 1,684<br>(24.7%) |
| Missing | 6,170<br>(3.3%) | 386 (1.4%) | 151 (1.0%) | 24 (0.3%) | 15 (1.2%) | 11 (1.2%) | 114 (6.6%) | 24 (1.5%) | 15 (0.7%) | 69 (1.0%) |
| **Consumed alcohol to fall asleep in the past month** |  |  |  |  |  |  |  |  |  |  |
| Daily | 2,512<br>(1.4%) | 620 (2.3%) | 415 (2.8%) | 108 (1.4%) | 53 (4.4%) | 16 (1.8%) | 46 (2.7%) | 33 (2.1%) | 71 (3.3%) | 245 (3.6%) |
| More than once a week | 4,323<br>(2.3%) | 1,143 (4.3%) | 601 (4.1%) | 222 (3.0%) | 51 (4.2%) | 24 (2.7%) | 95 (5.5%) | 51 (3.2%) | 101 (4.7%) | 325 (4.8%) |
| 3-4 times | 3,385<br>(1.8%) | 1,171 (4.4%) | 598 (4.0%) | 177 (2.4%) | 46 (3.8%) | 32 (3.5%) | 76 (4.4%) | 36 (2.2%) | 76 (3.5%) | 315 (4.6%) |
| 1-2 times | 9,884<br>(5.3%) | 2,787<br>(10.4%) | 1,346 (9.1%) | 568 (7.6%) | 129 (10.7%) | 47 (5.2%) | 214<br>(12.5%) | 124 (7.7%) | 183 (8.5%) | 589 (8.7%) |
| Not at all | 158,517<br>(85.7%) | 20,583<br>(77.0%) | 11,677 (78.9%) | 6,399<br>(85.3%) | 911 (75.5%) | 774 (85.7%) | 1,175<br>(68.4%) | 1,341<br>(83.3%) | 1,709 (79.2%) | 5,244<br>(77.1%) |
| Missing | 6,435<br>(3.5%) | 425 (1.6%) | 159 (1.1%) | 32 (0.4%) | 17 (1.4%) | 10 (1.1%) | 111 (6.5%) | 24 (1.5%) | 17 (0.8%) | 87 (1.3%) |

| **Caffeine consumption** |  |  |  |  |  |  |  |  |  |  |
| --- | --- | --- | --- | --- | --- | --- | --- | --- | --- | --- |
| Non-drinker | 26,423<br>(14.3%) | 4,815<br>(18.0%) | 2,572 (17.4%) | 1,331<br>(17.7%) | 208 (17.2%) | 169 (18.7%) | 245<br>(14.3%) | 291 (18.1%) | 355 (16.5%) | 1,210<br>(17.8%) |
| 1-3 servings/day | 66,944<br>(36.2%) | 10,095<br>(37.8%) | 5,217 (35.3%) | 2,626<br>(35.0%) | 421 (34.9%) | 319 (35.3%) | 584<br>(34.0%) | 564 (35.1%) | 761 (35.3%) | 2,543<br>(37.4%) |
| 4+ servings/day | 85,317<br>(46.1%) | 11,384<br>(42.6%) | 6,830 (46.2%) | 3,513<br>(46.8%) | 559 (46.3%) | 403 (44.6%) | 773<br>(45.0%) | 727 (45.2%) | 1,021 (47.3%) | 2,966<br>(43.6%) |
| Missing | 6,372<br>(3.4%) | 435 (1.6%) | 177 (1.2%) | 36 (0.5%) | 19 (1.6%) | 12 (1.3%) | 115 (6.7%) | 27 (1.7%) | 20 (0.9%) | 86 (1.3%) |

### Family history of sleep disorders

There was an over-representation of family history of sleep disorders in cases vs non-case. For example, 34.1% of those with RLS reported positive family history of RLS compared to 11.2% of those without RLS; 31.5% of insomnia cases reported positive family history of insomnia compared to 18.8% of those without; 13.5% of OSA cases reported positive family history of OSA compared to 6.9% of those without; and 11.9% of those with nightmare disorder reported positive family history of nightmare disorder compared to 5.4% of those without (see eTable 7).

## Discussion

We introduce a data resource for sleep and circadian research, which is now accessible for integrative and longitudinal data analyses. The main strength of this descriptive study lies in its enhanced characterisation of sleep and circadian health - and their disruption - within the UK Biobank, offering insights that were previously inaccessible due to low-resolution measurement. Very few cohort or population-based studies concurrently assess multiple sleep and circadian disorders. Importantly, these data can now be directly linked to a wide range of additional biomedical data, including genetic, lifestyle, and health information, as well as biological samples, to provide deeper insight into the aetiology of disordered sleep and relationships with disease.

Sleep questionnaire respondents tended to be better educated, healthier, and from a higher socioeconomic background than those who did not participate, suggesting a possible ‘healthy volunteer’ selection bias within the UK Biobank cohort (which is known to be somewhat healthier than the general population) ^34,35^. Patterning of sleep health by sociodemographic factors was broadly consistent with previous work ^36,37^. Overall, the sample reported poorer global sleep quality scores relative to population-based studies in other European countries (e.g. ^38,39^). It is, of course, possible that those experiencing poor sleep were more motivated to complete the questionnaire. However, rates of possible sleep disorder, such as insomnia ^40^ and RLS ^41^, were broadly consistent with the general population literature, supporting the face validity of our case criteria, but our findings cannot be interpreted as general population prevalence estimates. Sex differences were apparent for several disorders, and consistent with contemporary literature, but we observed an unexpected preponderance of females versus males in possible OSA cases, which deserves further investigation. Reinforcing previous work ^42,43^, disordered sleep appeared to be associated with cognitive impairment, fatigue, anxiety, and depression.

Findings reported here should be interpreted with caution due to the descriptive nature of the study; no statistical analyses or covariate adjustments were performed. Operationally defined sleep disorders were derived from self-report measures and there was no confirmation from clinical interviews, polysomnography, or prospective sleep-wake monitoring. This raises the possibility that some disorders are underestimated, while others may be overestimated. Questions mainly focused on current symptoms, or symptoms within the past year, and thus we did not capture lifetime episodes, diagnoses, or treatment history. While we deployed validated questionnaires, it was necessary in some instances to create alternative scoring criteria or include additional items to index possible disorder. Some disorders were defined based on limited descriptive features (e.g. DSWPD), indexed by just single questionnaire items focused on frequency of specific symptoms (e.g. sleepwalking, nightmares), or comprised a combination of items with uncertain predictive validity (e.g. using two defined categories from the Berlin questionnaire for OSA). This notwithstanding, sleep health profiles patterned in the expected direction across cases (e.g. extreme sleep timing for circadian sleep-wake phase disorders, excessive daytime sleepiness for obstructive sleep apnoea) and there was clear evidence of disrupted sleep at the baseline assessment (approximately 14 years earlier) in those who subsequently met case criteria on the 2023 sleep questionnaire. Future data linkage, including formal diagnosis by a healthcare professional, may be needed to confidently capture rare sleep disorders, like narcolepsy.

To retain as much data as possible, we created case/non-case definitions that accounted for ambiguous responses such as ‘prefer not to answer,’ ‘varies significantly,’ ‘do not know,’ and ‘not applicable’, as well as missing responses from nested questions. These responses were typically not part of the original validated questionnaires and required additional consideration during data processing. When handling atypical sleep-timing responses - likely reflecting confusion between 12-hour versus 24-hour format - we cross-checked with other questionnaire items and corrected responses where appropriate. Researchers planning to analyse UK Biobank sleep data are encouraged to review our criteria, including cleaning pipeline (see Appendix 4), and employ scoring approaches consistent with their study-specific objectives.

Disordered sleep and poor sleep quality were common within UK Biobank participants who completed the detailed sleep questionnaire. These data, including proposed phenotype definitions, can now be integrated with a range of biomedical information to advance understanding of sleep.

## Data availability

The access to the UK Biobank can be requested on the UK Biobank website. The suggested phenotypes will be returned to the UK Biobank showcase after the acceptance of this manuscript. Researchers interested in obtaining early access to the phenotypes can do so using our codebase directly on the UK Biobank Research Analysis Platform.

## Acknowledgment

We would like to thank the UK Biobank team (in particular Kelly Davies, Fenella Starkey, Jill Rodd, Sarah Ewart, Christopher Stibbards, Howard Callen and Janet Maccora) and the participants for their contribution to this study. This research was conducted under UK Biobank project 6818.

## Footnotes

1 Pubmed search on 12th September 2025 (“uk biobank”[Title/Abstract] AND “circadian”[Title/Abstract]) OR (“uk biobank”[Title/Abstract] AND “sleep”[Title/Abstract])

