## Appendix for "Sleep and circadian health in the UK Biobank: Report on the 2023 sleep questionnaire enhancement"

#### Supplementary Information

**Appendix 1:** Rationale and procedure for administration of the sleep web-based questionnaire for UK Biobank

**Appendix 2:** Questions with multiple versions

**Appendix 3:** Phenotype coding

**Appendix 4:** Dealing with missing and uncertain responses

**Appendix 5:** Dealing with time-related issues

**Appendix 6**: Data fields for all single-item variables

**Appendix 7:** STROBE guideline tables

**Appendix 8:** eTables

**Appendix 1:** Rationale and procedure for administration of the sleep web-based questionnaire for UK Biobank

### UK Biobank

### Sleep web-based questionnaire

Version 1.1

http://www.ukbiobank.ac.uk/

7th May 2024

Enabling scientific discoveries that improve human health

This document details the rationale and procedure for administration of the sleep web-based questionnaire for UK Biobank.

#### Contents

1. Introduction – scientific rationale ... 4
2. List of contributors ... 5
3. Content ... 6
4. Piloting ... 9
5. Administration ... 9
6. References ... 10
Appendix 1: Questions and format of the questionnaire ... 13

### 1. Introduction – scientific rationale

Good quality, restorative sleep is central to health. Laboratory manipulations of sleep show acute adverse behavioural and physiological consequences[1], and prospective studies demonstrate that sleep and circadian rhythm disruption increases risk for a range of diseases, including psychiatric disorders, cancer, and type 2 diabetes[2]. Sleep disorders affect approximately 25% of the population and can substantially impair quality of life. Disordered sleep may reflect altered sleep duration, quality, or timing, as well as abnormal behaviours that emerge during sleep, in association with daytime dysfunction (including excessive somnolence). A further ~20% of the population experience insufficient sleep, often engendered by curtailment of sleep time due to work schedule or prioritisation of social and leisure activities. Both categories of sleep disruption represent significant challenges to health and well-being, and have been estimated to cost the UK economy >£40 billion per year[3]. Understanding the aetiology and consequences of sleep disruption has the potential to transform sleep therapeutics and inform public health guidance and intervention.

**Potential benefits of enhanced sleep measurement in the UK Biobank**

Current measurement of sleep and sleep disorders in UK Biobank is limited to single-item measures of insomnia, sleep duration, snoring, daytime sleepiness, and chronotype, or diagnostic codes extracted from primary care records. Sleep disorders are often under-recognised in clinical practice, and therefore codes from primary care records fail to capture the true magnitude and burden of disordered sleep[4]. Wrist-worn accelerometry has been collected in a sub-sample of UKB participants, which is a major strength, enabling assessment of circadian rest-activity rhythms as well as estimates of sleep timing, duration, and continuity[5, 6]. Over the last decade, a significant number of studies have been published using sleep and circadian-related data generated from UKB, including seminal GWAS of insomnia symptoms[7, 8], sleep duration[9] and chronotype[10].

While the sleep field recognises the uniqueness of the UK Biobank resource, it is clear that more fine-grained measurement of sleep and its disorders will help harness the full range of available biomedical data to drive novel discoveries[11]. For example, enhanced phenotyping will more precisely characterise heterogeneity within disorder categories and reduce misclassification between related sleep disorders (e.g. insomnia and restless legs syndrome). Because sleep disorders often go undiagnosed in primary care, we need good measures that capture probable cases to permit comparison with controls on genetics, biomarkers, structural and functional brain health, and other disease indices. Doing so will provide an unrivalled opportunity to define the underpinning biology and putative consequences of a range of sleep disorders, while accounting for the influence of other sleep (and non-sleep) confounding variables. There are no existing large-scale studies that do this in a comprehensive manner. Beyond disordered sleep, we also want to describe variation in normal sleep patterns and association with environmental and lifestyle factors, as well as change over time. For example, better characterisation of changes in sleep over time will help researchers interrogate associations between sleep and cognitive decline, enriched by concurrent cognitive testing and neuroimaging data. Middle-to-late adulthood is also a period characterised by a marked increase in the prevalence of several sleep disorders (e.g., insomnia, OSA, REM sleep behaviour disorder). More precise sleep phenotyping through questionnaire methods, in combination with actigraphic data, has the potential to identify novel biomarkers of sleep disorders and permit examination of predictive relationships with subsequent disease and mortality.

### 2. List of contributors

The following academics and clinicians provided input and advice on the content and structure of the sleep questionnaire:

**Questionnaire development lead:**

- Simon Kyle, Professor of Experimental and Clinical Sleep Research, Nuffield Department of Clinical Neurosciences, University of Oxford

**Other main expert contributors:**

- Colin Espie, Professor of Sleep Medicine, Nuffield Department of Clinical Neurosciences, University of Oxford

- Emmanuel Mignot, Professor of Psychiatry and Behavioral Sciences, Department of Psychiatry and Behavioral Sciences, Stanford University
- Derk-Jan Dijk, Professor of Sleep and Physiology, Surrey Sleep Research Centre, University of Surrey

### 3. Content

The UK Biobank sleep questionnaire has been designed to capture key domains of self-reported sleep health and sleep disorders using validated questionnaires and bespoke items where appropriate. Potential causes of sleep disruption are probed using attributional questions from the World Sleep Survey and Pittsburgh Sleep Quality Index, and through assessment of other key lifestyle (e.g. shift-work, exercise, alcohol, caffeine) and environmental (e.g. light, noise) variables. The use of wearable devices (to monitor sleep) and family history of sleep disorders are also probed. Chronotype is assessed with the reduced version of the Morning-Eveningness Questionnaire, supplemented by questions on sleep timing during work vs non-work days (where relevant).

There is no existing questionnaire with adequate psychometric properties that permits identification of all the main sleep disorder types. We therefore selected published scales commonly used in clinical research to assess common sleep disorders, supplemented by amended and additional questionnaire items. These include the Sleep Condition Indicator (insomnia), Berlin questionnaire (obstructive sleep apnoea), Cambridge-Hopkins questionnaire (restless legs syndrome), Brief Screen for Sleep Disorders (questions assessing delayed and advanced sleep-wake phase disorder) and Shift Work Disorder Screening Questionnaire (shift work sleep disorder), Alliance Sleep Questionnaire (narcolepsy, parasomnias) and single-item screen for REM sleep behaviour disorder (RBD1Q).

Sleep before the pandemic was assessed using bespoke questionnaire items to enable comparison with present day. We also assessed potential correlates of sleep and sleep disorders, including sleepiness (bespoke questionnaire items), fatigue (Flinders Fatigue Scale), depression and anxiety (PHQ-4), cognitive impairment (BC-CCI), and accidents

(Alliance questionnaire). We have selected brief measures where possible and have incorporated questionnaire branching where appropriate to tailor relevance. Our sleep questionnaire measures are compatible with other large-scale population-based studies (e.g., The Rotterdam Study[12], The HUNT-study[13]) and sleep-lab based studies (e.g., STAGES; The Stanford Technology Analytics and Genomics in Sleep study[14]), potentially facilitating comparisons and data pooling.

Some of the items in the sleep questionnaire have been asked of UK Biobank participants in previous questionnaires, and these are identified in Appendix 1. A detailed guide to the sources of all questions used in the questionnaire is included below.

| Domain | Source/tool | Notes about source/tool | Reference |
|---|---|---|---|
| rMEQ | Reduced MEQ | Assesses chronotype i.e. the extent to which one is morning or evening type. | Adan, A. & Almirall, H. (1991)[15] |
| Work and sleep | Shift work – bespoke | Screening question | N/A |
| | Shift work sleep disorder questionnaire adapted with permission of author | Screens for high risk of Shift Work Disorder (SWD) in a shift-working population. | Barger, L. K. *et al.* (2012)[16] |
| | Typical sleep times on work and non-work days - bespoke | Ascertains typical waking and sleeping times on working and non-working days. | N/A |
| Quality of sleep | Pittsburgh Sleep Quality Index (PSQI), adapted with permission of author | Assesses sleep quality over a 1-month time interval. | Buysse, D. J. *et al.* (1989)[17] |
| | Amended World Sleep Survey environmental influences on sleep questions | Assesses attributed causes of sleep disruption. | Espie et al. unpublished. |
| | Change to sleep patterns and sleep quality since the Covid-19 pandemic – bespoke | Assesses the impact that the pandemic has had on sleep patterns. | N/A |
| Sleep Condition Indicator | Sleep Condition Indicator, amended with permission of the author | Measures sleep problems against the DSM-5 criteria for insomnia disorder. | Espie, C. A. *et al.* (2014)[18] |
| | Wake up earlier than intended – bespoke | None | N/A |
| Sleep disorders | BAP (circadian rhythm disorders) | Screens for probable delayed and advanced sleep-wake phase disorder. | Wilson, S. *et al.* (2010)[19] |

| | | | |
|---|---|---|---|
| | Questions based on Alliance Sleep Questionnaire - narcolepsy and parasomnia items | The Alliance Sleep Questionnaire (ASQ) is a comprehensive, online sleep questionnaire that assesses sleep symptoms. It is administered to ~8,000 patients annually in the Stanford Sleep Medicine Centre. Note that all items are not included in this questionnaire so comparability cannot be assumed. | Leary, E.B. *et al.* (2014)[20] |
| | REM Sleep Behaviour Disorder Single-Question Screen | A screening question for dream enactment with a simple yes/no response. | Postuma, R.B. et al (2012)[21] |
| | Berlin Questionnaire: Sleep Apnoea | Validated questionnaire used to identify the risk (low to high) of sleep disordered breathing (obstructive sleep apnoea). Has been used in primary care and non-primary care settings. | Netzer, N. *et al. (*1999)[22] |
| | Alliance Sleep Questionnaire accident or near miss question | As above | As above |
| Fatigue | Flinders Fatigue Scale | Measures daytime fatigue (differentiated from sleepiness) | Gradisar, M. *et al.* (2007)[23] |
| Restless legs syndrome (RLS) | Questions based on CH-RLSq (Cambridge-Hopkins Restless Legs Syndrome Questionnaire), amended with permission of the authors | CH-RLSq is a well-validated questionnaire for identification of subjects likely to be diagnosed with RLS. Note that all items are not included in this questionnaire so comparability cannot be assumed. | Burchell, B. *et al.* (2008)[24] |
| Sleep consequences | Bespoke questions | Assesses self-perceived sleepiness, sleepiness in passive situation and sleepiness in active situation. This new scale will be validated against the Epworth Sleepiness Scale which is administered to ~8,000 patients annually in the Stanford Sleep Medicine Centre. | N/A |
| | British Columbia Cognitive Complaints Inventory | Assesses perceived cognitive difficulties | Iverson, G.L. & Lam R.W. (2013)[25] |
| Family history | Alliance Sleep Questionnaire Family History | As above | As above |
| Lifestyle and behaviours | Key lifestyle and variables (sleep tracker, travel, alcohol, exercise, use of electronic devices, time outside, caffeine) – bespoke | None | N/A |
| | Alliance Sleep Questionnaire | As above | As above |
| | Questions about hours spent outdoors, | None | None |

| | | | |
|---|---|---|---|
| | | previously used in UK Biobank questionnaire – bespoke | |
| PHQ-4 | Patient Health Questionnaire-4 | An established research and clinical tool. All or part of the scale has previously been completed by UK Biobank participants at several time points. | Kroenke, K. *et al.* (2009)[26] |

The full list of questions can be found in Appendix 1.

### 4. Piloting

Prior to inviting all participants with a contact email address (approximately 330,000) to complete it, UK Biobank piloted this questionnaire with ~13,000 participants to ensure that the online platform and procedures were adequately robust and that the questionnaire was acceptable in terms of content and length. Several questions and portions of text in the 'work and sleep, and 'sleep consequences' modules were found to need revision during the pilot. Data captured using the different question wording is presented in different data fields. Examples of this are WaS1 and WaS1_ALT.

### 5. Administration

**5.1:** The questionnaire administration process for UK Biobank participants with an email address was as follows:

- an initial invitation email (which included a hyperlink to their personalised questionnaire);
- a reminder email to non-responders sent two weeks after the initial invitation;
- a reminder email sent to partial responders (i.e. those who had only completed part of the questionnaire) two weeks after they started the questionnaire;
- a 'last-chance' invitation sent to non-responders four months after the initial invitation.

**5.2:** Participants for whom UK Biobank did not have an email address were encouraged via the information on the UK Biobank website to complete the online questionnaire by logging on directly to the participant website.

**5.3:** The median time for participants to fully complete the questionnaire was 22.6 minutes. Excluding the top 5 per cent of completion times (which were outliers resulting from there being no timeout on the questionnaire modules), 77 per cent of participants completed the questionnaire in 30 minutes or less.

**5.4:** Data were removed from participants who failed the identity check at the beginning of the online questionnaire: that is, the dates of birth they provided did not match UK Biobank records. These amounted to 0.22% of all respondents.

**5.5:** Researchers are advised to review the distributions of fields that contain data from self-reported measures before using them for analyses. Extreme values may exist in the data,

and no attempt to verify the accuracy of responses has been made.

**5.6:** Email invitations are also routinely sent to those participants who have recently updated their email address (and who have not yet completed the questionnaire). We therefore anticipate that data will continue to accrue for a small number of participants.

### Appendix 1: Questions and format of the questionnaire

**Sleep questionnaire (v3.6, 29th February 2024)**

**Introduction**

Good quality, restorative sleep is central to health. Understanding the biological and environmental factors that lead to poor sleep as well as the consequences of sleep disruption has the potential to help millions of people have a better night's sleep.

UK Biobank provides a unique opportunity to gather comprehensive information about sleep. We hope to combine the results of this questionnaire with other information you have provided, to gain a deeper understanding of the genetic determinants of sleep and its relationship with health and disease.

| Q.ID | Field I.D | Stem | Question identical to previous questionnaire | Question similar to previous questionnaire | Responses |
|---|---|---|---|---|---|
| **Identity check** | | | | | |
| **ID_INTRO 1** | | This questionnaire is participant specific. It should only be completed by the person named on the email invitation OR the person who logged in to the participant website.<br><br>We just want to check your date of birth. This is so that we can double-check that this questionnaire has been completed by the correct person (and not, for example, by someone who shares an email address with you). | | | |
| **ID_INTRO 2** | | Please enter your details below: | | | |
| **ID_DAY** | | Day of birth: | | | *[DropdownList1: 31 choices 1 – 31.]* |
| **ID_MONTH** | | Month of birth: | | | *[DropdownList2: 12 choices for months: "January" to "December".]* |
| **ID_YEAR** | | Year of birth: | | | *[Text box allowing integer values and it allows selection of an integer between 1934 and 1971.]* |
| **Sleep behaviour** | | | | | |
| **MEQINTRO** | | We would like to ask you some questions about your sleep patterns. | | | |
| **MEQ1** | 30425 | Approximately what time would you get up if you were entirely free to plan your day? | | | *[Select one from]*<br>- 01=5:00–6:30am<br>- 02=6:30–7:45am<br>- 03=7:45–9:45am<br>- 04=9:45–11:00am<br>- 05=11:00–12 noon<br>- 06=Other time of the day<br>- DA=Prefer not to answer |
| **MEQ2** | 30426 | During the first half hour after you wake up in the morning, how do you usually feel? | | | *[Select one from]*<br>- 01=Very tired<br>- 02=Fairly tired<br>- 03=Fairly refreshed |

| | | | | | |
|---|---|---|---|---|---|
| | | | | | - 04=Very refreshed<br>- DA=Prefer not to answer |
| **MEQ3** | 3042<br>7 | At approximately what time in the evening do you usually feel tired, and, as a result, in need of sleep? | | | *[Select one from]*<br>- 01=8:00–9:00pm<br>- 02=9:00–10:15pm<br>- 03=10:15 pm–12:45am<br>- 04=12:45–2:00am<br>- 05=2:00–3:00am<br>- 06=Other time of the day<br>- DA=Prefer not to answer |
| **MEQ4** | 3042<br>8 | At approximately what time of day do you usually feel your best? | | | *[Select one from]*<br>- 01=5:00–8:00am<br>- 02=8:00–10:00am<br>- 03=10:00 am–5:00pm<br>- 04=5:00–10:00pm<br>- 05=10:00 PM–5:00am<br>- DA=Prefer not to answer |
| **MEQ5** | 3042<br>9 | One hears about “morning-types” and “evening-types.” Which one of these types do you consider yourself to be? | | Field ID:<br>1180<br>(Touchscr<br>een sleep<br>questions<br>) | *[Select one from]*<br>- 01=Definitely a morning-type<br>- 02=Rather more a morning-type than an evening-type<br>- 03=Rather more an evening-type than a morning-type<br>- 04=Definitely an evening-type<br>- DK=Do not know<br>- DA=Prefer not to answer |
| **Work and sleep** | | | | | |
| **WASIN<br>TRO** | | This module is about your work patterns. | | | |
| **WASIN<br>TRO_A<br>LT** | | This module is about your work patterns. Please complete this module even if you are not currently working, for example if you are retired. | | | |
| **WaS1** | 3043<br>0 | In the past month, did you typically work a non-standard shift schedule?<br><br>*Here we define non-standard shift schedule as working hours outside the hours of 8am to 8pm.* | | | *[Select one from]*<br>- 01=No – I didn’t work<br>- 02=No – I worked standard hours (e.g. 9:00am-5:00pm)<br>- 03=Yes – I worked evening shifts (work typically finished between 8:00pm and midnight)<br>- 04=Yes - I worked morning shifts (work typically started between 4:00am and 7:00am)<br>- 05=Yes – I worked night shifts (work typically took place between 8:00pm and 8:00am)<br>- 06=Yes – I worked rotating shifts or had irregular work hours<br>DA=Prefer not to answer |
| **WaS1_<br>ALT** | 3043<br>1 | In the past month, did you typically work a non-standard shift schedule?<br><br>*Here we define non-standard shift schedule as working hours outside the hours of 8am to 8pm.* | | | *[Select one from]*<br>- 01=No – I didn’t work<br>- 02=No – I worked standard hours (e.g. 9:00am-5:00pm)<br>- 03=Yes – I worked evening shifts (work typically finished between 8:00pm and midnight)<br>- 04=Yes - I worked morning shifts (work typically started between 4:00am and 7:00am)<br>- 05=Yes – I worked night shifts (work typically took place between 8:00pm and 8:00am) |

| | | | | | |
|---|---|---|---|---|---|
| | | *If you are retired, please answer "No, I didn't work".* | | | - 06=Yes – I worked rotating shifts or had irregular work hours<br>DA=Prefer not to answer |
| **WaS2** | 30432 | In the past month, **while working non-standard shifts**, did you have a problem *with waking up too early and not being able to get back to sleep?* | | | *[Select one from]*<br>- 01=No<br>- 02=Yes, a minor problem<br>- 03=Yes, a considerable problem<br>- 04=Yes, a serious problem<br>- DA=Prefer not to answer |
| **WaS3** | 30433 | In the past month, **while working non-standard shifts**, was your *sense of well-being:* | | | *[Select one from]*<br>- 01=very good?<br>- 02=fairly good?<br>- 03=fairly bad?<br>- 04=very bad?<br>- DA=Prefer not to answer |
| **WaS4** | 30434 | In the past month, **during your non-standard shift**, how likely were you *to doze off at work?* | | | *[Select one from]*<br>- 01=Not likely at all<br>- 02=Slightly likely<br>- 03=Moderately likely<br>- 04=Highly likely<br>- DA=Prefer not to answer |
| **WaS5** | 30435 | In the past month, how likely were you *to doze off or fall asleep* while driving **after at least two days off from work?** | | | *[Select one from]*<br>- 01=Not likely at all<br>- 02=Slightly likely<br>- 03=Moderately likely<br>- 04=Highly likely<br>- DA=Prefer not to answer |
| **WaS6** | 30436 | On workdays in the past month, what time did you typically fall asleep? (This may be different to the time you went to bed) | | | - Menu1 Hours from 9:00pm to 8:00pm in hour increments<br>- Menu2 Minutes from 00 to 55 in 5 minute increments<br>OR<br>- V=Varies significantly<br>- DA=Prefer not to answer |
| **WaS7** | 30437 | On workdays in the past month, what time did you typically wake-up? (By 'wake up' we mean your final awakening time before getting up for the day) | | | - Menu3 Hours from 5:00am to 4:00am in hour increments<br>- Menu4 Minutes from 00 to 55 in 5 minute increments<br>OR<br>- V=Varies significantly<br>- DA=Prefer not to answer |
| **WaS8** | 30438 | On non-working days in the past month, what time did you typically fall asleep?<br>(This may be different to the time you went to bed)" | | | - Menu5 Hours from 9:00pm to 8:00pm in hour increments<br>- Menu6 Minutes from 00 to 55 in 5 minute increments<br>OR<br>- V=Varies significantly<br>- DA=Prefer not to answer |
| **WaS8_ALT** | 30439 | On non-working days in the past month, what time did you typically fall asleep? (This may be different from the time you went to bed).<br><br>By non-working days, we mean any days that you did not work, including if you are retired. | | | - Menu5 Hours from 9:00pm to 8:00pm in hour increments<br>- Menu6 Minutes from 00 to 55 in 5 minute increments<br>OR<br>- V=Varies significantly<br>- DA=Prefer not to answer |

| | | | | | |
|---|---|---|---|---|---|
| **WaS9** | 30440 | On non-working days in the past month, what time did you typically wake up?<br>(By 'wake up' we mean your final awakening time before getting up for the day) | | | - Menu7 Hours from 5:00am to 4:00am in hour increments<br>- Menu8 Minutes from 00 to 55 in 5 minute increments<br>OR<br>- V=Varies significantly<br>- DA=Prefer not to answer |
| **WaS9_ ALT** | 30441 | On non-working days in the past month, what time did you typically wake up?<br>(By 'wake up' we mean your final awakening time before getting up for the day)<br>By non-working days, we mean any days that you did not work, including if you are retired. | | | - Menu7 Hours from 5:00am to 4:00am in hour increments<br>- Menu8 Minutes from 00 to 55 in 5 minute increments<br>OR<br>- V=Varies significantly<br>- DA=Prefer not to answer- 00 No<br>- DA Prefer not to answer |
| **Quality of sleep** | | | | | |
| **Quality INTRO** | | You may notice some overlap between questions in this section and other sections – this is so that we can ensure that we collect accurate information from you. We would be grateful if you could complete all questions even if you think you have answered them already. | | | |
| **Quality 1INTRO** | | The following questions relate to your usual sleep habits during the **past month only**. Your answers should indicate the most accurate reply for the **majority** of days and nights in the past month.<br>By "night" we mean the time period you expect to be sleeping, and by "morning" we mean the time you expect to wake/get up.<br><br>*[Copyright notices are required for questions Quality1-Quality5i, Quality5p, Quality 7, Quality 8 and Quality9-Quality11.*<br><br>*Notice is: "Copyright 1989 and 2010. University of Pittsburgh. All rights reserved."* | | | |

| | | | | | |
|---|---|---|---|---|---|
| **Quality 1** | 3044 2 | During the past month, what time have you usually gone to bed at night? | | | *[Menu1 only allowing one selection, of an hour between 9pm and 8pm. - Menu2 only allowing one selection, of minutes from 00 to 55 in 5-minute increments. Prefixed “Bed time”. If Quality1=Prefer not to answer (DA), no other selection can be made.]*<br><br>- Menu1 [Hours from 9pm to 8pm. Display “12 midnight” instead of “12am” and “12 noon” instead of “12pm”.]<br>- Menu2 [Minutes from 00 to 55 in 5-minute increments]<br>OR<br>- DA = Prefer not to answer |
| **Quality 2** | 3044 3 | During the past month, how long has it usually taken you to fall asleep each night? | | | *[Menu3 only allowing one selection, of an integer between 0 and 8. Menu3 is suffixed “hour(s)”.*<br>*Menu4 only allowing one selection, of minutes from 00 to 55 in 5-minute increments. Menu4 is suffixed “minutes”. If Quality2=Prefer not to answer (DA), no other selection can be made*.]<br><br>- Menu3 [Hours from 0 to 8]<br>- Menu4 [Minutes from 00 to 55 in 5-minute increments]<br>OR<br>- DA = Prefer not to answe |
| **Quality 3** | 3044 4 | During the past month, what time have you usually got up in the morning? | | | *[Menu5 only allowing one selection, of an hour between 5am and 4am. Menu6 only allowing one selection, of minutes from 00 to 55 in 5-minute increments. Prefixed “Getting up time”. If Quality3=Prefer not to answer (DA), no other selection can be made.]*<br><br>- Menu5 [Hours from 5am to 4am. Display “12 midnight” instead of “12am” and “12 noon” instead of “12pm”.]<br>- Menu6 [Minutes from 00 to 55 in 5-minute increments]<br>OR<br>- DA = Prefer not to answer |
| **Quality 4** | 3044 5 | During the past month, how many hours of **actual sleep** did you get each night?<br><br>(This may be different to the number of hours you spent in bed.) | | | *[Menu7 only allowing one selection, of an integer between 0 and 20. Menu7 is suffixed “hour(s)”.*<br>*Menu8 only allowing one selection, of minutes from 00 to 55 in 5-minute increments.*<br>*Menu8 is suffixed “minutes”. If Quality4=Prefer not to answer (DA), no other selection can be made*.]<br><br>- Menu7 [Hours from 0 to 20]<br>- Menu8 [Minutes from 00 to 55 in 5-minute increments]<br>OR<br>- DA = Prefer not to answer |
| **Quality 5INTRO** | | For each of the next questions, select the one best response. Please answer all questions.<br><br>By “night” we mean the time period you expect | | | |

| | | | | | |
|---|---|---|---|---|---|
| | | to be sleeping, and by "morning" we mean the time you expect to wake/get up. | | | |
| **BLOCK Quality 5** | | **During the past month**, how often have you had trouble sleeping because: | | | |
| **Quality 5a** | 3044 6 | you cannot get to sleep within 30 minutes? | | | [*Select one from*]<br>- 00=Not during the past month<br>- 01=Less than once a week<br>- 02=Once or twice a week<br>- 03=Three or more times a week<br>- DA=Prefer not to answer |
| **Quality 5b** | 3044 7 | you wake up in the middle of the night or early morning? | | | [*Select one from*]<br>- 00=Not during the past month<br>- 01=Less than once a week<br>- 02=Once or twice a week<br>- 03=Three or more times a week<br>- DA=Prefer not to answer |
| **Quality 5c** | 3044 8 | you have to get up to use the bathroom? | | | [*Select one from*]<br>- 00=Not during the past month<br>- 01=Less than once a week<br>- 02=Once or twice a week<br>- 03=Three or more times a week<br>- DA=Prefer not to answer |
| **Quality 5d** | 3044 9 | you cannot breathe comfortably? | | | [*Select one from*]<br>- 00=Not during the past month<br>- 01=Less than once a week<br>- 02=Once or twice a week<br>- 03=Three or more times a week<br>- DA=Prefer not to answer |
| **Quality 5e** | 3045 0 | you cough or snore loudly? | | | [*Select one from*]<br>- 00=Not during the past month<br>- 01=Less than once a week<br>- 02=Once or twice a week<br>- 03=Three or more times a week<br>- DA=Prefer not to answer |
| **Quality 5f** | 3045 1 | you feel too cold? | | | [*Select one from*]<br>- 00=Not during the past month<br>- 01=Less than once a week<br>- 02=Once or twice a week<br>- 03=Three or more times a week<br>- DA=Prefer not to answer |
| **Quality 5g** | 3045 2 | you feel too hot? | | | [*Select one from*]<br>- 00=Not during the past month<br>- 01=Less than once a week<br>- 02=Once or twice a week<br>- 03=Three or more times a week<br>- DA=Prefer not to answer |
| **Quality 5h** | 3045 3 | you have bad dreams? | | | [*Select one from*]<br>- 00=Not during the past month<br>- 01=Less than once a week<br>- 02=Once or twice a week<br>- 03=Three or more times a week<br>- DA=Prefer not to answer |
| **Quality 5i** | 3045 4 | you have pain? | | | [*Select one from*]<br>- 00=Not during the past month<br>- 01=Less than once a week<br>- 02=Once or twice a week<br>- 03=Three or more times a week<br>- DA=Prefer not to answer |

| | | | | | |
|---|---|---|---|---|---|
| **Quality 5j** | 30455 | you find it noisy? | | | [*Select one from*]<br>- 00=Not during the past month<br>- 01=Less than once a week<br>- 02=Once or twice a week<br>- 03=Three or more times a week<br>- DA=Prefer not to answer |
| **Quality 5k** | 30456 | you find the bed uncomfortable? | | | [*Select one from*]<br>- 00=Not during the past month<br>- 01=Less than once a week<br>- 02=Once or twice a week<br>- 03=Three or more times a week<br>- DA=Prefer not to answer |
| **Quality 5l** | 30457 | you are disturbed by the light levels? | | | [*Select one from*]<br>- 00=Not during the past month<br>- 01=Less than once a week<br>- 02=Once or twice a week<br>- 03=Three or more times a week<br>- DA=Prefer not to answer |
| **Quality 5m** | 30458 | you are worried about something? | | | [*Select one from*]<br>- 00=Not during the past month<br>- 01=Less than once a week<br>- 02=Once or twice a week<br>- 03=Three or more times a week<br>- DA=Prefer not to answer |
| **Quality 5n** | 30459 | you are disturbed by children? | | | [*Select one from*]<br>- 00=Not during the past month<br>- 01=Less than once a week<br>- 02=Once or twice a week<br>- 03=Three or more times a week<br>- DA=Prefer not to answer |
| **Quality 5o** | 30460 | you are disturbed by your bed partner? | | | [*Select one from*]<br>- 00=Not during the past month<br>- 01=Less than once a week<br>- 02=Once or twice a week<br>- 03=Three or more times a week<br>- DA=Prefer not to answer |
| **Quality 5p** | 30461 | of some other reason(s)? | | | [*Select one from*]<br>- 00=Not during the past month<br>- 01=Less than once a week<br>- 02=Once or twice a week<br>- 03=Three or more times a week<br>- DA=Prefer not to answer |
| **Quality 6** | 30462 | Thinking about a typical night in the past month, how many times did you wake up **during** the night? | | | *[Select one from]*<br>- 00=0 times<br>- 01=1-2 times<br>- 02=3-4 times<br>- 03=5-6 times<br>- 04=7-8 times<br>- 05=9-10 times<br>- 06=More than 10 times<br>- DA=Prefer not to answer |
| **Quality 7** | 30463 | During the past month, how often have you taken over the counter medicine (non-prescription) to help you sleep? | | | *[Select one from]*<br>- 00=Not during the past month<br>- 01=Less than once a week<br>- 02=Once or twice a week<br>- 03=Three or more times a week<br>- DA=Prefer not to answer |
| **Quality 8** | 30464 | During the past month, how often have you taken prescription medicine to help you sleep? | | | *[Select one from]*<br>- 00=Not during the past month<br>- 01=Less than once a week<br>- 02=Once or twice a week<br>- 03=Three or more times a week<br>- DA=Prefer not to answer |

| | | | | | |
|---|---|---|---|---|---|
| **Quality 9** | 30465 | During the past month, how often have you had trouble staying awake while driving, eating meals or engaging in social activity? | | | *[Select one from]*<br>- 00=Not during the past month<br>- 01=Less than once a week<br>- 02=Once or twice a week<br>- 03=Three or more times a week<br>- DA=Prefer not to answer |
| **Quality 10** | 30466 | During the past month, how much of a problem has it been for you to keep up enough enthusiasm to get things done? | | | *[Select one from]*<br>- 00=No problem at all<br>- 01=Only a slight problem<br>- 02=Somewhat of a problem<br>- 03=A very big problem<br>- DA=Prefer not to answer |
| **Quality 11** | 30467 | During the past month, how would you rate your sleep quality overall? | | | *[Select one from]*<br>- 01=Very good<br>- 02=Fairly good<br>- 03=Fairly bad<br>- 04=Very bad<br>- DA=Prefer not to answer |
| **Quality 12INTRO** | | For each of the next questions, we would like you to think about your sleep quality in 2019, i.e. before the COVID-19 pandemic began. | | | |
| **Quality 12** | 30468 | In 2019 (i.e. before the pandemic), typically how many nights a week did you have a problem with your sleep (e.g. issues falling asleep, waking in the night, waking before you intended to)? | | | *[Select one from]*<br>- 00=Never<br>- 01=Less than once a week<br>- 02=Once or twice a week<br>- 03=Three or more times a week<br>- DK=Do not know/remember<br>- DA=Prefer not to answer |
| **Quality 13** | 30469 | Compared to now, how likely were you in 2019 (i.e. before the pandemic) to fall asleep when you didn't intend to? | | | *[Select one from]*<br>- 00=Not likely at all<br>- 01=Slightly likely<br>- 02=Moderately likely<br>- 03=Highly likely<br>- DK=Do not know/remember<br>- DA=Prefer not to answer |
| **Quality 14** | 30470 | Thinking about a typical night in 2019 (i.e. before the pandemic), how would you have rated your sleep quality?<br>(By "night", we mean the time period you expect to be sleeping.) | | | *[Select one from]*<br>- 01=Very good<br>- 02=Fairly good<br>- 03=Fairly bad<br>- 04=Very bad<br>- DK=Do not know/remember<br>- DA=Prefer not to answer |
| **Quality 15** | 30471 | Has your usual number of hours of sleep per night changed since 2019 i.e. since before the pandemic?<br>(By "a little bit", we mean less than an hour and by "a lot", we mean more than an hour. By "night", we mean the time period you expect to be sleeping.) | | | *[Select one from]*<br>- 01= Yes, I sleep a lot more now compared to before the pandemic<br>- 02= Yes, I sleep a little bit more now compared to before the pandemic<br>- 00= No, the amount I sleep has not changed<br>- 03=Yes, I sleep a little bit less now compared to before the pandemic<br>- 04=Yes, I sleep a lot less now compared to before the pandemic<br>- DK=Do not know/remember<br>- V=It varies significantly<br>- DA=Prefer not to answer |

| | | | | | |
|---|---|---|---|---|---|
| **Quality 15a** | 30472 | In 2019 (i.e. before the pandemic), how many hours of **actual sleep** did you get each night? (This may be different to the number of hours you spent in bed.) | | | *[Menu9 only allowing one selection, of an integer between 0 and 20. Menu9 is suffixed "hour(s)" Menu10 only allowing one selection, of minutes from 00 to 55 in 5-minute increments. Menu10 is suffixed "minutes". If Quality15a=Prefer not to answer (DA), no other selection can be made.]*<br><br>- Menu9 [Hours from 0 to 20]<br>- Menu10 [Minutes from 00 to 55 in 5-minute increments]<br>OR<br>- DA = Prefer not to answer |
| **Quality 15b** | 30473 | Please tell us why. (Select all that apply) | | | *[Select one or more from 01-07. DK and DA are exclusive. If Quality15b=Do not know (DK) or Prefer not to answer (DA), no other selection can be made]*<br><br>- 01=I have retired<br>- 02=I have changed my job<br>- 03=I have changed my working hours e.g. gone part-time or begun working shifts<br>- 04=I now work from home so I no longer need to commute<br>- 05=I have/had physical health issues<br>- 06=I have/had mental health issues<br>- 07=Other reason(s)<br>- DK=Do not know<br>- DA=Prefer not to answer |
| **Insomnia** | | | | | |
| **SCI-INTRO** | | We would like to know more about your quality of sleep. By "night" we mean the time period you expect to be sleeping, and by "morning" we mean the time you expect to wake/get up. | | | |
| **BLOCK SCI1** | | Thinking about a typical night in the **past month**… | | | [Select one from the following for each of the statements] |
| **SCI1a** | 30474 | ... how long does it take you to fall asleep? | | | [*Select one from*]<br>- 01=0-15 mins<br>- 02=16-30 mins<br>- 03=31-45 mins<br>- 04=46-60 mins<br>- 05=61 mins or more<br>- DA=Prefer not to answer |
| **SCI1b** | 30536 | If you then wake up during the night, how long are you awake for in total? (add all the wakenings up) | | | *[Select one from]*<br>- 01=0-15 mins<br>- 02=16-30 mins<br>- 03=31-45 mins<br>- 04=46-60 mins<br>- 05=61 mins or more<br>- DA=Prefer not to answer |
| **SCI1c** | 30537 | ... how long before you *intend* to wake up do you actually wake up? (e.g. if you want to wake up at 7am but wake up at 6:15am most | | | *[Select one from]*<br>- 01=I don't wake up too early<br>- 02=Up to 15 mins early<br>- 03=16-30 mins early<br>- 04=31-45 mins early<br>- 05=46-60 mins early |

| | | | | | |
|---|---|---|---|---|---|
| | | mornings, this would be a difference of 45 minutes.) | | | - 06=More than 60 mins early<br>- DA=Prefer not to answer |
| **SCI1d** | 30538 | ... how many nights a week do you have a problem with your sleep? | | | *[Select one from]*<br>- 01=0-1<br>- 02=2<br>- 03=3<br>- 04=4<br>- 05=5-7<br>- DA=Prefer not to answer |
| **SCI1e** | 30539 | ... how would you rate your sleep quality? | | | *[Select one from]*<br>- 01=Very good<br>- 02=Good<br>- 03=Average<br>- 04=Poor<br>- 05=Very poor<br>- DA=Prefer not to answer |
| **BLOCK SCI2** | | Thinking about the past month, to what extent has poor sleep... | | | |
| **SCI2a** | 30540 | ... affected your mood, energy or relationships? | | | *[Select one from]*<br>- 01=Not at all<br>- 02=A little<br>- 03=Somewhat<br>- 04=Much<br>- 05=Very much<br>- DA=Prefer not to answer |
| **SCI2b** | 30541 | ... affected your concentration, productivity or ability to stay awake? | | | *[Select one from]*<br>- 01=Not at all<br>- 02=A little<br>- 03=Somewhat<br>- 04=Much<br>- 05=Very much<br>- DA=Prefer not to answer |
| **SCI2c** | 30542 | ... troubled you in general? | | | *[Select one from]*<br>- 01=Not at all<br>- 02=A little<br>- 03=Somewhat<br>- 04=Much<br>- 05=Very much<br>- DA=Prefer not to answer |
| **SCI3** | 30543 | Finally....<br>... how long have you had a problem with your sleep? | | | *[Select one from]*<br>- 01=I don’t have a problem<br>- 02=Less than 1 month<br>- 03=1-2 months<br>- 04=3-6 months<br>- 05=7-12 months<br>- 06=1-5 years<br>- 07=6-10 years<br>- 08=More than 10 years<br>- DA=Prefer not to answer |
| **Sleep disturbances** | | | | | |
| **CRD-INTRO** | | We would like to know about any sleep problems and symptoms often associated with sleep problems that you might experience. | | | |
| **CRD1** | 30544 | Do you tend to sleep well but just at the “wrong times”? | | | *[Select one from]*<br>- 01=Yes<br>- 00=No |

| | | | | | |
|---|---|---|---|---|---|
| | | | | | - DA=Prefer not to answer |
| **CRD1a** | 3054 5 | Can you sleep well enough, but only if you stay up very late? | | | *[Select one from]*<br>- 01=Yes<br>- 00=No<br>- DA=Prefer not to answer |
| **CRD1b** | 3054 6 | Are you in a very sound sleep at normal waking time and could sleep on for hours more? | | | *[Select one from]*<br>- 01=Yes<br>- 00=No<br>- DA=Prefer not to answer |
| **CRD1c** | 3054 7 | Can you sleep well enough, but only if you go to bed very early? | | | *[Select one from]*<br>- 01=Yes<br>- 00=No<br>- DA=Prefer not to answer |
| **CRD1d** | 3054 8 | Do you wake very early, bright and alert and no longer sleepy? | | | *[Select one from]*<br>- 01=Yes<br>- 00=No<br>- DA=Prefer not to answer |
| **BLOCK Nar1** | | Do you currently experience any of these types of muscle weakness in the following situations? (please select all that apply) | | | |
| **Nar1a** | 3054 9 | Buckling of the knees? | | | *[Select one or more from 01-05. 06, 00 and DA are exclusive. If Nar1a=* I used to but not currently *(06) or I have never experienced this (00) or Prefer not to answer (DA), no other selection can be made]*<br>- 01=When I tell or hear a joke<br>- 02=When I laugh<br>- 03=When I am angry<br>- 04=When I am making a quick verbal response in a playful context<br>- 05=In a different situation<br>- 06=I used to but not currently<br>- 00=I have never experienced this<br>- DA=Prefer not to answer |
| **Nar1b** | 3055 0 | Sagging or dropping of your jaw? | | | *[Select one or more from 01-05. 06, 00 and DA are exclusive. If Nar1b= I used to but not currently (06) or I have never experienced this (00) or Prefer not to answer (DA), no other selection can be made]*<br>- 01=When I tell or hear a joke<br>- 02=When I laugh<br>- 03=When I am angry<br>- 04=When I am making a quick verbal response in a playful context<br>- 05=In a different situation<br>- 06=I used to but not currently<br>- 00=I have never experienced this<br>- DA=Prefer not to answer |
| **Nar1c** | 3055 1 | Abrupt dropping of your head and/or shoulders? | | | *[Select one or more from 01-05. 06, 00 and DA are exclusive. If Nar1c= I used to but not currently (06) or I have never experienced this (00) or Prefer not to answer (DA), no other selection can be made]*<br>- 01=When I tell or hear a joke<br>- 02=When I laugh<br>- 03=When I am angry<br>- 04=When I am making a quick verbal response in a playful context<br>- 05=In a different situation |

| | | | | | |
|---|---|---|---|---|---|
| | | | | | - 06=I used to but not currently<br>- 00=I have never experienced this<br>- DA=Prefer not to answer |
| **Nar1d** | 3055<br>2 | Weakness in your arms? | | | *[Select one or more from 01-05. 06, 00 and DA are exclusive. If Nar1d= I used to but not currently (06) or I have never experienced this (00) or Prefer not to answer (DA), no other selection can be made]*<br>- 01=When I tell or hear a joke<br>- 02=When I laugh<br>- 03=When I am angry<br>- 04=When I am making a quick verbal response in a playful context<br>- 05=In a different situation<br>- 06=I used to but not currently<br>- 00=I have never experienced this<br>- DA=Prefer not to answer |
| **Nar1e** | 3055<br>3 | Slurring of speech? | | | *[Select one or more from 01-05. 06, 00 and DA are exclusive. If Nar1e=* I used to but not currently *(06) or I have never experienced this (00) or Prefer not to answer (DA), no other selection can be made]*<br>- 01=When I tell or hear a joke<br>- 02=When I laugh<br>- 03=When I am angry<br>- 04=When I am making a quick verbal response in a playful context<br>- 05=In a different situation<br>- 06=I used to but not currently<br>- 00=I have never experienced this<br>- DA=Prefer not to answer |
| **Nar1f** | 3055<br>4 | Falling to the ground, unable to move? | | | *[Select one or more from 01-05. 06, 00 and DA are exclusive. If Nar1f= I used to but not currently (06) or I have never experienced this (00) or Prefer not to answer (DA), no other selection can be made]*<br>- 01=When I tell or hear a joke<br>- 02=When I laugh<br>- 03=When I am angry<br>- 04=When I am making a quick verbal response in a playful context<br>- 05=In a different situation<br>- 06=I used to but not currently<br>- 00=I have never experienced this<br>- DA=Prefer not to answer |
| **Para1** | 3055<br>5 | How often do you walk in your sleep? | | | *[Select one from]*<br>- 00=Never<br>- 01=Not in the past year<br>- 02=1-2 times per year<br>- 03=1-2 times per month<br>- 04=1-2 times per week<br>- 05=3-4 times per week<br>- 06=Almost every day<br>- 07=Every day<br>- DK=Do not know<br>- DA=Prefer not to answer |
| **Para2** | 3055<br>6 | Do you grind your teeth while you sleep? | | | *[Select one from]*<br>- 00=Never<br>- 01=Not in the past year<br>- 02=1-2 times per year<br>- 03=1-2 times per month<br>- 04=1-2 times per week<br>- 05=3-4 times per week |

| | | | | | |
|---|---|---|---|---|---|
| | | | | | - 06=Almost every day<br>- 07=Every day<br>- DK=Do not know<br>- DA=Prefer not to answer |
| **Para3** | 30557 | Have you ever been told, or suspected yourself, that you seem to ‘act out your dreams’ while asleep (for example, flailing your arms in the air, making running movements, etc.)? | | | *[Select one from]*<br>- 01=Yes<br>- 00=No<br>- DK=Do not know<br>- NA=Not applicable<br>- DA=Prefer not to answer |
| **Para4** | 30558 | How often do you have violent or injurious behaviour during sleep (for example, punching, kicking, leaping or running away from the bed)? | | | *[Select one from]*<br>- 00=Never<br>- 01=Not in the past year<br>- 02=1-2 times per year<br>- 03=1-2 times per month<br>- 04=1-2 times per week<br>- 05=3-4 times per week<br>- 06=Almost every day<br>- 07=Every day<br>- DK=Do not know<br>- DA=Prefer not to answer |
| **Para5** | 30559 | How often do you have nightmares (frightening dreams)? | | | *[Select one from]*<br>- 00=Never<br>- 01=Not in the past year<br>- 02=1-2 times per year<br>- 03=1-2 times per month<br>- 04=1-2 times per week<br>- 05=3-4 times per week<br>- 06=Almost every day<br>- 07=Every day<br>- DK=Do not know<br>- DA=Prefer not to answer |
| **Para5a** | 30560 | How often do you remember your dreams? | | | *[Select one from]*<br>- 00=Never<br>- 01=Not in the past year<br>- 02=1-2 times per year<br>- 03=1-2 times per month<br>- 04=1-2 times per week<br>- 05=3-4 times per week<br>- 06=Almost every day<br>- 07=Every day<br>- DK=Do not know<br>- DA=Prefer not to answer |
| **Para6** | 30561 | Do you have seizures, convulsions or “fits” during sleep? | | | *[Select one from]*<br>- 01=Yes<br>- 00=No<br>- DK=Do not know<br>- DA=Prefer not to answer |
| **ApnINTRO** | | Please choose the correct response to each question. | | | |
| **Apn1** | 30562 | Do you snore? | | | *[Select one from]*<br>01=Yes<br>00=No<br>DK=Do not know<br>DA=Prefer not to answer |
| **Apn1a** | 30563 | Your snoring is: | | | *[Select one from]*<br>01=Slightly louder than breathing<br>02=As loud as talking<br>03=Louder than talking |

| | | | | | |
|---|---|---|---|---|---|
| | | | | | DK=Do not know<br>DA=Prefer not to answer |
| **Apn1b** | 3056<br>4 | How often do you snore? | | | *[Select one from]*<br>- 01=Almost every day<br>- 02=3-4 times per week<br>- 03=1-2 times per week<br>- 04=1-2 times per month<br>- 05=Rarely or never<br>- DK=Do not know<br>- DA=Prefer not to answer |
| **Apn1c** | 3056<br>5 | Has your snoring ever bothered other people? | | Field ID:<br>1210<br>(Touchscr een sleep questions<br>) | *[Select one from]*<br>- 01=Yes<br>- 00=No<br>- DK=Do not know<br>- NA=Not applicable<br>- DA=Prefer not to answer |
| **Apn2** | 3056<br>6 | Has anyone noticed that you stop breathing during your sleep? | | | *[Select one from]*<br>- 01=Almost every day<br>- 02=3-4 times per week<br>- 03=1-2 times per week<br>- 04=1-2 times per month<br>- 05=Rarely or never<br>- DK=Do not know<br>- NA=Not applicable<br>- DA=Prefer not to answer |
| **Apn3** | 3056<br>7 | How often do you feel tired or fatigued after your sleep? | | | *[Select one from]*<br>- 01=Almost every day<br>- 02=3-4 times per week<br>- 03=1-2 times per week<br>- 04=1-2 times per month<br>- 05=Rarely or never<br>- DA=Prefer not to answer |
| **Apn4** | 3056<br>8 | During your waking time, do you feel tired, fatigued or not up to par? | | | *[Select one from]*<br>- 01=Almost every day<br>- 02=3-4 times per week<br>- 03=1-2 times per week<br>- 04=1-2 times per month<br>- 05=Rarely or never<br>- DA=Prefer not to answer |
| **Apn5** | 3056<br>9 | Have you ever nodded off or fallen asleep while driving a vehicle? | | | *[Select one from]*<br>- 01=Yes<br>- 00=No<br>- NA=Not applicable<br>- DA=Prefer not to answer |
| **Apn5a** | 3057<br>0 | How often have you nodded off or fallen asleep while driving a vehicle? | | | *[Select one from]*<br>- 01=Almost every day<br>- 02=3-4 times a week<br>- 03=1-2 times per week<br>- 04=1-2 times per month<br>- 05=Rarely or never<br>- DA=Prefer not to answer |
| **Apn6** | 3057<br>1 | Thinking about the last year, how many times have you had an accident (motor vehicle, home or work related) or a near miss due to sleepiness? | | | *[Select one from]*<br>- 01=More than 10 times<br>- 02=5-10 times<br>- 03=2-5 times<br>- 04=Once<br>- 00=Never had an accident or near miss due to sleepiness<br>- DK=Do not know<br>- DA=Prefer not to answer |
| **Fatigue** | | | | | |

| | | | | | |
|---|---|---|---|---|---|
| **FATIGUE-INTRO** | | We would like to know a little bit about your experience of fatigue. | | | |
| **FATIGUE-INTRO 1** | | We are interested in the extent that you have felt **fatigued** (tired, weary, exhausted) over the last **two weeks**. We **do not** mean feelings of **sleepiness** (the likelihood of falling asleep). Please select the appropriate response in accordance with your average feelings over this two-week period. | | | |
| **F1** | 30572 | Was fatigue a problem for you? | | | *[Select one from]*<br>- 00=0 - Not at all<br>- 01=1<br>- 02=2 - Moderately<br>- 03=3<br>- 04=4 - Extremely |
| **F2** | 30573 | Did fatigue cause problems with your everyday functioning (e.g., work, social, family)? | | Field ID: 120127 (Pain questionnaire) | *[Select one from]*<br>- 00=0 - Not at all<br>- 01=1<br>- 02=2 - Moderately<br>- 03=3<br>- 04=4 - Extremely |
| **F3** | 30574 | Did fatigue cause you distress? | | | *[Select one from]*<br>- 00=0 - Not at all<br>- 01=1<br>- 02=2 - Moderately<br>- 03=3<br>- 04=4 - Extremely |
| **F4** | 30575 | How often did you suffer from fatigue? | | | *[Select one from]*<br>- 00=0 days per week<br>- 01=1-2 days per week<br>- 02=3-4 days per week<br>- 03=5-6 days per week<br>- 04=7 days per week |
| **F5** | 30576 | At what time(s) of the day did you typically experience fatigue? (Please select all that apply) | | | *[Select one or more from]*<br>- 00=Early morning<br>- 01=Mid morning<br>- 02=Midday<br>- 03=Mid afternoon<br>- 04=Late afternoon<br>- 05=Early evening<br>- 06=Late evening |
| **F6** | 30577 | How severe was the fatigue you experienced? | | | *[Select one from]*<br>- 00=0 - Not at all<br>- 01=1<br>- 02=2 - Moderate<br>- 03=3<br>- 04=4 - Extreme |
| **F7** | 30578 | How much was your fatigue caused by poor sleep? | | | *[Select one from]*<br>- 00=0 - Not at all<br>- 01=1<br>- 02=2 - Moderate<br>- 03=3<br>- 04=4 - Extreme |
| **Restless legs** | | | | | |

| | | | | | |
|---|---|---|---|---|---|
| **RLSINTRO** | | We'd like to know about any issues that you might experience with restless legs. | | | |
| **RLS1** | 30579 | Do you have recurrent uncomfortable feelings or sensations in your legs while you are sitting or lying down? | | | *[Select one from]*<br>- 01=Yes<br>- 00=No<br>- DK=Do not know<br>- DA=Prefer not to answer |
| **RLS2** | 30580 | Do you have a recurrent need or urge to move your legs while you are sitting or lying down? | | | *[Select one from]*<br>- 01=Yes<br>- 00=No<br>- DK=Do not know<br>- DA=Prefer not to answer |
| **RLS3** | 30581 | Are you more likely to have these feelings in your legs when you are resting (either sitting or lying down) or when you are physically active? | | | *[Select one from]*<br>- 01=Resting<br>- 00=Active<br>- DK=Do not know<br>- DA=Prefer not to answer |
| **RLS4** | 30582 | If you get up or move around when you have these feelings in your legs, do these feelings get any better while you actually keep moving? | | | *[Select one from]*<br>- 01=Yes<br>- 00=No<br>- DK=Do not know<br>- DA=Prefer not to answer |
| **RLS5** | 30583 | Which time(s) of day are these feelings in your legs most likely to occur?<br>(Please select one or more responses.) | | | *[Select one or more from 01-05. 06, DK and DA are exclusive. If RLS5=About equal at all times (06) or do not know (DK) or Prefer not to answer (DA), no other selection can be made.]*<br>- 01=Morning<br>- 02=Mid-day<br>- 03=Afternoon<br>- 04=Evening<br>- 05=Night<br>- 06=About equal at all times<br>- DK=Do not know<br>- DA=Prefer not to answer |
| **RLS6** | 30584 | Will simply changing leg position once without continuing to move usually relieve these feelings in your legs? | | | *[Select one from]*<br>- 01=Usually relieves<br>- 02= Does not usually relieve<br>- DK=Do not know<br>- DA=Prefer not to answer |
| **RLS7** | 30585 | Are these feelings in your legs always due to muscle cramps? | | | *[Select one from]*<br>- 01=Yes<br>- 00=No<br>- DK=Do not know<br>- DA=Prefer not to answer |
| **RLS8** | | In the past 12 months, how often did you experience these feelings in your legs? | | | *[Select one from]*<br>- 01=6-7 days per week<br>- 02=4-5 days per week<br>- 03=2-3 days per week<br>- 04=1 day per week<br>- 05=2 days per month<br>- 06=1 day per month or less<br>- 00=Never<br>- DK=Do not know<br>- DA=Prefer not to answer |
| **Sleep consequences** | | | | | |
| **SSS-INTRO** | | We would like you to tell us about situations that cause you to fall asleep | | | |

| | | | | | |
|---|---|---|---|---|---|
| | | when you don't intend to. | | | |
| **SSS1** | 3207<br>1 | In the past two weeks, how likely is it that you would fall asleep without intending to, or that you would struggle to stay awake while you were doing things? | | Field ID:<br>1220<br>(Touchscreen questionnaire) | *[Select one from]*<br>- 00=No chance<br>- 01=Slight chance<br>- 02=Moderate chance<br>- 03=High chance<br>- DA=Prefer not to answer |
| **SSS1_ ALT** | 3207<br>2 | Over the past two weeks, how likely is it that you would unintentionally fall asleep or doze off while you were doing things? | | Field ID:<br>1220<br>(Touchscreen questionnaire) | *[Select one from]*<br>- 00=No chance<br>- 01=Slight chance<br>- 02=Moderate chance<br>- 03=High chance<br>- DA=Prefer not to answer |
| **SSS1_ ALT2** | 3207<br>3 | Over the past two weeks, how likely is it that you would unintentionally fall asleep or doze off? | | Field ID:<br>1220<br>(Touchscreen questionnaire) | *[Select one from]*<br>- 00=No chance<br>- 01=Slight chance<br>- 02=Moderate chance<br>- 03=High chance<br>- DA=Prefer not to answer |
| **BLOCK _SSS2** | | How likely are you to have difficulties staying awake in the following situations?<br>This refers to how you have felt in the last 2 weeks.<br><br>0=No chance<br>1=Slight chance<br>2=Moderate chance<br>3=High chance<br>NA=Not applicable<br><br>It is important that you answer each question as best as you can. | | | |
| **BLOCK _SSS2_ ALT** | | Over the past two weeks, how likely is it that you would unintentionally fall asleep or doze off while doing the following activities?<br><br>0=No chance<br>1=Slight chance<br>2=Moderate chance<br>3=High chance<br>NA=Not applicable<br><br>It is important that you answer each question as best as you can. | | | |
| **BLOCK _SSS2_ ALT2** | | Over the past two weeks, how likely is it that you would fall | | | |

| | | | | | |
|---|---|---|---|---|---|
| | | asleep or doze off while doing the following activities?<br><br>0=No chance<br>1=Slight chance<br>2=Moderate chance<br>3=High chance<br>NA=Not applicable<br><br>It is important that you answer each question as best as you can. | | | |
| **SSS2a** | 32074 | Sitting at a desk/table working on a computer or tablet | | | *[Select one from]*<br>- 0=No chance<br>- 1=Slight chance<br>- 2=Moderate chance<br>- 3=High chance<br>- NA=Not applicable |
| **SSS2a_ALT** | 32075 | Sitting at a desk/table working on a computer or tablet | | | *[Select one from]*<br>- 0=No chance<br>- 1=Slight chance<br>- 2=Moderate chance<br>- 3=High chance<br>- NA=Not applicable |
| **SSS2a_ALT2** | 32076 | Sitting at a desk/table working on a computer or tablet | | | *[Select one from]*<br>- 0=No chance<br>- 1=Slight chance<br>- 2=Moderate chance<br>- 3=High chance<br>- NA=Not applicable |
| **SSS2b** | 32077 | Talking to someone on the phone | | | *[Select one from]*<br>- 0=No chance<br>- 1=Slight chance<br>- 2=Moderate chance<br>- 3=High chance<br>- NA=Not applicable |
| **SSS2b_ALT** | 32078 | Talking to someone on the phone | | | *[Select one from]*<br>- 0=No chance<br>- 1=Slight chance<br>- 2=Moderate chance<br>- 3=High chance<br>- NA=Not applicable |
| **SSS2b_ALT2** | 32079 | Talking to someone on the phone | | | *[Select one from]*<br>- 0=No chance<br>- 1=Slight chance<br>- 2=Moderate chance<br>- 3=High chance<br>- NA=Not applicable |
| **SSS2c** | 32080 | In a meeting with several people | | | *[Select one from]*<br>- 0=No chance<br>- 1=Slight chance<br>- 2=Moderate chance<br>- 3=High chance<br>- NA=Not applicable |

| | | | | | |
|---|---|---|---|---|---|
| **SSS2c_ALT** | 32081 | In a meeting with several people | | | *[Select one from]*<br>- 0=No chance<br>- 1=Slight chance<br>- 2=Moderate chance<br>- 3=High chance<br>- NA=Not applicable |
| **SSS2c_ALT2** | 32082 | In a meeting with several people | | | *[Select one from]*<br>- 0=No chance<br>- 1=Slight chance<br>- 2=Moderate chance<br>- 3=High chance<br>- NA=Not applicable |
| **SSS2d** | 32083 | Listening to someone talking in a class, lecture or at church | | | *[Select one from]*<br>- 0=No chance<br>- 1=Slight chance<br>- 2=Moderate chance<br>- 3=High chance<br>- NA=Not applicable |
| **SSS2d_ALT** | 32084 | Listening to someone talking in a class, lecture or at church | | | *[Select one from]*<br>- 0=No chance<br>- 1=Slight chance<br>- 2=Moderate chance<br>- 3=High chance<br>- NA=Not applicable |
| **SSS2d_ALT2** | 32085 | Listening to someone talking in a class, lecture or at church | | | *[Select one from]*<br>- 0=No chance<br>- 1=Slight chance<br>- 2=Moderate chance<br>- 3=High chance<br>- NA=Not applicable |
| **SSS2e** | 32086 | Playing cards or a board game with others | | | *[Select one from]*<br>- 0=No chance<br>- 1=Slight chance<br>- 2=Moderate chance<br>- 3=High chance<br>- NA=Not applicable |
| **SSS2e_ALT** | 32087 | Playing cards or a board game with others | | | *[Select one from]*<br>- 0=No chance<br>- 1=Slight chance<br>- 2=Moderate chance<br>- 3=High chance<br>- NA=Not applicable |
| **SSS2e_ALT2** | 32088 | Playing cards or a board game with others | | | *[Select one from]*<br>- 0=No chance<br>- 1=Slight chance<br>- 2=Moderate chance<br>- 3=High chance<br>- NA=Not applicable |
| **SSS2f** | 32089 | Driving a car | | | *[Select one from]*<br>- 0=No chance<br>- 1=Slight chance<br>- 2=Moderate chance<br>- 3=High chance<br>- NA=Not applicable |

| | | | | | |
|---|---|---|---|---|---|
| **SSS2f_ ALT** | 3209 0 | Driving a car | | | *[Select one from]*<br>- 0=No chance<br>- 1=Slight chance<br>- 2=Moderate chance<br>- 3=High chance<br>- NA=Not applicable |
| **SSS2f_ ALT2** | 3209 1 | Driving a car | | | *[Select one from]*<br>- 0=No chance<br>- 1=Slight chance<br>- 2=Moderate chance<br>- 3=High chance<br>- NA=Not applicable |
| **SSS2g** | 3209 2 | Playing a videogame | | | *[Select one from]*<br>- 0=No chance<br>- 1=Slight chance<br>- 2=Moderate chance<br>- 3=High chance<br>- NA=Not applicable |
| **SSS2g_ ALT** | 3209 3 | Playing a videogame | | | *[Select one from]*<br>- 0=No chance<br>- 1=Slight chance<br>- 2=Moderate chance<br>- 3=High chance<br>- NA=Not applicable |
| **SSS2g_ ALT2** | 3209 4 | Playing a videogame | | | *[Select one from]*<br>- 0=No chance<br>- 1=Slight chance<br>- 2=Moderate chance<br>- 3=High chance<br>- NA=Not applicable |
| **SSS2h** | 3209 5 | Lying down trying to take a nap | | | *[Select one from]*<br>- 0=No chance<br>- 1=Slight chance<br>- 2=Moderate chance<br>- 3=High chance<br>- NA=Not applicable |
| **SSS2h_ ALT** | 3209 6 | Lying down trying to take a nap | | | *[Select one from]*<br>- 0=No chance<br>- 1=Slight chance<br>- 2=Moderate chance<br>- 3=High chance<br>- NA=Not applicable |
| **SSS2_ ALT2** | 3209 7 | Lying down to rest | | | *[Select one from]*<br>- 0=No chance<br>- 1=Slight chance<br>- 2=Moderate chance<br>- 3=High chance<br>- NA=Not applicable |
| **SSS2i** | 3209 8 | Travelling as a passenger in a bus, train or car for more than 30 minutes | | | *[Select one from]*<br>- 0=No chance<br>- 1=Slight chance<br>- 2=Moderate chance<br>- 3=High chance<br>- NA=Not applicable |

| | | | | | |
|---|---|---|---|---|---|
| **SSS2i_ ALT** | 32099 | Travelling as a passenger in a bus, train or car for more than 30 minutes | | | *[Select one from]*<br>- 0=No chance<br>- 1=Slight chance<br>- 2=Moderate chance<br>- 3=High chance<br>- NA=Not applicable |
| **SSS2i_ ALT2** | 32100 | Travelling as a passenger in a bus, train or car for more than 30 minutes | | | *[Select one from]*<br>- 0=No chance<br>- 1=Slight chance<br>- 2=Moderate chance<br>- 3=High chance<br>- NA=Not applicable |
| **SSS2j** | 32101 | Watching a movie/film at home | | | *[Select one from]*<br>- 0=No chance<br>- 1=Slight chance<br>- 2=Moderate chance<br>- 3=High chance<br>- NA=Not applicable |
| **SSS2j_ ALT** | 32102 | Watching a movie/film at home | | | *[Select one from]*<br>- 0=No chance<br>- 1=Slight chance<br>- 2=Moderate chance<br>- 3=High chance<br>- NA=Not applicable |
| **SSS2j_ ALT2** | 32103 | Watching a movie/film at home | | | *[Select one from]*<br>- 0=No chance<br>- 1=Slight chance<br>- 2=Moderate chance<br>- 3=High chance<br>- NA=Not applicable |
| **BLOCK -CC1** | | Please rate your problems with concentration, memory, and thinking skills **during the past 7 days**. | | | |
| **CC1a** | 32104 | Forgetfulness/memory problems | | | *[Select one from]*<br>- 00=Not at all<br>- 01=Some<br>- 02=Quite a bit<br>- 03=Very much<br>- DA=Prefer not to answer |
| **CC1b** | 32105 | Poor concentration | | | *[Select one from]*<br>- 00=Not at all<br>- 01=Some<br>- 02=Quite a bit<br>- 03=Very much<br>- DA=Prefer not to answer |
| **CC1c** | 32106 | Trouble expressing my thoughts | | | *[Select one from]*<br>- 00=Not at all<br>- 01=Some<br>- 02=Quite a bit<br>- 03=Very much<br>- DA=Prefer not to answer |
| **CC1d** | 32107 | Trouble finding the right word | | | *[Select one from]*<br>- 00=Not at all<br>- 01=Some<br>- 02=Quite a bit<br>- 03=Very much |

| | | | | | |
|---|---|---|---|---|---|
| | | | | | - DA=Prefer not to answer |
| **CC1e** | 3210 8 | Slow thinking speed | | | *[Select one from]*<br>- 00=Not at all<br>- 01=Some<br>- 02=Quite a bit<br>- 03=Very much<br>- DA=Prefer not to answer |
| **CC1f** | 3210 9 | Trouble figuring things out or solving problems | | | *[Select one from]*<br>- 00=Not at all<br>- 01=Some<br>- 02=Quite a bit<br>- 03=Very much<br>- DA=Prefer not to answer |
| **Family history** | | | | | |
| **FH-INTRO** | | We would like to know a little bit about the sleep patterns of your family members. | | | |
| **FH1** | 3211 0 | How many full brothers and sisters (living or deceased) do you have from the same birth parents? | | | *[Select one from]*<br>- 00=0<br>- 01=1<br>- 02=2<br>- 03=3<br>- 04=4<br>- 05=5<br>- 06=6<br>- 07=7 or more<br>- DK=Do not know<br>- DA=Prefer not to answer |
| **BLOCK -FH2** | | Have any close BLOOD relatives (including brother/sister, mother/father, son/daughter) had any of the following? | | | |
| **FH2a** | 3211 1 | Insomnia (difficulty falling asleep or staying asleep) | | | *[Select one from]*<br>- 01=Yes<br>- 00=No<br>- DK=Do not know<br>- DA=Prefer not to answer |
| **FH2b** | 3211 2 | Sleep apnoea (breathing pauses during sleep) | | | *[Select one from]*<br>- 01=Yes<br>- 00=No<br>- DK=Do not know<br>- DA=Prefer not to answer |
| **FH2c** | 3211 3 | Narcolepsy (difficulty staying awake or having “sleep attacks”) | | | *[Select one from]*<br>- 01=Yes<br>- 00=No<br>- DK=Do not know<br>- DA=Prefer not to answer |
| **FH2d** | 3211 4 | Restless leg syndrome (RLS) (uncontrollable urge to move the legs) | | | *[Select one from]*<br>- 01=Yes<br>- 00=No<br>- DK=Do not know<br>- DA=Prefer not to answer |
| **FH2e** | 3211 5 | Sleep walking | | | *[Select one from]*<br>- 01=Yes<br>- 00=No<br>- DK=Do not know<br>- DA=Prefer not to answer |
| **FH2f** | 3211 6 | Night terrors (partial waking from sleep with | | | *[Select one from]*<br>- 01=Yes |

| | | | | | |
|---|---|---|---|---|---|
| | | behaviours such as screaming, kicking, panic, sleep walking, thrashing or mumbling) | | | - 00=No<br>- DK=Do not know<br>- DA=Prefer not to answer |
| **Lifestyle routines** | | | | | |
| **BLOCK -LR1** | | During the past month how often have you done any of the following? | | | |
| **LR1a** | 32117 | Used a sleep-tracking device to monitor your sleep pattern? For example, using a wristband (e.g. Fitbit) or smartphone app? | | | *[Select one from]*<br>- 01=Daily<br>- 02=More than once a week<br>- 03=3-4 times<br>- 04=1-2 times<br>- 05=Have a sleep-tracking device but did not use it in last month<br>- NA=Do not have a sleep-tracking device<br>- DA=Prefer not to answer |
| **LR1b** | 32118 | Travelled to other time zones? | | | *[Select one from]*<br>- 01=Daily<br>- 02=More than once a week<br>- 03=3-4 times<br>- 04=1-2 times<br>- 05=Not at all<br>- DA=Prefer not to answer |
| **LR1c** | 32119 | Consumed alcohol to help you fall asleep? | | | *[Select one from]*<br>- 01=Daily<br>- 02=More than once a week<br>- 03=3-4 times<br>- 04=1-2 times<br>- 05=Not at all<br>- DA=Prefer not to answer |
| **LR1d** | 30475 | Napped or dozed during the day? | | Field ID: 1190 (Touchscreen questionnaire) | *[Select one from]*<br>- 01=Daily<br>- 02=More than once a week<br>- 03=3-4 times<br>- 04=1-2 times<br>- 05=Not at all<br>- DA=Prefer not to answer |
| **LR1di** | 30476 | When you do nap during the day, how long do you typically nap for? | | | *[Select one from]*<br>- 01=0-20 minutes<br>- 02=21-40 minutes<br>- 03=41-60 minutes<br>- 04=1-2 hours<br>- 05=More than 2 hours<br>- DK=Do not know<br>- DA=Prefer not to answer |
| **LR1e** | 30477 | Exercised in a way that required a moderate amount of effort or noticeably accelerated your heart rate? | | | *[Select one from]*<br>- 01=Daily<br>- 02=More than once a week<br>- 03=3-4 times<br>- 04=1-2 times<br>- 05=Not at all<br>- NA=Unable to exercise<br>- DA=Prefer not to answer |
| **LR1ei** | 30478 | What time of the day do you usually exercise? | | | *[Select one from]*<br>- 01=Early morning<br>- 02=Late morning<br>- 03=Early afternoon<br>- 04=Late afternoon<br>- 05=Between evening meal and bedtime<br>- 06=Varies significantly |

| | | | | | |
|---|---|---|---|---|---|
| | | | | | - DA=Prefer not to answer |
| **LR2** | 3047<br>9 | How long before bedtime do you last use a computer, tablet, mobile phone or television? | | | *[Select one from]*<br>- 01=I use them in bed<br>- 02=Less than 1 hour<br>- 03=1-2 hours<br>- 04=2-3 hours<br>- 05=3 hours or longer<br>- NA=Not applicable<br>- DA=Prefer not to answer |
| **LR3** | 3048<br>0 | How many servings of caffeine do you typically have in one day? (one serving equals one small mug of tea or coffee, or one can of caffeinated soft drinks, e.g. cola or energy drink) | | | - Number<br>OR<br>- 01=I rarely or never drink caffeine<br>- DA=Prefer not to answer |
| **LR3a** | 3048<br>1 | What time of day do you usually drink your last caffeinated drink? | | | *[Select one from]*<br>- 01=Early morning<br>- 02=Late morning<br>- 03=Early afternoon<br>- 04=Late afternoon<br>- 05=Between evening meal and bedtime<br>- 06=Varies significantly<br>- DA=Prefer not to answer |
| **LR4** | 3048<br>2 | In a typical day in summer, how many hours do you spend outdoors? | Field ID:<br>1050<br>(Touchscr een questionn aire) | | - Number<br>OR<br>-10=Less than an hour a day<br>-1=Do not know<br>-3=Prefer not to answer |
| **LR5** | 3048<br>3 | In a typical day in winter, how many hours do you spend outdoors? | Field ID:<br>1060<br>(Touchscr een questionn aire) | | - Number<br>OR<br>-10=Less than an hour a day<br>-1=Do not know<br>-3=Prefer not to answer |
| Recent feelings | | | | | |
| **PHQ-4 INTRO** | | We would like to know about how you have been feeling recently. | | | |
| **BLOCK PHQ-4 1** | | Over the last 2 weeks, how often have you been bothered by any of the following problems? | | | |
| **PHQ-4 1a** | 3048<br>4 | Feeling nervous, anxious or on edge | Field ID:<br>20506<br>(Mental health questionn aire)<br><br>Field ID:<br>28735<br>(Health and well-being questionn aire) | | *[Select one from]*<br>- 01=Not at all<br>- 02=Several days<br>- 03=More than half the days<br>- 04=Nearly every day<br>- DA=Prefer not to answer |

| | | | | | |
|---|---|---|---|---|---|
| | | | Field ID: 29058 (Mental well-being questionnaire) | | |
| **PHQ-4 1b** | 30485 | Not being able to stop or control worrying | Field ID: 20509 (Mental health questionnaire)<br><br>Field ID: 28736 (Health and well-being questionnaire)<br><br>Field ID: 29059 (Mental well-being questionnaire) | | *[Select one from]*<br>- 01=Not at all<br>- 02=Several days<br>- 03=More than half the days<br>- 04=Nearly every day<br>- DA=Prefer not to answer |
| **PHQ-4 1c** | 30486 | Little interest or pleasure in doing things | Field ID: 20514 (Mental health questionnaire)<br><br>Field ID: 120104 (Pain questionnaire)<br><br>Field ID: 28737 (Health and well-being questionnaire)<br><br>Field ID: 29002 (Mental well-being questionnaire) | | *[Select one from]*<br>- 01=Not at all<br>- 02=Several days<br>- 03=More than half the days<br>- 04=Nearly every day<br>- DA=Prefer not to answer |
| **PHQ-4 1d** | 30487 | Feeling down, depressed or hopeless | Field ID: 20510 (Mental | | *[Select one from]*<br>- 01=Not at all<br>- 02=Several days |

| | | | | | |
|---|---|---|---|---|---|
| | | | health questionnaire)<br><br>Field ID: 120105 (Pain questionnaire)<br><br>Field ID: 28738 (Health and well-being questionnaire)<br><br>Field ID: 29003 (Mental well-being questionnaire) | | - 03=More than half the days<br>- 04=Nearly every day<br>- DA=Prefer not to answer |

**Appendix 2:** Questions with multiple versions

**Table 1.** Revisions in the 'Work and sleep' module

| Version 1 (n = 62,871) | | | Version 2 (n = 121,275) | | |
|---|---|---|---|---|---|
| **Field ID** | **Stem** | **Responses** | **Field ID** | **Stem** | **Responses** |
| | This module is about your work patterns. | | | This module is about your work patterns. ***Please complete this module even if you are not currently working, for example if you are retired.*** | |
| 30430 | In the past month, did you typically work a non-standard shift schedule?<br><br>Here we define non-standard shift schedule as working hours outside the hours of 8am to 8pm. | - 01=No – I didn't work<br>- 02=No – I worked standard hours (e.g. 9:00am-5:00pm)<br>- 03=Yes – I worked evening shifts (work typically finished between 8:00pm and midnight)<br>- 04=Yes - I worked morning shifts (work typically started between 4:00am and 7:00am)<br>- 05=Yes – I worked night shifts (work typically took place between 8:00pm and 8:00am)<br>- 06=Yes – I worked rotating shifts or had irregular work hours<br>DA=Prefer not to answer | 30431 | In the past month, did you typically work a non-standard shift schedule?<br><br>Here we define non-standard shift schedule as working hours outside the hours of 8am to 8pm.<br>***If you are retired, please answer "No, I didn't work".*** | - 01=No – I didn't work<br>- 02=No – I worked standard hours (e.g. 9:00am-5:00pm)<br>- 03=Yes – I worked evening shifts (work typically finished between 8:00pm and midnight)<br>- 04=Yes - I worked morning shifts (work typically started between 4:00am and 7:00am)<br>- 05=Yes – I worked night shifts (work typically took place between 8:00pm and 8:00am)<br>- 06=Yes – I worked rotating shifts or had irregular work hours<br>DA=Prefer not to answer |
| **Version 1 (n = 62,147)** | | | **Version 2 (n = 120,625)** | | |

| Field ID | Stem | Responses | Field ID | Stem | Responses |
|---|---|---|---|---|---|
| 30438 | On non-working days in the past month, what time did you typically fall asleep?<br>(This may be different to the time you went to bed)” | - Menu5 Hours from 9:00pm to 8:00pm in hour increments<br>- Menu6 Minutes from 00 to 55 in 5 minute increments<br>OR<br>- V=Varies significantly<br>- DA=Prefer not to answer | 30439 | On non-working days in the past month, what time did you typically fall asleep?<br>(This may be different from the time you went to bed).<br><br>***By non-working days, we mean any days that you did not work, including if you are retired.*** | - Menu5 Hours from 9:00pm to 8:00pm in hour increments<br>- Menu6 Minutes from 00 to 55 in 5 minute increments<br>OR<br>- V=Varies significantly<br>- DA=Prefer not to answer |
| 30440 | On non-working days in the past month, what time did you typically wake up?<br>(By ‘wake up’ we mean your final awakening time before getting up for the day) | - Menu7 Hours from 5:00am to 4:00am in hour increments<br>- Menu8 Minutes from 00 to 55 in 5 minute increments<br>OR<br>- V=Varies significantly<br>- DA=Prefer not to answer | 30441 | On non-working days in the past month, what time did you typically wake up?<br>(By ‘wake up’ we mean your final awakening time before getting up for the day)<br>***By non-working days, we mean any days that you did not work, including if you are retired.*** | - Menu7 Hours from 5:00am to 4:00am in hour increments<br>- Menu8 Minutes from 00 to 55 in 5 minute increments<br>OR<br>- V=Varies significantly<br>- DA=Prefer not to answer- 00 No<br>- DA Prefer not to answer |

**Table 2.** Revisions in the 'sleep consequences' module

| Version 1 (n = 7,826) | | | Version 2 (n = 52,907) | | | Version 3 (n = 118,605) | | |
|---|---|---|---|---|---|---|---|---|
| Field ID | Stem | Responses | Field ID | Stem | Responses | Field ID | Stem | Responses |
| 32071 | In the past two weeks, how likely is it that you would fall asleep without intending to, or that you would struggle to stay awake while you were doing things? | - 00=No chance<br>- 01=Slight chance<br>- 02=Moderate chance<br>- 03=High chance<br>- DA=Prefer not to answer | 32072 | ***Over*** the past two weeks, how likely is it that you would ***unintentionally fall asleep or doze off*** while you were doing things? | - 00=No chance<br>- 01=Slight chance<br>- 02=Moderate chance<br>- 03=High chance<br>- DA=Prefer not to answer | 32073 | Over the past two weeks, how likely is it that you would unintentionally fall asleep or doze off? | - 00=No chance<br>- 01=Slight chance<br>- 02=Moderate chance<br>- 03=High chance<br>- DA=Prefer not to answer |
| | How likely are you to have difficulties staying awake in the following situations? This refers to how you have felt in the last 2 weeks.<br><br>0=No chance<br>1=Slight chance<br>2=Moderate chance<br>3=High chance<br>NA=Not applicable<br><br>It is important that you answer each question as best as you can. | | | Over the past two weeks, how likely is it that you would unintentionally fall asleep or doze off while doing the following activities?<br><br>0=No chance<br>1=Slight chance<br>2=Moderate chance<br>3=High chance<br>NA=Not applicable<br><br>It is important that you answer each question as best as you can. | | | Over the past two weeks, how likely is it that you would ***fall asleep or doze off while doing the following activities***?<br><br>0=No chance<br>1=Slight chance<br>2=Moderate chance<br>3=High chance<br>NA=Not applicable<br><br>It is important that you answer each question as best as you can. | |

| | | | | | | | | |
|---|---|---|---|---|---|---|---|---|
| 32074 | Sitting at a desk/table working on a computer or tablet | - 0=No chance<br>- 1=Slight chance<br>- 2=Moderate chance<br>- 3=High chance<br>- NA=Not applicable | 32075 | Sitting at a desk/table working on a computer or tablet | - 0=No chance<br>- 1=Slight chance<br>- 2=Moderate chance<br>- 3=High chance<br>- NA=Not applicable | 32076 | Sitting at a desk/table working on a computer or tablet | - 0=No chance<br>- 1=Slight chance<br>- 2=Moderate chance<br>- 3=High chance<br>- NA=Not applicable |
| 32077 | Talking to someone on the phone | - 0=No chance<br>- 1=Slight chance<br>- 2=Moderate chance<br>- 3=High chance<br>- NA=Not applicable | 32078 | Talking to someone on the phone | - 0=No chance<br>- 1=Slight chance<br>- 2=Moderate chance<br>- 3=High chance<br>- NA=Not applicable | 32079 | Talking to someone on the phone | - 0=No chance<br>- 1=Slight chance<br>- 2=Moderate chance<br>- 3=High chance<br>- NA=Not applicable |
| 32080 | In a meeting with several people | - 0=No chance<br>- 1=Slight chance<br>- 2=Moderate chance<br>- 3=High chance<br>- NA=Not applicable | 32081 | In a meeting with several people | - 0=No chance<br>- 1=Slight chance<br>- 2=Moderate chance<br>- 3=High chance<br>- NA=Not applicable | 32082 | In a meeting with several people | - 0=No chance<br>- 1=Slight chance<br>- 2=Moderate chance<br>- 3=High chance<br>- NA=Not applicable |
| 32083 | Listening to someone talking in a class, lecture or at church | - 0=No chance<br>- 1=Slight chance<br>- 2=Moderate chance<br>- 3=High chance<br>- NA=Not applicable | 32084 | Listening to someone talking in a class, lecture or at church | - 0=No chance<br>- 1=Slight chance<br>- 2=Moderate chance<br>- 3=High chance<br>- NA=Not applicable | 32085 | Listening to someone talking in a class, lecture or at church | - 0=No chance<br>- 1=Slight chance<br>- 2=Moderate chance<br>- 3=High chance<br>- NA=Not applicable |
| 32086 | Playing cards or a board game with others | - 0=No chance<br>- 1=Slight chance<br>- 2=Moderate chance<br>- 3=High chance<br>- NA=Not applicable | 32087 | Playing cards or a board game with others | - 0=No chance<br>- 1=Slight chance<br>- 2=Moderate chance<br>- 3=High chance<br>- NA=Not applicable | 32088 | Playing cards or a board game with others | - 0=No chance<br>- 1=Slight chance<br>- 2=Moderate chance<br>- 3=High chance<br>- NA=Not applicable |

| | | | | | | | | |
|---|---|---|---|---|---|---|---|---|
| 32089 | Driving a car | - 0=No chance<br>- 1=Slight chance<br>- 2=Moderate chance<br>- 3=High chance<br>- NA=Not applicable | 32090 | Driving a car | - 0=No chance<br>- 1=Slight chance<br>- 2=Moderate chance<br>- 3=High chance<br>- NA=Not applicable | 32091 | Driving a car | - 0=No chance<br>- 1=Slight chance<br>- 2=Moderate chance<br>- 3=High chance<br>- NA=Not applicable |
| 32092 | Playing a videogame | - 0=No chance<br>- 1=Slight chance<br>- 2=Moderate chance<br>- 3=High chance<br>- NA=Not applicable | 32093 | Playing a videogame | - 0=No chance<br>- 1=Slight chance<br>- 2=Moderate chance<br>- 3=High chance<br>- NA=Not applicable | 32094 | Playing a videogame | - 0=No chance<br>- 1=Slight chance<br>- 2=Moderate chance<br>- 3=High chance<br>- NA=Not applicable |
| 32095 | Lying down trying to take a nap | - 0=No chance<br>- 1=Slight chance<br>- 2=Moderate chance<br>- 3=High chance<br>- NA=Not applicable | 32096 | Lying down trying to take a nap | - 0=No chance<br>- 1=Slight chance<br>- 2=Moderate chance<br>- 3=High chance<br>- NA=Not applicable | 32097 | Lying down ***to rest*** | - 0=No chance<br>- 1=Slight chance<br>- 2=Moderate chance<br>- 3=High chance<br>- NA=Not applicable |
| 32098 | Traveling as a passenger in a bus, train or car for more than 30 minutes | - 0=No chance<br>- 1=Slight chance<br>- 2=Moderate chance<br>- 3=High chance<br>- NA=Not applicable | 32099 | Traveling as a passenger in a bus, train or car for more than 30 minutes | - 0=No chance<br>- 1=Slight chance<br>- 2=Moderate chance<br>- 3=High chance<br>- NA=Not applicable | 32100 | Traveling as a passenger in a bus, train or car for more than 30 minutes | - 0=No chance<br>- 1=Slight chance<br>- 2=Moderate chance<br>- 3=High chance<br>- NA=Not applicable |
| 32101 | Watching a film at home or the cinema | - 0=No chance<br>- 1=Slight chance<br>- 2=Moderate chance<br>- 3=High chance<br>- NA=Not applicable | 32102 | Watching a film at home or the cinema | - 0=No chance<br>- 1=Slight chance<br>- 2=Moderate chance<br>- 3=High chance<br>- NA=Not applicable | 32103 | Watching a film at home or the cinema | - 0=No chance<br>- 1=Slight chance<br>- 2=Moderate chance<br>- 3=High chance<br>- NA=Not applicable |

#### **Appendix 3:** Phenotype coding

| Tables | Disorder Classification | Disorder | Disorder's fields and codes - POST DATA RELEASE (08.04.2025) |
|---|---|---|---|
| | | | * Dealing with midday bedtime and evening wake time<br>* Remove blood sibling requirements for family history of sleep disorders<br>* Dealing with SE > 100%<br>* Remove numerical coding to improve clarity<br>* OSA reclassification<br>* Sleep quality global measure, component 5, added 30461<br>* CTSWD code changes<br>* Sleep apnoea, added 30565 and 30569<br>* Remove narcolepsy case definitions |
| **Sleep disorders** | | | |
| | **1. Insomnia Disorder** | **1.1. Chronic Insomnia Disorder**<br>- SOL, WASO or EMA: > 30 mins<br>- Frequency of disturbance: ≥ 3 nights a week<br>- Sleep quality: average or poor or very poor<br>- Daytime functioning: somewhat, much or very much impaired<br>- Chronicity of problem: ≥ 3 months | <u>**Possible case** is defined as those meeting the following **insomnia disorder criteria**:</u><br><br>{<br>[SOL (30474) = (31-45 mins) or (46-60 mins) or (61 mins or more)]<br>OR<br>[WASO (30536) = (31-45 mins) or (46-60 mins) or (61 mins or more)]<br>OR<br>[EMA (30537) = (31-45 mins early) or (46-60 mins early) or (More than 60 mins early)]<br>}<br><br>AND<br>[Frequency (30538) = (3) or (4) or (5-7)]<br>AND<br>[Sleep quality (30539) = (Average) or (Poor) or (Very poor)]<br>AND<br><br>{<br>[Daytime functioning - mood, energy or relationships (30540) = (Somewhat) or (Much) or (Very much)]<br>OR<br>[Concentration, productivity or ability to stay awake (30541) = (Somewhat) or (Much) or (Very much)]<br>}<br><br>AND<br>[Chronicity of problem (30543) = (3-6 months) or (7-12 months) or (1-5 years) or (6-10 years) or (More than 10 years)] |

| | | | |
|---|---|---|---|
| | | ***1.1.1. Sleep Onset Insomnia only (subtype)***<br>- SOL: > 30 mins<br>- WASO and EMA: ≤ 30 mins<br>- Frequency of disturbance: ≥ 3 nights a week<br>- Sleep quality: average or poor or very poor<br>- Daytime functioning: somewhat, much or very much impaired<br>- Chronicity of problem: ≥ 3 months | [SOL (30474) = (31-45 mins) or (46-60 mins) or (61 mins or more)]<br>AND<br>[WASO (30536) = (0-15 mins) or (16-30 mins)]<br>AND<br>[EMA (30537) = (I don't wake up too early) or (Up to 15 mins early) or (16-30 mins early)]<br>AND<br>[Frequency (30538) = (3) or (4) or (5-7)]<br>AND<br>[Sleep quality (30539) = (Average) or (Poor) or (Very poor)]<br>AND<br><br>{<br>[Daytime functioning - mood, energy or relationships (30540) = (Somewhat) or (Much) or (Very much)]<br>OR<br>[Concentration, productivity or ability to stay awake (30541) = (Somewhat) or (Much) or (Very much)]<br>}<br><br>AND<br>[Chronicity of problem (30543) = (3-6 months) or (7-12 months) or (1-5 years) or (6-10 years) or (More than 10 years)] |

| | | | |
|---|---|---|---|
| | | ***1.1.2. Sleep Maintenance Insomnia only (subtype)***<br>- WASO: > 30 mins<br>- SOL and EMA: ≤ 30 mins<br>- Frequency of disturbance: ≥ 3 nights a week<br>- Sleep quality: average or poor or very poor<br>- Daytime functioning: somewhat, much or very much impaired<br>- Chronicity of problem: ≥ 3 months | [SOL (30474) = (0-15 mins) or (16-30 mins)]<br>AND<br>[WASO (30536) = (31-45 mins) or (46-60 mins) or (61 mins or more)]<br>AND<br>[EMA (30537) = (I don't wake up too early) or (Up to 15 mins early) or (16-30 mins early)]<br>AND<br>[Frequency (30538) = (3) or (4) or (5-7)]<br>AND<br>[Sleep quality (30539) = (Average) or (Poor) or (Very poor)]<br>AND<br><br>{<br>[Daytime functioning - mood, energy or relationships (30540) = (Somewhat) or (Much) or (Very much)]<br>OR<br>[Concentration, productivity or ability to stay awake (30541) = (Somewhat) or (Much) or (Very much)]<br>}<br><br>AND<br>[Chronicity of problem (30543) = (3-6 months) or (7-12 months) or (1-5 years) or (6-10 years) or (More than 10 years)] |

| | | | |
|---|---|---|---|
| | | ***1.1.3. Early Morning Awakening Insomnia only (subtype)***<br>- EMA: > 30 mins<br>- SOL and WASO: ≤ 30 mins<br>- Frequency of disturbance: ≥ 3 nights a week<br>- Sleep quality: average or poor or very poor<br>- Daytime functioning: somewhat, much or very much impaired<br>- Chronicity of problem: ≥ 3 months | [SOL (30474) = (0-15 mins) or (16-30 mins)]<br>AND<br>[WASO (30536) = (0-15 mins) or (16-30 mins)]<br>AND<br>[EMA (30537) = (31-45 mins early) or (46-60 mins early) or (More than 60 mins early)]<br>AND<br>[Frequency (30538) = (3) or (4) or (5-7)]<br>AND<br>[Sleep quality (30539) = (Average) or (Poor) or (Very poor)]<br>AND<br><br>{<br>[Daytime functioning - mood, energy or relationships (30540) = (Somewhat) or (Much) or (Very much)]<br>OR<br>[Concentration, productivity or ability to stay awake (30541) = (Somewhat) or (Much) or (Very much)]<br>}<br><br>AND<br>[Chronicity of problem (30543) = (3-6 months) or (7-12 months) or (1-5 years) or (6-10 years) or (More than 10 years)] |

| | | | |
|---|---|---|---|
| | **2. Sleep-Related Breathing Disorders** | **2.1. Obstructive Sleep Apnoea** | **Possible case** is defined as those considered **high risk** of sleep apnoea<br><br>Sleep apnea risk is categorised into 'high risk', 'medium-to-high risk', 'low-to-medium risk', and 'low risk' based on the scoring of the following two categories:<br><br>**Category 1 - Snoring and breathing (≥ 2 points = positive):**<br>[Snore (30562) = (Yes)]: 1 point<br>+<br>[Snore type (30563) = (Louder than talking)]: 1 point<br>+<br>[Snore frequency (30564) = (almost everyday) or (3-4 times per week)]: 1 point<br>+<br>[Snore bothered others (30565) = (yes)]: 1 point<br>+<br>[Stop breathing (30566) = (almost everyday) or (3-4 times per week)]: 2 points<br><br>**Category 2 - Fatigue and tired (≥ 2 points = positive):**<br>[After wake (30567) = (almost everyday) or (3-4 times per week)] :1 point<br>+<br>[During wake (30568) = (almost everyday) or (3-4 times per week)]: 1 point<br>+<br>{[Driving (30569) = (yes)]<br>AND<br>[Frequency (30570) = (almost everyday) or (3-4 times per week)]}: 1 point<br><br>If 30569 falling asleep when driving is No, then the driving component gets 0 point instead of missing.<br><br>**Overall scoring:**<br>'High risk' of sleep apnoea if both categories 1 and 2 are positive<br>'Medium-to-high risk' of sleep apnoea if only category 1 is positive (regardless of whether category 2 is negative or missing)<br>'Low-to-medium risk' of sleep apnoea if only category 2 is positive (regardless of whether category 1 is negative or missing)<br>'Low risk' is both categories = negative OR one category is negative and the other is missing.<br>"Missing" if both categories are missing. |

| | | | |
|---|---|---|---|
| | **3. Circadian Rhythm Sleep-Wake Disorders** | **3.1. Delayed Sleep-Wake Phase Disorder** | **Possible case** is defined as:<br><br>[Wrong time (30544) = 01 (Yes)]<br>AND<br>[Stay up late (30545) = 01 (Yes)]<br>AND<br>[Sleep at normal waking hours (30546) = 01 (Yes)]<br>AND<br>[Eveningness (30425 - 30429; Chronotype summary score; see below) = Moderately evening type (8-11) or Definitely evening type (4-7)] |
| | | **3.2. Advanced Sleep-Wake Phase Disorder** | **Possible case** is defined as:<br><br>[Wrong time (30544) = 01 (Yes)]<br>AND<br>[Sleep early (30547) = 01 (Yes)]<br>AND<br>[Wake early (30548) = 01 (Yes)]<br>AND<br>[Morningness (30425 - 30429; Chronotype summary score; see below) = Definitely morning type (22-25) or Moderately morning type (18-21)] |
| | | **3.3. Shift Work Disorder** | **Possible case** is defined as Shiftwork Disorder based on following items:<br><br>[Shift work schedule in past month (30430 or 30431) = (evening shifts, 8:00pm to midnight) or (morning shifts, 4:00am to 7:00am) or (night shifts, 8:00pm to 8:00am) or (rotating shifts or irregular work hours)]<br><br>AND, while working shifts:<br><br>{<br>[Problem with waking up too early and difficulty getting back to sleep (30432) = (Yes, a minor problem) or (Yes, a considerable problem) or (Yes, a serious problem)]<br>OR<br>[Doze off at work (30434) = (Slightly likely) or (Moderately likely) or (Highly likely)]<br>}<br><br>AND<br>[Sense of well-being (30433) = (Fairly bad) or (Very bad)] |
| | **4. Parasomnias** | **4.1. NREM-Related Parasomnias** | |

| | | | |
|---|---|---|---|
| | | ***4.1.1. Sleepwalking*** | **Possible case** is defined as those with **frequent** sleepwalking<br><br>Sleepwalking is categorised into frequent sleepwalking and infrequent sleepwalking (versus never) based on responses to the following question:<br><br>**Frequent sleepwalking:**<br>[Sleep walking (30555) = (1-2 times per month) or (1-2 times per week) or (3-4 times per week) or (Almost every day) or (Every day)]<br><br>**Infrequent sleepwalking:**<br>[Sleep walking (30555) = (Not in the past year) or (1-2 times per year)] |
| | | **4.2. REM-Related Parasomnias** | |
| | | ***4.2.1. REM Sleep Behaviour Disorder*** | **Possible case** is defined as:<br><br>[Act out dream (30557) = (Yes)]<br>AND<br>[Violent behaviour (30558) = (1-2 times per month) or (1-2 times per week) or (3-4 times per week) or (Almost every day) or (Every day)] |
| | | ***4.2.2. Nightmare Disorder*** | **Possible case** is defined as:<br><br>[Nightmare (30559) = (1-2 times per week) or (3-4 times per week) or (Almost every day) or (Every day)] |
| | **5. Sleep-Related Movement Disorders** | **5.1. Restless Legs Syndrome** | **Possible case** is defined as those with **persistent** RLS<br><br>To capture clinically RLS, as well as those on the path towards severe RLS, the three subtypes of RLS are categorised based on frequency of symptoms:<br><br>**Persistent RLS**<br>[Uncomfortable feeling (30579) = (Yes)]<br>AND<br>[Urge (30580) = (Yes)]<br>AND<br>[When resting (30581) = (Resting)]<br>AND<br>[Better when active (30582) = (Yes)]<br>AND<br>[Time of day (30583) = answer includes (Afternoon) or (Evening) or (Night), but NOT (Morning) or (Mid-day) or (About equal at all times)]<br>AND<br>[Leg position (30584) = (Does not usually relieve)]<br>AND<br>[Muscle cramps (30585) = (No)]<br>AND<br>[Frequency of symptoms (32070) = (6-7 days per week) or (4-5 days per week) or (2-3 days per week)] |

| | | | |
|---|---|---|---|
| | | | **Intermittent RLS**<br>[Uncomfortable feeling (30579) = (Yes)]<br>AND<br>[Urge (30580) = (Yes)]<br>AND<br>[When resting (30581) = (Resting)]<br>AND<br>[Better when active (30582) = (Yes)]<br>AND<br>[Time of day (30583) = (Afternoon) or (Evening) or (Night)]<br>AND<br>[Leg position (30584) = (Does not usually relieve)]<br>AND<br>[Muscle cramps (30585) = (No)]<br>AND<br>[Frequency of symptoms (32070) = (1 day per week) or (2 days per month)]<br><br>**Occasional RLS**<br>[Uncomfortable feeling (30579) = (Yes)]<br>AND<br>[Urge (30580) = (Yes)]<br>AND<br>[When resting (30581) = (Resting)]<br>AND<br>[Better when active (30582) = (Yes)]<br>AND<br>[Time of day (30583) = (Afternoon) or (Evening) or (Night)]<br>AND<br>[Leg position (30584) = (Does not usually relieve)]<br>AND<br>[Muscle cramps (30585) = (No)]<br>AND<br>[Frequency of symptoms (32070) = (1 day per month or less)] |
| **Potential correlates of sleep disorders** | | | |

| | | | |
|---|---|---|---|
| | **Sleepiness** | | Sleepiness is assessed by one global question:<br><br>Without intention (32071 / 32072 / 32073) [0=No chance, 1=Slight chance, 2=Moderate chance, 3=High chance]<br><br>And by new scale (Situational Sleepiness Scale; SSS) assessing likelihood of dozing/falling asleep in 10 specific situations. Total score ranges from 0-30. Higher scores indicate higher levels of sleepiness.<br><br>*Difficulties staying awake when:*<br>Sitting and working (32074 / 32075 / 32076) [0=No chance, 1=Slight chance, 2=Moderate chance, 3=High chance]<br>+<br>Talking to someone on the phone (32077 / 32078 / 32079) [0=No chance, 1=Slight chance, 2=Moderate chance, 3=High chance]<br>+<br>Meeting with several people (32080 / 32081 / 32082) [0=No chance, 1=Slight chance, 2=Moderate chance, 3=High chance]<br>+<br>Listening in lecture (32083 / 32084 / 32085) [0=No chance, 1=Slight chance, 2=Moderate chance, 3=High chance]<br>+<br>Playing cards (32086 / 32087 / 32088) [0=No chance, 1=Slight chance, 2=Moderate chance, 3=High chance]<br>+<br>Driving (32089 / 32090 / 32091) [0=No chance, 1=Slight chance, 2=Moderate chance, 3=High chance]<br>+<br>Paying a videogame (32092 / 32093 / 32094) [0=No chance, 1=Slight chance, 2=Moderate chance, 3=High chance]<br>+<br>Lying down trying to nap (32095 / 32096 / 32097) [0=No chance, 1=Slight chance, 2=Moderate chance, 3=High chance]<br>+<br>Travelling as a passenger (32098 / 32099 / 32100) [0=No chance, 1=Slight chance, 2=Moderate chance, 3=High chance]<br>+<br>Watching a film (32101 / 32102 / 32103) [0=No chance, 1=Slight chance, 2=Moderate chance, 3=High chance] |

| | | | |
|---|---|---|---|
| | **Fatigue** | | Level of fatigue is defined by the total score of the items below. Score ranges from 0-31. Higher scores indicate higher levels of fatigue.<br><br>Problem (30572) [00=0 - Not at all, 01=1, 02=2 - Moderately, 03=3, 04=4 - Extremely]<br>+<br>Functioning (30573) [00=0 - Not at all, 01=1, 02=2 - Moderately, 03=3, 04=4 - Extremely]<br>+<br>Distress (30574) [00=0 - Not at all, 01=1, 02=2 - Moderately, 03=3, 04=4 - Extremely]<br>+<br>Frequency (30575) [00=0 days per week, 01=1-2 days per week, 02=3-4 days per week, 03=5-6 days per week, 04=7 days per week]<br>+<br>Time of day *(30576) [00=Early morning, 01=Mid morning, 02=Midday , 03=Mid afternoon, 04=Late afternoon, 05=Early evening, 06=Late evening]<br>+<br>Severity (30577) [00=0 - Not at all, 01=1, 02=2 - Moderate, 03=3, 04=4 - Extreme]<br>+<br>Poor sleep (30578) [00=0 - Not at all, 01=1, 02=2 - Moderate, 03=3, 04=4 - Extreme]<br><br>*1 point for each response for time of day Q, such that 2 responses (early morning and mid morning) equates to 2 points |
| | **Depression** | | Depression caseness is defined by ≥ 3 of the total score of the items below:<br><br>Little pleasure (30486) [0 (Not at all), 1 (Several days), 2 (More than half the days), 3 (Nearly every day)]<br>+<br>Feeling down (30487) [0 (Not at all), 1 (Several days), 2 (More than half the days), 3 (Nearly every day)] |
| | **Anxiety** | | Anxiety caseness is defined by ≥ 3 of the total score of the items below:<br><br>Anxious (30484) [0 (Not at all), 1 (Several days), 2 (More than half the days), 3 (Nearly every day)]<br>+<br>Worry (30485) [0 (Not at all), 1 (Several days), 2 (More than half the days), 3 (Nearly every day)] |

| | | | |
|---|---|---|---|
| | **Cognitive impairment** | | Cognitive complaint is defined by the total score of the items below. Score ranges from 0-18. Higher scores indicate greater severity of cognitive complaints.<br><br>**Scoring per questions:**<br><br>Forgetfulness (32104) [00=Not at all, 01=Some, 02=Quite a bit, 03=Very much]<br>+<br>Poor concentration (32105) [00=Not at all, 01=Some, 02=Quite a bit, 03=Very much]<br>+<br>Trouble expressing my thoughts (32106) [00=Not at all, 01=Some, 02=Quite a bit, 03=Very much]<br>+<br>Trouble finding the right word (32107) [00=Not at all, 01=Some, 02=Quite a bit, 03=Very much]<br>+<br>Slow thinking speed (32108) [00=Not at all, 01=Some, 02=Quite a bit, 03=Very much]<br>+<br>Trouble solving problems (32109) [00=Not at all, 01=Some, 02=Quite a bit, 03=Very much] |
| | **Accidents** | | Descriptive summary:<br><br>Accident due to sleepiness - frequency (30571) [More than 10 times, 5-10 times, 2-5 times, Once, Never had an accident or near miss due to sleepiness] |
| **Sleep health dimensions** | | | |

| | | | |
|---|---|---|---|
| | **Sleep quality global measure** | Component 1 - Subjective sleep quality<br>Component 2 - Sleep latency<br>Component 3 - Sleep duration<br>Component 4 - Sleep efficiency<br>Component 5 - Sleep disturbance<br>Component 6 - Use of sleep medication<br>Component 7 - Daytime dysfunction | In scoring the PSQI, seven component scores are derived, each scored 0 (no difficulty) to 3 (severe difficulty). The component scores are summed to produce a global score (range 0 to 21). Higher scores indicate worse sleep quality.<br><br>Component 1 - Subjective sleep quality<br>Sleep quality (30467) [0 (Very good), 1 (Fairly good), 2 (Fairly bad), 3 (Very bad)]<br><br>Component 2 - Sleep latency<br>SOL (30443) [0 (<= 15 mins), 1 (16-30 mins), 2 (31-60 mins), 3 (> 60 mins)]<br>+<br>SOL > 30 (30446) [0 (Not during the past month, 1 (Less than once a week), 2 (Once or twice a week), 3 (Three or more times a week)]<br><br>Sum of the above subscores: 0 (0), 1 (1-2), 2 (3-4), 3 (5-6)<br><br>Component 3 - Sleep duration<br>TST (30445) [0 (>= 7 hrs), 1 (< 7 hrs and >= 6 hrs), 2 (< 6 hrs and >= 5 hrs), 3 (< 5 hrs)]<br><br>Component 4 - Sleep efficiency (see also row 32 for definition)<br>Hours slept = TST (30445)<br>Hours in bed = Wake time (30444) - Bedtime (30442)<br>Sleep efficiency = (hours slept / hours in bed) x 100<br><br>SE = 0 (>= 85%), 1 (75-84%), 2 (65-74%), 3 (< 65%)<br><br>1. For SE over 100%: round down people's SE if they are within 110% (basically allowing them mis-remember up to one hour assuming a 7.5 hour sleep duration)<br>2. Exclude all the outliers greater than 110%<br>3. Exlcude all the participants whose time in bed is < 3 hours or > 16 hours<br><br><br>Component 5 - Sleep disturbance<br>WASO, EMA (30447) [0 (Not during the past month), 1 (Less than once a week), 2 (Once or twice a week), 3 (Three or more times a week)<br>+<br>Bathroom (30448) [0 (Not during the past month), 1 (Less than once a week), 2 (Once or twice a week), 3 (Three or more times a week)<br>+<br>Breathing (30449) [0 (Not during the past month), 1 (Less than once a week), 2 (Once or twice a week), 3 (Three or more times a week)<br>+<br>Snore (30450) [0 (Not during the past month), 1 (Less than once a week), 2 (Once or twice a week), 3 (Three or more times a week)<br>+<br>Too cold (30451) [0 (Not during the past month), 1 (Less than once a week), 2 (Once or twice a week), 3 (Three or more times a week)<br>+ |

| | | | |
|---|---|---|---|
| | | | Too hot (30452) [0 (Not during the past month), 1 (Less than once a week), 2 (Once or twice a week), 3 (Three or more times a week)<br>+<br>Bad dreams (30453) [0 (Not during the past month), 1 (Less than once a week), 2 (Once or twice a week), 3 (Three or more times a week)<br>+<br>Pain (30454) [0 (Not during the past month), 1 (Less than once a week), 2 (Once or twice a week), 3 (Three or more times a week)<br>+<br>Other reasons (30461) [0 (Not during the past month), 1 (Less than once a week), 2 (Once or twice a week), 3 (Three or more times a week)<br><br><br>Sum of the above subscores: 0 (0), 1 (1-9), 2 (10-18), 3 (19-27)]<br><br>Component 6 - Use of sleep medication<br>Non-prescribed medications (30463) [0 (Not during the past month), 1 (Less than once a week), 2 (Once or twice a week), 3 (Three or more times a week)]<br>+<br>Prescripted medication (30464) [0 (Not during the past month), 1 (Less than once a week), 2 (Once or twice a week), 3 (Three or more times a week)]<br><br>Sum of the above subscores: 0 (0), 1 (1-2), 2 (3-4), 3 (5-6)<br><br>Component 7 - Daytime dysfunction<br>Stay awake during the day (30465) [0 (Not during the past month), 1 (Less than once a week), 2 (Once or twice a week), 3 (Three or more times a week)]<br>+<br>Enthusiasm (30466) [0 (No problem at all), 1 (Only a slight problem), 2 (Somewhat of a problem), 3 (A very big problem)]<br><br>Sum of the above subscores: 0 (0), 1 (1-2), 2 (3-4), 3 (5-6) |
| | **Sleep health dimension** | Satisfaction | Sleep quality (30467) [(Very good), (Fairly good), (Fairly bad), (Very bad)] |
|  |  | Timing | To calculate midpoint of sleep (MSF), find the midpoint of clock time between sleep onset and wake time:<br><br>sleep duration = wake time - (bedtime + SOL)<br>If sleep duration < 0: sleep duration += 24 hours<br>MSF = wake time - sleep duration /2<br>If MSF < 0: MSF += 24 hours<br><br>Exclude all the participants whose sleep duration is < 3 hours or > 16 hours |

| | | | |
|---|---|---|---|
| | | | Below are the relevant items for calculating MSF:<br>Bedtime (30442)<br>SOL (30443)<br>Wake time (30444) |
| | | Efficiency | To calculate sleep efficiency (SE), calculate the percentage of time asleep in bed from the total time in bed:<br><br>Sleep effiency = (Hours slept / Hours in bed) x 100<br><br>Below are the relevant items for calculating SE:<br>Hours slept = TST (30445)<br>Hours in bed = Wake time (30444) - Bedtime (30442)<br><br>If bedtime is before midnight, Hours in bed = Wake time - (Bedtime -24)<br>If bedtime is after midnight, Hours in bed = Wake time - Bedtime<br><br>1. For SE over 100%: round down people's SE if they are within 110% (basically allowing them mis-remember up to one hour assuming a 7.5 hour sleep duration)<br>2. Exclude all the outliers greater than 110%<br>3. Exlcude all the participants whose time in bed is < 3 hours or > 16 hours |
| | | Duration | TST (30445) [short (< 7 hours), optimal (7-9 hours), long (> 9 hours)] |
| **Potential correlates of sleep disruption** | | | |

| | | | |
|---|---|---|---|
| | **Chronotype** | | Chronotype is defined by the total score of the items below (range 4-25) and categorised into chronotype:<br><br>**Scoring per questions:**<br><br>[Get up time (30425) = 5 (5:00–6:30am), 4 (6:30–7:45am), 3 (7:45–9:45am), 2 (9:45–11:00am), 1 (11:00–12 noon)]<br>+<br>[Feeling after waking (30426) = 1 (Very tired), 2 (Fairly tired), 3 (Fairly refreshed), 4 (Very refreshed)]<br>+<br>[Sleep time (30427) = 5 (8:00–9:00pm), 4 (9:00–10:15pm), 3 (10:15 pm–12:45am), 2 (12:45–2:00am), 1 (2:00–3:00am)]<br>+<br>[Feeling your best (30428) = 5 (5:00–8:00am), 4 (8:00–10:00am), 3 (10:00 am–5:00pm), 2 (5:00–10:00pm), 1 (10:00 PM–5:00am)]<br>+<br>[Chronotype (30429) = 6 (Definitely a morning-type), 4 (Rather more a morning-type than an evening-type), 2 (Rather more an evening-type than a morning-type), 0 (Definitely an evening-type)]<br><br>**Overall scoring:**<br><br>Definitely morning type: 22-25<br>Moderately morning type: 18-21<br>Neither type: 12-17<br>Moderately evening type: 8-11<br>Definitely evening type: 4-7 |

| | | | |
|---|---|---|---|
| | **Family history of sleep disorders** | | Descriptive summary:<br><br>**Family history of insomnia**<br>[Insomnia (32111) = (Yes)]<br><br>**Family history of sleep apnea**<br>[Sleep apnea (32112) = (Yes)]<br><br>**Family history of restless leg syndrome**<br>[Restless leg syndrome (32114) = (Yes)]<br><br>**Family history of sleep walking**<br>[Sleep walking (32115) = (Yes)]<br><br>**Family history of night terror**<br>[Night terrors (32116) = (Yes)] |

**Appendix 4:** Suggested approach to dealing with uncertain responses and missing data.

To retain as many responses as possible (and therefore cases and non-cases for each phenotype), we applied the following principles when dealing with missing data or ambiguous responses:

- All "**prefer not to answer**" responses were set to missing to respect participants' preference.
- The "**varies significantly**" response option was used in questions asking for clock times (e.g. what time did you wake up?). When the outcomes **required precise clock timings** (e.g. calculating sleep efficiency, mid-point of sleep period), we set all "varies significantly" responses to missing.
- **"Do not know" responses for questions probing experiences during the wake period** (e.g. uncomfortable sensations in your legs), or in relation to questions **involving other people *telling* them something** (e.g. have you ever been told…) could be interpreted as meaning they have rarely or never experienced such things, or that others have not told them they engage in that behaviour. It may, therefore, be appropriate to code a "do not know" response as "never" or "rarely or never" in these instances. However, in questions relating to **other people observing** (e.g. has anyone noticed…) or **being impacted by participant behaviour** (e.g. has your snoring ever bothered other people), it may be more appropriate to treat a "do not know" response as missing given the potential for ambiguity.
- A "**not applicable**" response for questions related to a **specific behaviour** (e.g. acting out your dreams while asleep) or in relation to questions involving **other people** (e.g. 'your snoring bothered other people', or 'others noticed that you stop breathing during sleep') could be interpreted as meaning the symptom, behaviour, or experience was not relevant to a participant. It may, therefore, be appropriate to code a "not applicable" response as "never" or "rarely or never".
- However, a "**not applicable**" response for questions related to a **specific activity** (e.g. 30569 - Have you ever nodded off or fallen asleep while driving a vehicle) could be interpreted as meaning they do not engage in the activity (i.e. driving) or it could mean that they have never fallen asleep while driving. Given the ambiguity, a "not applicable" response to such questions were treated as missing.
- A "do not know" response in **frequency-based responses** were set to missing.
- In the **chronotype** question ("one hears about "morning-types" and "evening-types." Which one of these types do you consider yourself to be?"), there were four available responses (and their scoring for the chronotype composite score): definitely a

morning-type (6), rather more a morning-type than an evening-type (4), rather more an evening-type than a morning-type (2), definitely an evening-type (0). A “do not know” response to this question likely indicates that the individual does not endorse the extreme, and therefore has preference for neither morning nor evening. When calculating composite score for chronotype, we coded “do not know” as “3” to represent intermediate chronotype, consistent with a previously published study (Jones et al., 2019); this is the score in between “rather more a morning-type than an evening-type” and “rather more an evening-type than a morning-type”.

- Jones, S. E., Lane, J. M., Wood, A. R., van Hees, V. T., Tyrrell, J., Beaumont, R. N., … & Weedon, M. N. (2019). Genome-wide association analyses of chronotype in 697,828 individuals provides insights into circadian rhythms. Nature communications, 10(1), 343.

On the next page, we specify how missing data were dealt with and the assumptions made for each of the questionnaire items.

| Prefer not to answer | Varies significantly |
|---|---|
| • All questions<br> ○ **Code response as missing** | **Work and sleep**<br>• 30436 - On workdays in the past month, what time did you typically fall asleep? (This may be different to the time you went to bed)<br> ○ **Code response as missing**<br>• 30437 - On workdays in the past month, what time did you typically wake-up?<br> ○ **Code response as missing**<br>• 30438 - On non-working days in the past month, what time did you typically fall asleep?<br> ○ **Code response as missing**<br>• 30439 - On non-working days in the past month, what time did you typically fall asleep?<br> ○ **Code response as missing**<br>• 30440 - On non-working days in the past month, what time did you typically wake up?<br> ○ **Code response as missing**<br>• 30441 - On non-working days in the past month, what time did you typically wake up?<br> ○ **Code response as missing** |

| Do not know | Not applicable |
|---|---|
| **MEQ**<br>• 30429 - One hears about "morning-types" and "evening-types." Which one of these types do you consider yourself to be?<br> ○ Definitely a morning-type<br> ○ Rather more a morning-type than an evening-type<br> ○ Rather more an evening-type than a morning-type<br> ○ Definitely an evening-type<br> ○ **Code response as "3"**<br> ▪ **This is the score in between "rather more a morning-type than an evening-type" and "rather more an evening-type than a morning-type"** |  |
| **Parasomnia**<br>• 30555 - **How often** do you walk in your sleep?<br> ○ Not in the past year<br> ○ 1-2 times per year<br> ○ 1-2 times per month<br> ○ 1-2 times per week | **Parasomnia**<br>• 30557 - Have you ever been told, or suspected yourself, that you seem to 'act out your dreams' while asleep (for example, flailing your arms in the air, making running movements, etc.)?<br> ○ Yes<br> ○ No |

| | |
|---|---|
| ○ 3-4 times per week<br>○ Almost every day<br>○ Every day<br>○ **Code response as missing**<br><br>• 30557 - Have **you ever been told**, or suspected yourself, that you seem to 'act out your dreams' while asleep (for example, flailing your arms in the air, making running movements, etc.)?<br>○ Yes<br>○ No<br>○ **Code response as "No"**<br><br>• 30558 - **How often** do you have violent or injurious behaviour during sleep (for example, punching, kicking, leaping or running away from the bed)?<br>○ Never<br>○ Not in the past year<br>○ 1-2 times per year<br>○ 1-2 times per month<br>○ 1-2 times per week<br>○ 3-4 times per week<br>○ Almost every day<br>○ Every day<br>○ **Code response as missing**<br><br>• 30559 - **How often** do you have nightmares (frightening dreams)?<br>○ Never<br>○ Not in the past year<br>○ 1-2 times per year<br>○ 1-2 times per month<br>○ 1-2 times per week<br>○ 3-4 times per week<br>○ Almost every day<br>○ Every day<br>○ **Code response as missing** | ○ **Code response as "No"** |
| **Apnea**<br>• 30562 - Do you snore? | **Apnea**<br>• 30565 - Has your snoring ever **bothered other people**? |

| | |
|---|---|
| ◦ Yes<br>◦ No<br>◦ **Code response as missing**<br><br>• 30563 - Your snoring is:<br>◦ Slightly louder than breathing<br>◦ As loud as talking<br>◦ Louder than talking<br>◦ **Code response as missing**<br><br>• 30564 - **How often** do you snore?<br>◦ Almost every day<br>◦ 3-4 times per week<br>◦ 1-2 times per week<br>◦ 1-2 times per month<br>◦ Rarely or never<br>◦ **Code response as missing**<br><br>• 30565 - Has your snoring ever **bothered other people**?<br>◦ Yes<br>◦ No<br>◦ **Code response as missing**<br><br>• 30566 - **Has anyone noticed** that you stop breathing during your sleep?<br>◦ Almost every day<br>◦ 3-4 times per week<br>◦ 1-2 times per week<br>◦ 1-2 times per month<br>◦ Rarely or never<br>◦ **Code response as missing**<br><br>• 30567 - **How often** do you feel tired or fatigued after your sleep?<br>◦ Almost every day<br>◦ 3-4 times per week<br>◦ 1-2 times per week<br>◦ 1-2 times per month | ◦ Yes<br>◦ No<br>◦ **Code response as “No”**<br><br>• 30566 - **Has anyone noticed** that you stop breathing during your sleep?<br>◦ Almost every day<br>◦ 3-4 times per week<br>◦ 1-2 times per week<br>◦ 1-2 times per month<br>◦ Rarely or never<br>◦ **Code response as “Rarely or never”**<br><br>• 30569 - Have you ever nodded off or fallen asleep while driving a vehicle?<br>◦ Yes<br>◦ No<br>◦ **Code response as missing** |

| |
|---|
| - Code response as missing<br><br>• 30571 - Thinking about the last year, **how many** times have you had an accident (motor vehicle, home or work related) or a near miss due to sleepiness?<br> - More than 10 times<br> - 5-10 times<br> - 2-5 times<br> - Once<br> - **Code response as missing** |
| <u>**Restless legs**</u><br>• 30579 - Do you have recurrent uncomfortable feelings or sensations in your legs while you are sitting or lying down?<br> - Yes<br> - No<br> - **Code response as "No"**<br><br>• 30580 - Do you have a recurrent need or urge to move your legs while you are sitting or lying down?<br> - Yes<br> - No<br> - **Code response as "No"**<br><br>• 30581 - Are you **more likely** to have these feelings in your legs when you are resting (either sitting or lying down) or when you are physically active?<br> - Resting<br> - Active<br> - **Code response as missing**<br><br>• 30582 - If you get up or move around when you have these feelings in your legs, do these feelings get any better while you actually keep moving?<br> - Yes<br> - No<br> - **Code response as missing**<br><br>• 30583 - **Which time**(s) of day are these feelings in your legs most likely to occur? |

- Morning
- Mid-day
- Afternoon
- Evening
- Night
- About equal at all times
- **Code response as missing**

- 30584 - Will simply changing leg position once without continuing to move usually relieve these feelings in your legs?
  - Usually relieves
  - Does not usually relieve
  - **Code response as missing**

- 30585 - Are these feelings in your legs always due to muscle cramps?
  - Yes
  - No
  - **Code response as missing**

- 32070 - In the past 12 months, **how often** did you experience these feelings in your legs?
  - 6-7 days per week
  - 4-5 days per week
  - 2-3 days per week
  - 1 day per week
  - 2 days per month
  - 1 day per month or less
  - **Code response as missing**

**Family history**

- Have any close BLOOD relatives (including brother/sister, mother/father, son/daughter) had any of the following?
- 32111 - Insomnia (difficulty falling asleep or staying asleep)
  - Yes
  - No
  - **Code response as missing**

| | |
|---|---|
| • 32112 - Sleep apnoea (breathing pauses during sleep)<br>○ Yes<br>○ No<br>○ **Code response as missing**<br><br>• 32113 - Narcolepsy (difficulty staying awake or having “sleep attacks”)<br>○ Yes<br>○ No<br>○ **Code response as missing**<br><br>• 32114 - Restless leg syndrome (RLS) (uncontrollable urge to move the legs)<br>○ Yes<br>○ No<br>○ **Code response as missing**<br><br>• 32115 - Sleep walking<br>○ Yes<br>○ No<br>○ **Code response as missing**<br><br>• 32116 - Night terrors (partial waking from sleep with behaviours such as screaming, kicking, panic, sleep walking, thrashing or mumbling)<br>○ Yes<br>○ No<br>○ **Code response as missing** | |
| **Lifestyle**<br>• 30482 - In a typical day in summer, **how many hours** do you spend outdoors?<br>○ Number OR<br>○ Less than an hour a day<br>○ **Code response as missing**<br><br>• 30483 - In a typical day in winter, **how many hours** do you spend outdoors?<br>○ Number OR<br>○ Less than an hour a day<br>○ **Code response as missing** | **Lifestyle**<br>• 30479 - **How long** before bedtime do you last use a computer, tablet, mobile phone or television?<br>○ I use them in bed<br>○ Less than 1 hour<br>○ 1-2 hours<br>○ 2-3 hours<br>○ 3 hours or longer<br>○ **Code response as missing** |

**Appendix 5:** Dealing with time-related issues

When using sleep timing variables with responses encoded in timestamps e.g. 8am and 9pm, we performed a minimal set of quality checks to ensure that computed values were within a sensible range, instead of error in the timestamp response. For computed time in bed, which require timestamp inputs, we manually removed values that were less than 3 or greater than 16 hours. This quality check influenced not only duration variables but also sleep midpoint and sleep efficiency calculations.

**Appendix 6**: Data fields for all single-item variables

**Table 3.** Data field for all single-item variables

| **Variable** | **Data Field** | **Modelling** |
|---|---|---|
| **From baseline questionnaire (2006-2010)** | | |
| Age at baseline | 34, 52, 53 | Continuous |
| Age at survey completion | 34, 52, 30489, 30491, 30493, 32120, 32122, 32124, 32126, 32128, 32129, 32132, 32134, | Continuous |
| Sex | 31 | Female, Male |
| Ethnicity | 21000 | Asian, Black, White, Other |
| Highest qualification | 6138 | None of the above, National exams at ages 16-18 years (A levels/AS levels or equivalent,<br>O levels/GCSEs or equivalent, CSEs or equivalent), Vocational qualifications (NVQ or HND<br>or HNC or equivalent, Other professional qualifications eg: nursing, teaching), College or<br>University degree |
| Current employment status | 6142 | Yes, No |
| Townsend Deprivation Score | 22189 | Quintiles by 2011 UK census |
| BMI | 21001 | Continuous |
| Self-rated health | 2178 | Poor, Fair, Good, Excellent |
| Smoking status | 20116 | Never-smoker, Ex-smoker, Current smoker |
| Alcohol consumption | 1558 | <1, 1-2, 3-4 times/week, Daily drinker |
| Prescribed sleep medications | 20003 | Yes, No |
| Sleep duration | 1160 | Continuous |
| Insomnia | 1200 | Table 14 |

| | | |
|---|---|---|
| Daytime sleepiness | 1220 | Table 15 |
| Chronotype | 1180 | Table 16 |
| Shiftwork | 826, 3426 | Table 17 |
| Getting up in the morning | 1170 | Not at all easy, Not very easy, Fairly easy, Very easy |
| Snoring | 1210 | Yes, No |
| **From sleep questionnaire (2023)** | | |
| Types of shiftwork | 30430, 30431 | Normal hours (9am-5pm), evening shifts (work typically finished between 20:00-00:00), morning shifts (work typically started between 04:00-07:00), night shifts (20:00-08:00), rotating shifts or irregular work hours, do not work |
| Time of falling asleep on workdays | 30436 | Clock time |
| Time of waking up on workdays | 30437 | Clock time |
| Time of falling asleep on non-workdays | 30438, 30439 | Clock time |
| Time of waking up on non-workdays | 30440, 30441 | Clock time |
| Time spent outdoors in summer | 30482 | Number of hours, Less than 1 hour a day |
| Time spent outdoors in winter | 30483 | Number of hours, Less than 1 hour a day |
| Use of electronic device before bedtime | 30479 | I use them in bed, Less than 1 hour, 1-2 hours, 2-3 hours, 3 hours or longer |
| Exercise - frequency | 30477 | Daily, More than once a week, 3-4 times, 1-2 times, Not at all |
| Nap - frequency | 30475 | Daily, More than once a week, 3-4 times, 1-2 times, Not at all |
| Consumed alcohol to fall asleep - frequency | 32119 | Daily, More than once a week, 3-4 times, 1-2 times, Not at all |

| | | |
|---|---|---|
| Caffeine consumption | 30480 | Number of servings, I rarely or never drink caffeine |
| Accident due to sleepiness - frequency | 30571 | More than 10 times, 5-10 times, 2-5 times, Once, Never had an accident or near miss due to sleepiness |

**Appendix 7:** STROBE guideline tables

| | Item No | Recommendation | Page |
|---|---|---|---|
| **Title and abstract** | 1 | (*a*) Indicate the study's design with a commonly used term in the title or the abstract | 1 |
| | | (*b*) Provide in the abstract an informative and balanced summary of what was done and what was found | 2 |
| **Introduction** | | | |
| Background/rationale | 2 | Explain the scientific background and rationale for the investigation being reported | 4, 5 |
| Objectives | 3 | State specific objectives, including any prespecified hypotheses | 5, 6 |
| **Methods** | | | |
| Study design | 4 | Present key elements of study design early in the paper | 7 |
| Setting | 5 | Describe the setting, locations, and relevant dates, including periods of recruitment, exposure, follow-up, and data collection | 7-9 |
| Participants | 6 | (*a*) Give the eligibility criteria, and the sources and methods of selection of participants | 8 |
| Variables | 7 | Clearly define all outcomes, exposures, predictors, potential confounders, and effect modifiers. Give diagnostic criteria, if applicable | 8-10 |
| Data sources/ measurement | 8* | For each variable of interest, give sources of data and details of methods of assessment (measurement). Describe comparability of assessment methods if there is more than one group | 8-10 |
| Bias | 9 | Describe any efforts to address potential sources of bias | 10,11 |
| Study size | 10 | Explain how the study size was arrived at | 8 |
| Quantitative variables | 11 | Explain how quantitative variables were handled in the analyses. If applicable, describe which groupings were chosen and why | 9, 10 |
| Statistical methods | 12 | (*a*) Describe all statistical methods, including those used to control for confounding | / |
| | | (*b*) Describe any methods used to examine subgroups and interactions | / |
| | | (*c*) Explain how missing data were addressed | 10 |

| | | | |
|---|---|---|---|
| | | (*d*) If applicable, describe analytical methods taking account of sampling strategy | / |
| | | (*e*) Describe any sensitivity analyses | / |
| **Results** | | | |
| Participants | 13* | (a) Report numbers of individuals at each stage of study—eg numbers potentially eligible, examined for eligibility, confirmed eligible, included in the study, completing follow-up, and analysed | 12 |
| | | (b) Give reasons for non-participation at each stage | 12 |
| | | (c) Consider use of a flow diagram | 23 |
| Descriptive data | 14* | (a) Give characteristics of study participants (eg demographic, clinical, social) and information on exposures and potential confounders | 12 |
| | | (b) Indicate number of participants with missing data for each variable of interest | 12, in tables |
| Outcome data | 15* | Report numbers of outcome events or summary measures | In tables |
| Main results | 16 | (*a*) Give unadjusted estimates and, if applicable, confounder-adjusted estimates and their precision (eg, 95% confidence interval). Make clear which confounders were adjusted for and why they were included | 12-16, in tables |
| | | (*b*) Report category boundaries when continuous variables were categorized | 12-16, in tables |
| | | (*c*) If relevant, consider translating estimates of relative risk into absolute risk for a meaningful time period | / |
| Other analyses | 17 | Report other analyses done—eg analyses of subgroups and interactions, and sensitivity analyses | / |
| **Discussion** | | | |
| Key results | 18 | Summarise key results with reference to study objectives | 17 |
| Limitations | 19 | Discuss limitations of the study, taking into account sources of potential bias or imprecision. Discuss both direction and magnitude of any potential bias | 17, 18 |
| Interpretation | 20 | Give a cautious overall interpretation of results considering objectives, limitations, multiplicity of analyses, results from similar studies, and other relevant evidence | 17, 18 |
| Generalisability | 21 | Discuss the generalisability (external validity) of the study results | 18 |

| | | | |
|---|---|---|---|
| **Other information** | | | |
| Funding | 22 | Give the source of funding and the role of the funders for the present study and, if applicable, for the original study on which the present article is based | 20 |

**Appendix 8:** eTables

**eTable 1.** Sleep health at baseline (2006-2010) for UK Biobank Sleep Cohort: participant count and proportions by subgroup

| | **Baseline assessment** (n = 502,128) | **Sleep questionnaire** (n = 185,056) | **Invited but did not participate** (n = 144,047) |
|---|---|---|---|
| **Sleep duration** | | | |
| <7 hours | 123,147 (24.5%) | 41,291 (22.3%) | 34,931 (24.2%) |
| 7-8.9 hours | 336,445 (67.0%) | 132,297 (71.5%) | 98,083 (68.1%) |
| 9+ hours | 38,325 (7.6%) | 10,969 (5.9%) | 10,145 (7.0%) |
| Missing | 4,211 (0.8%) | 499 (0.3%) | 888 (0.6%) |
| **Chronotype** | | | |
| Definitely morning type | 120,267 (24.0%) | 41,748 (22.6%) | 32,876 (22.8%) |
| Intermediate type | 283,532 (56.5%) | 107,874 (58.3%) | 83,492 (58.0%) |
| Definitely evening type | 40,061 (8.0%) | 15,408 (8.3%) | 11,645 (8.1%) |
| Missing | 58,268 (11.6%) | 20,026 (10.8%) | 16,034 (11.1%) |
| **Insomnia** | | | |
| With frequent insomnia | 141,290 (28.1%) | 49,265 (26.6%) | 38,831 (27.0%) |
| Without frequent insomnia | 359,335 (71.6%) | 135,602 (73.3%) | 104,915 (72.8%) |
| Missing | 1,503 (0.3%) | 189 (0.1%) | 301 (0.2%) |
| **Daytime sleepiness** | | | |
| With frequent sleepiness | 14,088 (2.8%) | 3,950 (2.1%) | 3,624 (2.5%) |
| Without frequent sleepiness | 484,307 (96.5%) | 180,747 (97.7%) | 139,643 (96.9%) |
| Missing | 3,733 (0.7%) | 359 (0.2%) | 780 (0.5%) |
| **Shift work** | | | |
| Day work | 236,664 (47.1%) | 106,086 (57.3%) | 70,720 (49.1%) |
| Shift work without night shifts | 24,200 (4.8%) | 8,655 (4.7%) | 7,172 (5.0%) |
| Shift work with night shifts | 18,111 (3.6%) | 6,178 (3.3%) | 5,717 (4.0%) |
| Shift work with unknown schedule | 77 (0.0%) | 11 (0.0%) | 24 (0.0%) |
| Permanent night shift work | 7,117 (1.4%) | 2,153 (1.2%) | 2,134 (1.5%) |
| Missing | 215,959 (43.0%) | 61,973 (33.5%) | 58,280 (40.5%) |
| **Difficulty getting up in the morning** | | | |
| Very easy | 160,384 (31.9%) | 56,316 (30.4%) | 43,915 (30.5%) |
| Fairly easy | 246,463 (49.1%) | 94,567 (51.1%) | 72,838 (50.6%) |
| Not very easy | 69,874 (13.9%) | 26,423 (14.3%) | 20,740 (14.4%) |
| Not at all easy | 19,739 (3.9%) | 6,357 (3.4%) | 5,415 (3.8%) |
| Missing | 5,668 (1.1%) | 1,393 (0.8%) | 1,139 (0.8%) |
| **Snoring** | | | |
| Yes | 173,262 (34.5%) | 61,998 (33.5%) | 52,274 (36.3%) |
| No | 291,846 (58.1%) | 111,407 (60.2%) | 82,689 (57.4%) |
| Missing | 37,020 (7.4%) | 11,651 (6.3%) | 9,084 (6.3%) |

**eTable 2.** Sleep health dimensions stratified by sociodemographic factors at baseline (2006-2010)

| | | Sleep quality, mean (SD) | Satisfaction, n (%) | | | | Sleepiness, n (%) | | | | Timing, mean (SD) | Efficiency, mean (SD) | Sleep duration, n (%) | | | |
|---|---|---|---|---|---|---|---|---|---|---|---|---|---|---|---|---|
| | **Overall** | **PSQI** | **Very good** | **Fairly good** | **Fairly bad** | **Very bad** | **No chance** | **Slight chance** | **Moderate chance** | **High chance** | Sleep midpoint | Sleep efficiency (%) | Mean duration (SD) | Short (<7 hours) | Optimal (7-9 hours) | Long (>9 hours) |
| | (n = 185,056) | (n = 159,430) | (n = 36,216) | (n = 101,744) | (n = 37,390) | (n = 6,533) | (n = 72,703) | (n = 69,103) | (n = 25,843) | (n = 11,548) | (n = 165,692) | (n = 166,693) | | (n = 73,147) | (n = 99,086) | (n = 8,435) |
| **Overall** | | 6.65 (3.46) | 36,216 (19.6%) | 101,744 (55.0%) | 37,390 (20.2%) | 6,533 (3.5%) | 72,703 (39.3%) | 69,103 (37.3%) | 25,843 (14.0%) | 11,548 (6.2%) | 03:32 (75 min) | 79.1 (13.3) | 6.9 (1.2) | 73,147 (39.5%) | 99,086 (53.5%) | 8,435 (4.6%) |
| **Age at survey (years)** | | | | | | | | | | | | | | | | |
| <60 | 23,565 (12.7%) | 6.6 (3.5) | 3,575 (9.9%) | 12,745 (12.5%) | 5,680 (15.2%) | 1,093 (16.7%) | 10,813 (14.9%) | 8,093 (11.7%) | 2,404 (9.3%) | 1,244 (10.8%) | 03:11 (73 min) | 81.5 (12.9) | 6.83 (1.1) | 9,837 (13.4%) | 12,410 (12.5%) | 732 (8.7%) |
| 60-69 | 64,276 (34.7%) | 6.6 (3.5) | 11,743 (32.4%) | 35,503 (34.9%) | 13,685 (36.6%) | 2,445 (37.4%) | 27,684 (38.1%) | 23,685 (34.3%) | 7,610 (29.4%) | 3,523 (30.5%) | 03:29 (74 min) | 79.6 (13.2) | 6.94 (1.2) | 25,000 (34.2%) | 35,194 (35.5%) | 2,873 (34.1%) |
| 70-79 | 81,121 (43.8%) | 6.7 (3.4) | 17,037 (47.0%) | 44,843 (44.1%) | 15,356 (41.1%) | 2,524 (38.6%) | 29,493 (40.6%) | 31,318 (45.3%) | 12,608 (48.8%) | 5,370 (46.5%) | 03:39 (75 min) | 78.2 (13.3) | 6.94 (1.2) | 31,920 (43.6%) | 43,356 (43.8%) | 3,882 (46.0%) |
| 80+ | 16,094 (8.7%) | 6.8 (3.5) | 3,861 (10.7%) | 8,653 (8.5%) | 2,669 (7.1%) | 471 (7.2%) | 4,713 (6.5%) | 6,007 (8.7%) | 3,221 (12.5%) | 1,411 (12.2%) | 03:40 (73 min) | 77.9 (13.6) | 6.95 (1.2) | 6,390 (8.7%) | 8,126 (8.2%) | 948 (11.2%) |
| **Sex** | | | | | | | | | | | | | | | | |
| Female | 107,071 (57.9%) | 7.1 (3.5) | 18,971 (52.4%) | 58,025 (57.0%) | 23,903 (63.9%) | 4,336 (66.4%) | 44,699 (61.5%) | 39,090 (56.6%) | 13,627 (52.7%) | 6,268 (54.3%) | 03:34 (74 min) | 77.4 (13.4) | 6.87 (1.2) | 44,753 (61.2%) | 55,060 (55.6%) | 4,626 (54.8%) |
| Male | 77,985 (42.1%) | 6.0 (3.2) | 17,245 (47.6%) | 43,719 (43.0%) | 13,487 (36.1%) | 2,197 (33.6%) | 28,004 (38.5%) | 30,013 (43.4%) | 12,216 (47.3%) | 5,280 (45.7%) | 03:29 (76 min) | 81.4 (12.8) | 7.00 (1.2) | 28,394 (38.8%) | 44,026 (44.4%) | 3,809 (45.2%) |
| **Ethnicity** | | | | | | | | | | | | | | | | |
| Asian or Asian British | 2,063 (1.1%) | 6.7 (3.5) | 366 (1.0%) | 1,214 (1.2%) | 342 (0.9%) | 83 (1.3%) | 823 (1.1%) | 742 (1.1%) | 258 (1.0%) | 108 (0.9%) | 03:28 (88 min) | 80.9 (14.6) | 6.69 (1.3) | 990 (1.4%) | 914 (0.9%) | 80 (0.9%) |
| Black or Black British | 1,294 (0.7%) | 7.5 (3.9) | 230 (0.6%) | 660 (0.6%) | 253 (0.7%) | 90 (1.4%) | 425 (0.6%) | 455 (0.7%) | 204 (0.8%) | 91 (0.8%) | 03:38 (92 min) | 78.9 (15.6) | 6.33 (1.4) | 772 (1.1%) | 413 (0.4%) | 33 (0.4%) |
| Mixed Race or Other | 1,996 (1.1%) | 7.0 (3.7) | 352 (1.0%) | 1,042 (1.0%) | 437 (1.2%) | 113 (1.7%) | 809 (1.1%) | 681 (1.0%) | 267 (1.0%) | 131 (1.1%) | 03:35 (82 min) | 79.9 (14.5) | 6.67 (1.3) | 929 (1.3%) | 942 (1.0%) | 55 (0.7%) |
| White | 179,047 (96.8%) | 6.6 (3.5) | 35,138 (97.0%) | 98,476 (96.8%) | 36,220 (96.9%) | 6,222 (95.2%) | 70,392 (96.8%) | 67,001 (97.0%) | 25,001 (96.7%) | 11,174 (96.8%) | 03:32 (75 min) | 79.1 (13.2) | 6.93 (1.2) | 70,186 (96.0%) | 96,488 (97.4%) | 8,232 (97.6%) |
| Missing | 656 (0.4%) | 6.7 (3.5) | 130 (0.4%) | 352 (0.3%) | 138 (0.4%) | 25 (0.4%) | 254 (0.3%) | 224 (0.3%) | 113 (0.4%) | 44 (0.4%) | 03:35 (80 min) | 79.8 (14.1) | 6.90 (1.3) | 270 (0.4%) | 329 (0.3%) | 35 (0.4%) |
| **Highest qualification** | | | | | | | | | | | | | | | | |

| | | | | | | | | | | | | | | | | |
|---|---|---|---|---|---|---|---|---|---|---|---|---|---|---|---|---|
| College or University degree | 81,005 (43.8%) | 6.2 (3.3) | 16,772 (46.3%) | 45,481 (44.7%) | 15,517 (41.5%) | 2,100 (32.1%) | 32,825 (45.1%) | 30,949 (44.8%) | 10,654 (41.2%) | 4,483 (38.8%) | 03:30 (72 min) | 80.5 (12.3) | 7.01 (1.1) | 29,055 (39.7%) | 47,133 (47.6%) | 3,306 (39.2%) |
| Vocational qualifications | 50,578 (27.3%) | 6.9 (3.5) | 9,685 (26.7%) | 27,734 (27.3%) | 10,356 (27.7%) | 1,964 (30.1%) | 19,400 (26.7%) | 18,957 (27.4%) | 7,305 (28.3%) | 3,326 (28.8%) | 03:32 (74 min) | 78.5 (13.6) | 6.89 (1.2) | 20,898 (28.6%) | 26,035 (26.3%) | 2,460 (29.2%) |
| National exams at ages 16-18 | 38,692 (20.9%) | 7.0 (3.6) | 7,008 (19.4%) | 20,819 (20.5%) | 8,479 (22.7%) | 1,671 (25.6%) | 15,186 (20.9%) | 14,073 (20.4%) | 5,506 (21.3%) | 2,583 (22.4%) | 03:33 (77 min) | 77.8 (13.8) | 6.84 (1.2) | 16,558 (22.6%) | 19,320 (19.5%) | 1,808 (21.4%) |
| None of the above | 12,886 (7.0%) | 7.5 (3.8) | 2,417 (6.7%) | 6,697 (6.6%) | 2,634 (7.0%) | 712 (10.9%) | 4,538 (6.2%) | 4,460 (6.5%) | 2,123 (8.2%) | 1,021 (8.8%) | 03:39 (88 min) | 76.3 (15.0) | 6.77 (1.4) | 5,838 (8.0%) | 5,675 (5.7%) | 765 (9.1%) |
| Missing | 1,895 (1.0%) | 7.0 (3.5) | 334 (0.9%) | 1,013 (1.0%) | 404 (1.1%) | 86 (1.3%) | 754 (1.0%) | 664 (1.0%) | 255 (1.0%) | 135 (1.2%) | 03:40 (77 min) | 78.1 (14.1) | 6.85 (1.3) | 798 (1.1%) | 923 (0.9%) | 96 (1.1%) |
| **Townsend deprivation index** | | | | | | | | | | | | | | | | |
| Least Deprived | 76,751 (41.5%) | 6.5 (3.4) | 15,742 (43.5%) | 42,616 (41.9%) | 14,958 (40.0%) | 2,277 (34.9%) | 29,922 (41.2%) | 29,319 (42.4%) | 10,774 (41.7%) | 4,599 (39.8%) | 03:31 (72 min) | 79.1 (12.9) | 6.97 (1.1) | 29,116 (39.8%) | 42,502 (42.9%) | 3,540 (42.0%) |
| 2nd Quintile | 39,309 (21.2%) | 6.6 (3.4) | 7,828 (21.6%) | 21,836 (21.5%) | 7,737 (20.7%) | 1,275 (19.5%) | 15,449 (21.2%) | 14,699 (21.3%) | 5,505 (21.3%) | 2,465 (21.3%) | 03:31 (75 min) | 79.1 (13.2) | 6.94 (1.2) | 15,447 (21.1%) | 21,164 (21.4%) | 1,827 (21.7%) |
| 3rd Quintile | 27,699 (15.0%) | 6.7 (3.5) | 5,277 (14.6%) | 15,150 (14.9%) | 5,771 (15.4%) | 1,022 (15.6%) | 10,958 (15.1%) | 10,255 (14.8%) | 3,840 (14.9%) | 1,763 (15.3%) | 03:32 (75 min) | 79.2 (13.2) | 6.91 (1.2) | 11,110 (15.2%) | 14,674 (14.8%) | 1,239 (14.7%) |
| 4th Quintile | 22,441 (12.1%) | 6.8 (3.6) | 4,119 (11.4%) | 12,223 (12.0%) | 4,743 (12.7%) | 947 (14.5%) | 8,997 (12.4%) | 8,156 (11.8%) | 3,087 (11.9%) | 1,392 (12.1%) | 03:33 (77 min) | 79.1 (13.8) | 6.87 (1.2) | 9,241 (12.6%) | 11,587 (11.7%) | 1,032 (12.2%) |
| Most Deprived | 18,637 (10.1%) | 7.1 (3.7) | 3,220 (8.9%) | 9,808 (9.6%) | 4,113 (11.0%) | 1,004 (15.4%) | 7,298 (10.0%) | 6,591 (9.5%) | 2,607 (10.1%) | 1,308 (11.3%) | 03:36 (86 min) | 78.9 (14.3) | 6.78 (1.3) | 8,125 (11.1%) | 9,061 (9.1%) | 786 (9.3%) |
| Missing | 219 (0.1%) | 7.4 (3.5) | 30 (0.1%) | 111 (0.1%) | 68 (0.2%) | 8 (0.1%) | 79 (0.1%) | 83 (0.1%) | 30 (0.1%) | 21 (0.2%) | 03:34 (75 min) | 77.9 (13.9) | 6.82 (1.2) | 108 (0.1%) | 98 (0.1%) | 11 (0.1%) |
| **Employment status** | | | | | | | | | | | | | | | | |
| Employed | 123,177 (66.6%) | 6.5 (3.4) | 23,639 (65.3%) | 68,382 (67.2%) | 25,102 (67.1%) | 4,160 (63.7%) | 50,715 (69.8%) | 46,313 (67.0%) | 15,617 (60.4%) | 6,856 (59.4%) | 03:28 (74 min) | 79.8 (13.0) | 6.93 (1.1) | 48,109 (65.8%) | 67,356 (68.0%) | 5,188 (61.5%) |
| Not employed | 61,453 (33.2%) | 7.0 (3.5) | 12,501 (34.5%) | 33,159 (32.6%) | 12,187 (32.6%) | 2,347 (35.9%) | 21,825 (30.0%) | 22,631 (32.7%) | 10,178 (39.4%) | 4,663 (40.4%) | 03:39 (76 min) | 77.7 (13.7) | 6.91 (1.2) | 24,851 (34.0%) | 31,541 (31.8%) | 3,216 (38.1%) |
| Missing | 426 (0.2%) | 7.3 (3.8) | 76 (0.2%) | 203 (0.2%) | 101 (0.3%) | 26 (0.4%) | 163 (0.2%) | 159 (0.2%) | 48 (0.2%) | 29 (0.3%) | 03:39 (78 min) | 78.4 (14.9) | 6.85 (1.4) | 187 (0.3%) | 189 (0.2%) | 31 (0.4%) |
| **Body mass index** | | | | | | | | | | | | | | | | |
| Underweight | 1,005 (0.5%) | 6.2 (3.3) | 206 (0.6%) | 558 (0.5%) | 198 (0.5%) | 32 (0.5%) | 458 (0.6%) | 363 (0.5%) | 110 (0.4%) | 49 (0.4%) | 03:20 (83 min) | 80.4 (12.5) | 6.93 (1.1) | 378 (0.5%) | 570 (0.6%) | 36 (0.4%) |
| Normal weight | 70,668 (38.2%) | 6.3 (3.3) | 14,421 (39.8%) | 39,538 (38.9%) | 13,635 (36.5%) | 2,029 (31.1%) | 30,211 (41.6%) | 26,144 (37.8%) | 8,681 (33.6%) | 3,683 (31.9%) | 03:27 (72 min) | 79.7 (12.7) | 6.97 (1.1) | 26,341 (36.0%) | 40,009 (40.4%) | 2,861 (33.9%) |
| Overweight | 76,487 (41.3%) | 6.6 (3.4) | 15,400 (42.5%) | 42,315 (41.6%) | 14,919 (39.9%) | 2,507 (38.4%) | 29,338 (40.4%) | 28,885 (41.8%) | 11,008 (42.6%) | 4,807 (41.6%) | 03:32 (75 min) | 79.2 (13.2) | 6.94 (1.2) | 29,794 (40.7%) | 41,252 (41.6%) | 3,561 (42.2%) |
| Obese | 36,454 (19.7%) | 7.4 (3.7) | 6,106 (16.9%) | 19,096 (18.8%) | 8,551 (22.9%) | 1,942 (29.7%) | 12,527 (17.2%) | 13,566 (19.6%) | 5,980 (23.1%) | 2,973 (25.7%) | 03:42 (80 min) | 77.5 (14.4) | 6.79 (1.3) | 16,431 (22.5%) | 17,049 (17.2%) | 1,956 (23.2%) |
| Missing | 442 (0.2%) | 7.0 (3.6) | 83 (0.2%) | 237 (0.2%) | 87 (0.2%) | 23 (0.4%) | 169 (0.2%) | 145 (0.2%) | 64 (0.2%) | 36 (0.3%) | 03:42 (83 min) | 78.8 (14.2) | 6.80 (1.3) | 203 (0.3%) | 206 (0.2%) | 21 (0.2%) |

| | | | | | | | | | | | | | | | | |
|---|---|---|---|---|---|---|---|---|---|---|---|---|---|---|---|---|
| **Smoking status** | | | | | | | | | | | | | | | | |
| Never | 108,409 (58.6%) | 6.5 (3.4) | 21,732 (60.0%) | 60,048 (59.0%) | 21,357 (57.1%) | 3,515 (53.8%) | 42,777 (58.8%) | 41,239 (59.7%) | 14,680 (56.8%) | 6,501 (56.3%) | 03:30 (74 min) | 79.3 (13.1) | 6.94 (1.2) | 42,043 (57.5%) | 59,107 (59.7%) | 4,779 (56.7%) |
| Previous | 63,168 (34.1%) | 6.8 (3.5) | 12,079 (33.4%) | 34,578 (34.0%) | 13,105 (35.0%) | 2,294 (35.1%) | 24,407 (33.6%) | 23,239 (33.6%) | 9,322 (36.1%) | 4,138 (35.8%) | 03:33 (75 min) | 78.6 (13.4) | 6.91 (1.2) | 25,469 (34.8%) | 33,252 (33.6%) | 2,943 (34.9%) |
| Current | 13,017 (7.0%) | 7.0 (3.7) | 2,334 (6.4%) | 6,857 (6.7%) | 2,843 (7.6%) | 696 (10.7%) | 5,344 (7.4%) | 4,466 (6.5%) | 1,774 (6.9%) | 875 (7.6%) | 03:40 (85 min) | 79.6 (14.2) | 6.86 (1.3) | 5,440 (7.4%) | 6,501 (6.6%) | 696 (8.3%) |
| Missing | 462 (0.2%) | 7.1 (3.8) | 71 (0.2%) | 261 (0.3%) | 85 (0.2%) | 28 (0.4%) | 175 (0.2%) | 159 (0.2%) | 67 (0.3%) | 34 (0.3%) | 03:32 (91 min) | 78.1 (14.7) | 6.84 (1.3) | 195 (0.3%) | 226 (0.2%) | 17 (0.2%) |
| **Alcohol consumption frequency** | | | | | | | | | | | | | | | | |
| <1 times/week | 47,328 (25.6%) | 7.1 (3.7) | 8,698 (24.0%) | 25,088 (24.7%) | 10,307 (27.6%) | 2,289 (35.0%) | 17,757 (24.4%) | 17,262 (25.0%) | 6,980 (27.0%) | 3,603 (31.2%) | 03:33 (81 min) | 78.4 (14.2) | 6.82 (1.3) | 20,635 (28.2%) | 23,058 (23.3%) | 2,264 (26.8%) |
| 1-2 times/week | 47,529 (25.7%) | 6.7 (3.5) | 9,261 (25.6%) | 26,211 (25.8%) | 9,647 (25.8%) | 1,639 (25.1%) | 18,575 (25.5%) | 17,890 (25.9%) | 6,695 (25.9%) | 2,872 (24.9%) | 03:31 (74 min) | 79.1 (13.2) | 6.92 (1.2) | 18,988 (26.0%) | 25,356 (25.6%) | 2,117 (25.1%) |
| 3-4 times/week | 48,711 (26.3%) | 6.4 (3.3) | 9,595 (26.5%) | 27,526 (27.1%) | 9,479 (25.4%) | 1,357 (20.8%) | 19,580 (26.9%) | 18,574 (26.9%) | 6,536 (25.3%) | 2,637 (22.8%) | 03:31 (72 min) | 79.4 (12.7) | 6.98 (1.1) | 18,087 (24.7%) | 27,596 (27.9%) | 2,021 (24.0%) |
| Daily | 41,334 (22.3%) | 6.4 (3.3) | 8,631 (23.8%) | 22,837 (22.4%) | 7,931 (21.2%) | 1,240 (19.0%) | 16,730 (23.0%) | 15,317 (22.2%) | 5,615 (21.7%) | 2,428 (21.0%) | 03:32 (73 min) | 79.5 (12.9) | 6.99 (1.1) | 15,374 (21.0%) | 23,005 (23.2%) | 2,022 (24.0%) |
| Missing | 154 (0.1%) | 6.7 (3.7) | 31 (0.1%) | 82 (0.1%) | 26 (0.1%) | 8 (0.1%) | 61 (0.1%) | 60 (0.1%) | 17 (0.1%) | 8 (0.1%) | 03:45 (97 min) | 79.9 (14.6) | 6.92 (1.4) | 63 (0.1%) | 71 (0.1%) | 11 (0.1%) |
| **Self-reported overall health rating** | | | | | | | | | | | | | | | | |
| Excellent | 40,056 (21.6%) | 5.5 (3.1) | 12,192 (33.7%) | 21,148 (20.8%) | 5,485 (14.7%) | 724 (11.1%) | 18,171 (25.0%) | 14,459 (20.9%) | 4,578 (17.7%) | 1,890 (16.4%) | 03:24 (68 min) | 81.4 (12.1) | 7.09 (1.1) | 13,300 (18.2%) | 24,265 (24.5%) | 1,806 (21.4%) |
| Good | 111,924 (60.5%) | 6.6 (3.3) | 20,829 (57.5%) | 64,223 (63.1%) | 21,883 (58.5%) | 3,136 (48.0%) | 43,475 (59.8%) | 43,033 (62.3%) | 15,493 (60.0%) | 6,442 (55.8%) | 03:31 (74 min) | 79.2 (13.0) | 6.94 (1.1) | 43,838 (59.9%) | 60,657 (61.2%) | 4,870 (57.7%) |
| Fair | 28,549 (15.4%) | 8.1 (3.7) | 2,852 (7.9%) | 14,589 (14.3%) | 8,481 (22.7%) | 1,954 (29.9%) | 9,630 (13.2%) | 10,273 (14.9%) | 4,908 (19.0%) | 2,559 (22.2%) | 03:41 (83 min) | 76.4 (14.7) | 6.71 (1.3) | 13,640 (18.6%) | 12,535 (12.7%) | 1,432 (17.0%) |
| Poor | 4,092 (2.2%) | 9.7 (4.1) | 273 (0.8%) | 1,573 (1.5%) | 1,438 (3.8%) | 691 (10.6%) | 1,283 (1.8%) | 1,180 (1.7%) | 794 (3.1%) | 615 (5.3%) | 03:57 (93 min) | 73.0 (16.7) | 6.54 (1.7) | 2,172 (3.0%) | 1,436 (1.4%) | 306 (3.6%) |
| Missing | 435 (0.2%) | 7.3 (3.8) | 70 (0.2%) | 211 (0.2%) | 103 (0.3%) | 28 (0.4%) | 144 (0.2%) | 158 (0.2%) | 70 (0.3%) | 42 (0.4%) | 03:42 (98 min) | 77.8 (15.1) | 6.75 (1.5) | 197 (0.3%) | 193 (0.2%) | 21 (0.2%) |
| **Self-reported prescribed sleep medication** | | | | | | | | | | | | | | | | |
| Yes | 1,167 (0.6%) | 10.2 (3.9) | 79 (0.2%) | 467 (0.5%) | 433 (1.2%) | 166 (2.5%) | 500 (0.7%) | 367 (0.5%) | 158 (0.6%) | 97 (0.8%) | 04:02 (87 min) | 71.7 (16.2) | 6.45 (1.4) | 643 (0.9%) | 446 (0.5%) | 46 (0.5%) |
| No | 183,814 (99.3%) | 6.6 (3.4) | 36,125 (99.7%) | 101,231 (99.5%) | 36,946 (98.8%) | 6,363 (97.4%) | 72,174 (99.3%) | 68,706 (99.4%) | 25,676 (99.4%) | 11,450 (99.2%) | 03:32 (75 min) | 79.1 (13.2) | 6.93 (1.2) | 72,475 (99.1%) | 98,601 (99.5%) | 8,384 (99.4%) |
| Missing | 75 (0.0%) | 6.4 (3.5) | 12 (0.0%) | 46 (0.0%) | 11 (0.0%) | 4 (0.1%) | 29 (0.0%) | 30 (0.0%) | 9 (0.0%) | 1 (0.0%) | 03:33 (87 min) | 80.6 (12.5) | 7.00 (1.3) | 29 (0.0%) | 39 (0.0%) | 5 (0.1%) |

**eTable 3.** Sleep timing on work days and non-work days

| | Mean (SD) | Typical hours (n = 35,632) | Evening shifts (n = 1,408) | Morning shifts (n = 2,113) | Night shifts (n = 607) | Rotating shifts or had irregular work hours (n = 6,780) | Do not work (n = 136,220) |
|---|---|---|---|---|---|---|---|
| **Workdays** | Time of falling asleep | 22:58 (77 min) | 23:55 (94 min) | 21:57 (105 min) | 06:27 (337 min) | 23:07 (100 min) | / |
| | Time of waking up | 06:39 (107 min) | 07:29 (123 min) | 05:15 (106 min) | 12:03 (310 min) | 06:41 (117 min) | / |
| **Non-workdays** | Time of falling asleep | 23:14 (85 min) | 23:44 (92 min) | 22:35 (92 min) | 23:23 (135 min) | 23:18 (96 min) | 23:14 (88 min) |
| | Time of waking up | 07:32 (111 min) | 08:01 (121 min) | 06:57 (111 min) | 07:56 (151 min) | 07:29 (114 min) | 07:19 (126 min) |

***Note*:** Typical hours (9am-5pm), evening shifts (work typically finished between 8pm and midnight), morning shifts (work typically started between 4am and 7am), night shifts (work typically took place between 8pm and 8am).

**eTable 4.** Socioeconomic variables at baseline (2006-2010) by operationally defined sleep disorders

| | **Overall** (n = 185,056) | **Chronic Insomnia Disorder** (n = 26,729) | **Obstructive Sleep Apnoea** (n = 14,796) | **Restless Legs Syndrome** (n = 7,506) | **Delayed Sleep-Wake Phase Disorder** (n = 1,207) | **Advanced Sleep-Wake Phase Disorder** (n = 903) | **Shift Work Disorder** (n = 1,717) | **Sleepwalking** (n = 1,609) | **REM Sleep Behaviour Disorder** (n = 2,157) | **Nightmare Disorder** (n = 6,805) |
|---|---|---|---|---|---|---|---|---|---|---|
| **Age at survey (years)** | | | | | | | | | | |
| <60 | 23,565 (12.7%) | 4,278 (16.0%) | 2,881 (19.5%) | 971 (12.9%) | 241 (20.0%) | 118 (13.1%) | 729 (42.5%) | 198 (12.3%) | 295 (13.7%) | 1,065 (15.7%) |
| 60-69 | 64,276 (34.7%) | 9,599 (35.9%) | 5,966 (40.3%) | 2,745 (36.6%) | 472 (39.1%) | 332 (36.8%) | 797 (46.4%) | 556 (34.6%) | 772 (35.8%) | 2,583 (38.0%) |
| 70-79 | 81,121 (43.8%) | 10,850 (40.6%) | 5,178 (35.0%) | 3,204 (42.7%) | 428 (35.5%) | 383 (42.4%) | 177 (10.3%) | 684 (42.5%) | 925 (42.9%) | 2,681 (39.4%) |
| 80+ | 16,094 (8.7%) | 2,002 (7.5%) | 771 (5.2%) | 586 (7.8%) | 66 (5.5%) | 70 (7.8%) | 14 (0.8%) | 171 (10.6%) | 165 (7.6%) | 476 (7.0%) |
| **Sex** | | | | | | | | | | |
| Female | 107,071 (57.9%) | 18,342 (68.6%) | 8,513 (57.5%) | 5,456 (72.7%) | 765 (63.4%) | 502 (55.6%) | 968 (56.4%) | 820 (51.0%) | 604 (28.0%) | 3,721 (54.7%) |
| Male | 77,985 (42.1%) | 8,387 (31.4%) | 6,283 (42.5%) | 2,050 (27.3%) | 442 (36.6%) | 401 (44.4%) | 749 (43.6%) | 789 (49.0%) | 1,553 (72.0%) | 3,084 (45.3%) |
| **Ethnicity** | | | | | | | | | | |
| Asian or Asian British | 2,063 (1.1%) | 304 (1.1%) | 156 (1.1%) | 36 (0.5%) | 20 (1.7%) | 30 (3.3%) | 31 (1.8%) | 35 (2.2%) | 18 (0.8%) | 71 (1.0%) |
| Black or Black British | 1,294 (0.7%) | 226 (0.8%) | 95 (0.6%) | 19 (0.3%) | 12 (1.0%) | 17 (1.9%) | 41 (2.4%) | 18 (1.1%) | 10 (0.5%) | 50 (0.7%) |
| Mixed Race or Other | 1,996 (1.1%) | 368 (1.4%) | 215 (1.5%) | 47 (0.6%) | 23 (1.9%) | 12 (1.3%) | 47 (2.7%) | 21 (1.3%) | 23 (1.1%) | 86 (1.3%) |
| White | 179,047 (96.8%) | 25,727 (96.3%) | 14,270 (96.4%) | 7,380 (98.3%) | 1,148 (95.1%) | 843 (93.4%) | 1,588 (92.5%) | 1,531 (95.2%) | 2,101 (97.4%) | 6,558 (96.4%) |
| Missing | 656 (0.4%) | 104 (0.4%) | 60 (0.4%) | 24 (0.3%) | 4 (0.3%) | 1 (0.1%) | 10 (0.6%) | 4 (0.2%) | 5 (0.2%) | 40 (0.6%) |
| **Highest qualification** | | | | | | | | | | |

| | | | | | | | | | | |
|---|---|---|---|---|---|---|---|---|---|---|
| College or University degree | 81,005 (43.8%) | 10,907 (40.8%) | 5,779 (39.1%) | 2,779 (37.0%) | 513 (42.5%) | 347 (38.4%) | 661 (38.5%) | 679 (42.2%) | 808 (37.5%) | 3,143 (46.2%) |
| Vocational qualifications | 50,578 (27.3%) | 7,592 (28.4%) | 4,437 (30.0%) | 2,251 (30.0%) | 351 (29.1%) | 244 (27.0%) | 577 (33.6%) | 443 (27.5%) | 689 (31.9%) | 1,796 (26.4%) |
| National exams at ages 16-18 | 38,692 (20.9%) | 5,975 (22.4%) | 3,378 (22.8%) | 1,800 (24.0%) | 251 (20.8%) | 193 (21.4%) | 400 (23.3%) | 297 (18.5%) | 459 (21.3%) | 1,387 (20.4%) |
| None of the above | 12,886 (7.0%) | 1,954 (7.3%) | 1,042 (7.0%) | 606 (8.1%) | 79 (6.5%) | 106 (11.7%) | 59 (3.4%) | 173 (10.8%) | 182 (8.4%) | 406 (6.0%) |
| Missing | 1,895 (1.0%) | 301 (1.1%) | 160 (1.1%) | 70 (0.9%) | 13 (1.1%) | 13 (1.4%) | 20 (1.2%) | 17 (1.1%) | 19 (0.9%) | 73 (1.1%) |
| **Townsend deprivation index** | | | | | | | | | | |
| Least Deprived | 76,751 (41.5%) | 10,273 (38.4%) | 5,466 (36.9%) | 3,149 (42.0%) | 379 (31.4%) | 327 (36.2%) | 498 (29.0%) | 571 (35.5%) | 912 (42.3%) | 2,537 (37.3%) |
| 2nd Quintile | 39,309 (21.2%) | 5,324 (19.9%) | 3,027 (20.5%) | 1,565 (20.8%) | 205 (17.0%) | 212 (23.5%) | 312 (18.2%) | 339 (21.1%) | 466 (21.6%) | 1,316 (19.3%) |
| 3rd Quintile | 27,699 (15.0%) | 4,100 (15.3%) | 2,315 (15.6%) | 1,087 (14.5%) | 195 (16.2%) | 146 (16.2%) | 290 (16.9%) | 246 (15.3%) | 304 (14.1%) | 1,050 (15.4%) |
| 4th Quintile | 22,441 (12.1%) | 3,580 (13.4%) | 2,043 (13.8%) | 917 (12.2%) | 216 (17.9%) | 117 (13.0%) | 280 (16.3%) | 234 (14.5%) | 221 (10.2%) | 917 (13.5%) |
| Most Deprived | 18,637 (10.1%) | 3,399 (12.7%) | 1,914 (12.9%) | 781 (10.4%) | 209 (17.3%) | 99 (11.0%) | 334 (19.5%) | 212 (13.2%) | 247 (11.5%) | 969 (14.2%) |
| Missing | 219 (0.1%) | 53 (0.2%) | 31 (0.2%) | 7 (0.1%) | 3 (0.2%) | 2 (0.2%) | 3 (0.2%) | 7 (0.4%) | 7 (0.3%) | 16 (0.2%) |
| **Employment status** | | | | | | | | | | |
| Employed | 123,177 (66.6%) | 17,273 (64.6%) | 10,351 (70.0%) | 4,939 (65.8%) | 811 (67.2%) | 631 (69.9%) | 1,527 (88.9%) | 1,024 (63.6%) | 1,425 (66.1%) | 4,461 (65.6%) |
| Not employed | 61,453 (33.2%) | 9,380 (35.1%) | 4,403 (29.8%) | 2,553 (34.0%) | 391 (32.4%) | 270 (29.9%) | 190 (11.1%) | 581 (36.1%) | 727 (33.7%) | 2,318 (34.1%) |
| Missing | 426 (0.2%) | 76 (0.3%) | 42 (0.3%) | 14 (0.2%) | 5 (0.4%) | 2 (0.2%) | 0 (0.0%) | 4 (0.2%) | 5 (0.2%) | 26 (0.4%) |
| **Body mass index** | | | | | | | | | | |
| Underweight | 1,005 (0.5%) | 153 (0.6%) | 49 (0.3%) | 34 (0.5%) | 3 (0.2%) | 3 (0.3%) | 2 (0.1%) | 4 (0.2%) | 2 (0.1%) | 45 (0.7%) |

| | | | | | | | | | | |
|---|---|---|---|---|---|---|---|---|---|---|
| Normal weight | 70,668 (38.2%) | 9,505 (35.6%) | 3,811 (25.8%) | 2,772 (36.9%) | 327 (27.1%) | 316 (35.0%) | 547 (31.9%) | 525 (32.6%) | 614 (28.5%) | 2,285 (33.6%) |
| Overweight | 76,487 (41.3%) | 10,389 (38.9%) | 6,178 (41.8%) | 2,971 (39.6%) | 437 (36.2%) | 391 (43.3%) | 651 (37.9%) | 677 (42.1%) | 1,010 (46.8%) | 2,776 (40.8%) |
| Obese | 36,454 (19.7%) | 6,609 (24.7%) | 4,718 (31.9%) | 1,713 (22.8%) | 431 (35.7%) | 193 (21.4%) | 512 (29.8%) | 398 (24.7%) | 528 (24.5%) | 1,681 (24.7%) |
| Missing | 442 (0.2%) | 73 (0.3%) | 40 (0.3%) | 16 (0.2%) | 9 (0.7%) | 0 (0.0%) | 5 (0.3%) | 5 (0.3%) | 3 (0.1%) | 18 (0.3%) |
| **Smoking status** | | | | | | | | | | |
| Never | 108,409 (58.6%) | 15,082 (56.4%) | 7,910 (53.5%) | 4,169 (55.5%) | 619 (51.3%) | 520 (57.6%) | 957 (55.7%) | 902 (56.1%) | 1,120 (51.9%) | 3,716 (54.6%) |
| Previous | 63,168 (34.1%) | 9,306 (34.8%) | 5,314 (35.9%) | 2,746 (36.6%) | 393 (32.6%) | 316 (35.0%) | 515 (30.0%) | 576 (35.8%) | 832 (38.6%) | 2,418 (35.5%) |
| Current | 13,017 (7.0%) | 2,264 (8.5%) | 1,527 (10.3%) | 577 (7.7%) | 191 (15.8%) | 62 (6.9%) | 240 (14.0%) | 126 (7.8%) | 197 (9.1%) | 652 (9.6%) |
| Missing | 462 (0.2%) | 77 (0.3%) | 45 (0.3%) | 14 (0.2%) | 4 (0.3%) | 5 (0.6%) | 5 (0.3%) | 5 (0.3%) | 8 (0.4%) | 19 (0.3%) |
| **Alcohol consumption frequency** | | | | | | | | | | |
| <1 times/week | 47,328 (25.6%) | 8,291 (31.0%) | 4,680 (31.6%) | 2,309 (30.8%) | 466 (38.6%) | 300 (33.2%) | 638 (37.2%) | 464 (28.8%) | 475 (22.0%) | 1,925 (28.3%) |
| 1-2 times/week | 47,529 (25.7%) | 6,834 (25.6%) | 3,800 (25.7%) | 1,975 (26.3%) | 275 (22.8%) | 219 (24.3%) | 450 (26.2%) | 411 (25.5%) | 528 (24.5%) | 1,516 (22.3%) |
| 3-4 times/week | 48,711 (26.3%) | 6,294 (23.5%) | 3,377 (22.8%) | 1,756 (23.4%) | 242 (20.0%) | 207 (22.9%) | 361 (21.0%) | 388 (24.1%) | 606 (28.1%) | 1,613 (23.7%) |
| Daily | 41,334 (22.3%) | 5,288 (19.8%) | 2,924 (19.8%) | 1,459 (19.4%) | 224 (18.6%) | 176 (19.5%) | 267 (15.6%) | 343 (21.3%) | 546 (25.3%) | 1,742 (25.6%) |
| Missing | 154 (0.1%) | 22 (0.1%) | 15 (0.1%) | 7 (0.1%) | 0 (0.0%) | 1 (0.1%) | 1 (0.1%) | 3 (0.2%) | 2 (0.1%) | 9 (0.1%) |
| **Self-reported overall health rating** | | | | | | | | | | |
| Excellent | 40,056 (21.6%) | 3,221 (12.1%) | 1,331 (9.0%) | 1,191 (15.9%) | 135 (11.2%) | 203 (22.5%) | 182 (10.6%) | 266 (16.5%) | 358 (16.6%) | 936 (13.8%) |

| | | | | | | | | | | |
|---|---|---|---|---|---|---|---|---|---|---|
| Good | 111,924 (60.5%) | 14,766 (55.2%) | 7,923 (53.5%) | 4,428 (59.0%) | 595 (49.3%) | 536 (59.4%) | 900 (52.4%) | 935 (58.1%) | 1,170 (54.2%) | 3,656 (53.7%) |
| Fair | 28,549 (15.4%) | 7,000 (26.2%) | 4,364 (29.5%) | 1,561 (20.8%) | 368 (30.5%) | 139 (15.4%) | 527 (30.7%) | 326 (20.3%) | 486 (22.5%) | 1,718 (25.2%) |
| Poor | 4,092 (2.2%) | 1,644 (6.2%) | 1,124 (7.6%) | 309 (4.1%) | 108 (8.9%) | 22 (2.4%) | 105 (6.1%) | 74 (4.6%) | 140 (6.5%) | 471 (6.9%) |
| Missing | 435 (0.2%) | 98 (0.4%) | 54 (0.4%) | 17 (0.2%) | 1 (0.1%) | 3 (0.3%) | 3 (0.2%) | 8 (0.5%) | 3 (0.1%) | 24 (0.4%) |
| **Self-reported prescribed sleep medication** | | | | | | | | | | |
| Yes | 1,167 (0.6%) | 436 (1.6%) | 212 (1.4%) | 83 (1.1%) | 19 (1.6%) | 1 (0.1%) | 15 (0.9%) | 19 (1.2%) | 22 (1.0%) | 125 (1.8%) |
| No | 183,814 (99.3%) | 26,282 (98.3%) | 14,580 (98.5%) | 7,422 (98.9%) | 1,188 (98.4%) | 902 (99.9%) | 1,701 (99.1%) | 1,587 (98.6%) | 2,135 (99.0%) | 6,676 (98.1%) |
| Missing | 75 (0.0%) | 11 (0.0%) | 4 (0.0%) | 1 (0.0%) | 0 (0.0%) | 0 (0.0%) | 1 (0.1%) | 3 (0.2%) | 0 (0.0%) | 4 (0.1%) |

**eTable 5.** Association between baseline sleep difficulties (2006-2010) and operationally defined sleep disorders at follow-up (2023)

| | Overall (n = 185,056) | Chronic Insomnia Disorder (n = 26,729) | Obstructive Sleep Apnoea (n = 14,796) | Restless Legs Syndrome (n = 7,506) | Delayed Sleep-Wake Phase Disorder (n = 1,207) | Advanced Sleep-Wake Phase Disorder (n = 903) | Shift Work Disorder (n = 1,717) | Sleepwalking (n = 1,609) | REM Sleep Behaviour Disorder (n = 2,157) | Nightmare Disorder (n = 6,805) |
|---|---|---|---|---|---|---|---|---|---|---|
| **Sleep duration** | | | | | | | | | | |
| <7 hours | 41,291 (22.3%) | 9,685 (36.2%) | 4,461 (30.2%) | 2,155 (28.7%) | 385 (31.9%) | 218 (24.1%) | 661 (38.5%) | 406 (25.2%) | 520 (24.1%) | 1,837 (27.0%) |
| 7-8.9 hours | 132,297 (71.5%) | 15,452 (57.8%) | 9,128 (61.7%) | 4,876 (65.0%) | 700 (58.0%) | 604 (66.9%) | 969 (56.4%) | 1,092 (67.9%) | 1,469 (68.1%) | 4,383 (64.4%) |
| 9+ hours | 10,969 (5.9%) | 1,446 (5.4%) | 1,142 (7.7%) | 442 (5.9%) | 117 (9.7%) | 80 (8.9%) | 80 (4.7%) | 102 (6.3%) | 164 (7.6%) | 562 (8.3%) |
| Missing | 499 (0.3%) | 146 (0.5%) | 65 (0.4%) | 33 (0.4%) | 5 (0.4%) | 1 (0.1%) | 7 (0.4%) | 9 (0.6%) | 4 (0.2%) | 23 (0.3%) |
| **Chronotype** | | | | | | | | | | |
| Definitely morning type | 41,748 (22.6%) | 6,028 (22.6%) | 2,689 (18.2%) | 1,672 (22.3%) | 23 (1.9%) | 565 (62.6%) | 323 (18.8%) | 389 (24.2%) | 478 (22.2%) | 1,535 (22.6%) |
| Intermediate type | 107,874 (58.3%) | 15,461 (57.8%) | 8,772 (59.3%) | 4,401 (58.6%) | 450 (37.3%) | 300 (33.2%) | 955 (55.6%) | 908 (56.4%) | 1,219 (56.5%) | 3,765 (55.3%) |
| Definitely evening type | 15,408 (8.3%) | 2,713 (10.2%) | 1,917 (13.0%) | 743 (9.9%) | 691 (57.2%) | 4 (0.4%) | 260 (15.1%) | 138 (8.6%) | 218 (10.1%) | 807 (11.9%) |
| Missing | 20,026 (10.8%) | 2,527 (9.5%) | 1,418 (9.6%) | 690 (9.2%) | 43 (3.6%) | 34 (3.8%) | 179 (10.4%) | 174 (10.8%) | 242 (11.2%) | 698 (10.3%) |
| **Insomnia** | | | | | | | | | | |
| With frequent insomnia | 49,265 (26.6%) | 13,495 (50.5%) | 6,221 (42.0%) | 3,149 (42.0%) | 395 (32.7%) | 269 (29.8%) | 649 (37.8%) | 531 (33.0%) | 736 (34.1%) | 2,923 (43.0%) |
| Without frequent insomnia | 135,602 (73.3%) | 13,212 (49.4%) | 8,563 (57.9%) | 4,349 (57.9%) | 811 (67.2%) | 632 (70.0%) | 1,067 (62.1%) | 1,074 (66.7%) | 1,419 (65.8%) | 3,875 (56.9%) |
| Missing | 189 (0.1%) | 22 (0.1%) | 12 (0.1%) | 8 (0.1%) | 1 (0.1%) | 2 (0.2%) | 1 (0.1%) | 4 (0.2%) | 2 (0.1%) | 7 (0.1%) |
| **Daytime sleepiness** | | | | | | | | | | |

| | | | | | | | | | | |
|---|---|---|---|---|---|---|---|---|---|---|
| With frequent sleepiness | 3,950 (2.1%) | 1,015 (3.8%) | 759 (5.1%) | 251 (3.3%) | 63 (5.2%) | 38 (4.2%) | 59 (3.4%) | 43 (2.7%) | 100 (4.6%) | 280 (4.1%) |
| Without frequent sleepiness | 180,747 (97.7%) | 25,648 (96.0%) | 13,995 (94.6%) | 7,241 (96.5%) | 1,141 (94.5%) | 861 (95.3%) | 1,656 (96.4%) | 1,558 (96.8%) | 2,052 (95.1%) | 6,505 (95.6%) |
| Missing | 359 (0.2%) | 66 (0.2%) | 42 (0.3%) | 14 (0.2%) | 3 (0.2%) | 4 (0.4%) | 2 (0.1%) | 8 (0.5%) | 5 (0.2%) | 20 (0.3%) |
| **Shift work** | | | | | | | | | | |
| Day work | 106,086 (57.3%) | 14,412 (53.9%) | 8,572 (57.9%) | 4,145 (55.2%) | 640 (53.0%) | 500 (55.4%) | 873 (50.8%) | 852 (53.0%) | 1,153 (53.5%) | 3,699 (54.4%) |
| Shift work without night shifts | 8,655 (4.7%) | 1,436 (5.4%) | 854 (5.8%) | 407 (5.4%) | 89 (7.4%) | 65 (7.2%) | 223 (13.0%) | 81 (5.0%) | 129 (6.0%) | 377 (5.5%) |
| Shift work with night shifts | 6,178 (3.3%) | 1,043 (3.9%) | 698 (4.7%) | 300 (4.0%) | 62 (5.1%) | 48 (5.3%) | 304 (17.7%) | 66 (4.1%) | 105 (4.9%) | 290 (4.3%) |
| Shift work with unknown schedule | 11 (0.0%) | 2 (0.0%) | 2 (0.0%) | 0 (0.0%) | 0 (0.0%) | 0 (0.0%) | 1 (0.1%) | 0 (0.0%) | 0 (0.0%) | 3 (0.0%) |
| Permanent night shift work | 2,153 (1.2%) | 355 (1.3%) | 218 (1.5%) | 86 (1.1%) | 20 (1.7%) | 18 (2.0%) | 124 (7.2%) | 22 (1.4%) | 31 (1.4%) | 83 (1.2%) |
| Missing | 61,973 (33.5%) | 9,481 (35.5%) | 4,452 (30.1%) | 2,568 (34.2%) | 396 (32.8%) | 272 (30.1%) | 192 (11.2%) | 588 (36.5%) | 739 (34.3%) | 2,353 (34.6%) |
| **Difficulty getting up in the morning** | | | | | | | | | | |
| Very easy | 56,316 (30.4%) | 5,785 (21.6%) | 2,557 (17.3%) | 1,868 (24.9%) | 59 (4.9%) | 490 (54.3%) | 331 (19.3%) | 533 (33.1%) | 640 (29.7%) | 1,514 (22.2%) |
| Fairly easy | 94,567 (51.1%) | 12,980 (48.6%) | 6,896 (46.6%) | 3,741 (49.8%) | 353 (29.2%) | 356 (39.4%) | 773 (45.0%) | 750 (46.6%) | 1,021 (47.3%) | 3,221 (47.3%) |
| Not very easy | 26,423 (14.3%) | 5,729 (21.4%) | 3,767 (25.5%) | 1,405 (18.7%) | 476 (39.4%) | 44 (4.9%) | 442 (25.7%) | 242 (15.0%) | 339 (15.7%) | 1,427 (21.0%) |
| Not at all easy | 6,357 (3.4%) | 2,017 (7.5%) | 1,455 (9.8%) | 440 (5.9%) | 308 (25.5%) | 5 (0.6%) | 154 (9.0%) | 74 (4.6%) | 142 (6.6%) | 593 (8.7%) |

| | | | | | | | | | | |
|---|---|---|---|---|---|---|---|---|---|---|
| Missing | 1,393 (0.8%) | 218 (0.8%) | 121 (0.8%) | 52 (0.7%) | 11 (0.9%) | 8 (0.9%) | 17 (1.0%) | 10 (0.6%) | 15 (0.7%) | 50 (0.7%) |
| **Snoring** | | | | | | | | | | |
| Yes | 61,998 (33.5%) | 8,798 (32.9%) | 8,418 (56.9%) | 2,462 (32.8%) | 457 (37.9%) | 327 (36.2%) | 572 (33.3%) | 554 (34.4%) | 1,048 (48.6%) | 2,516 (37.0%) |
| No | 111,407 (60.2%) | 15,946 (59.7%) | 5,496 (37.1%) | 4,594 (61.2%) | 645 (53.4%) | 514 (56.9%) | 991 (57.7%) | 941 (58.5%) | 1,035 (48.0%) | 3,810 (56.0%) |
| Missing | 11,651 (6.3%) | 1,985 (7.4%) | 882 (6.0%) | 450 (6.0%) | 105 (8.7%) | 62 (6.9%) | 154 (9.0%) | 114 (7.1%) | 74 (3.4%) | 479 (7.0%) |

**eTable 6.** Potential correlates of sleep by operationally defined sleep disorders

| | **Overall** (n = 185,056) | **Chronic Insomnia Disorder** (n = 26,729) | **Obstructive Sleep Apnoea** (n = 14,796) | **Restless Legs Syndrome** (n = 7,506) | **Delayed Sleep-Wake Phase Disorder** (n = 1,207) | **Advanced Sleep-Wake Phase Disorder** (n = 903) | **Shift Work Disorder** (n = 1,717) | **Sleepwalking** (n = 1,609) | **REM Sleep Behaviour Disorder** (n = 2,157) | **Nightmare Disorder** (n = 6,805) |
|---|---|---|---|---|---|---|---|---|---|---|
| **Fatigue (FFS, 0-31. High = more fatigue)** | | | | | | | | | | |
| Mean (SD) | 5.8 (6.3) | 13.1 (6.4) | 13.9 (6.2) | 8.9 (7.0) | 11.1 (6.9) | 6.3 (6.0) | 14.0 (6.3) | 7.5 (7.0) | 8.8 (7.7) | 10.8 (7.4) |
| **Cognitive impairment (BC-CCI, 0-18. High = more cognitive impairments)** | | | | | | | | | | |
| Mean (SD) | 3.8 (3.2) | 6.3 (3.8) | 6.5 (3.9) | 5.0 (3.5) | 5.7 (3.9) | 4.5 (3.2) | 7.0 (4.1) | 4.7 (3.8) | 5.8 (4.1) | 5.8 (4.0) |
| **Depression (PHQ-2, 0-6. High = more depressed)** | | | | | | | | | | |
| Mean (SD) | 0.7 (1.2) | 1.8 (1.7) | 1.8 (1.8) | 1.1 (1.5) | 1.6 (1.8) | 0.8 (1.2) | 2.3 (1.8) | 1.0 (1.5) | 1.4 (1.7) | 1.8 (1.9) |
| Above threshold (≥ 3) | 12,437 (6.7%) | 6,331 (23.7%) | 3,641 (24.6%) | 876 (11.7%) | 264 (21.9%) | 64 (7.1%) | 536 (31.2%) | 191 (11.9%) | 405 (18.8%) | 1,696 (24.9%) |
| Below threshold (<3) | 166,102 (89.8%) | 19,933 (74.6%) | 10,983 (74.2%) | 6,599 (87.9%) | 923 (76.5%) | 827 (91.6%) | 1,069 (62.3%) | 1,392 (86.5%) | 1,731 (80.3%) | 5,024 (73.8%) |
| Missing | 6,517 (3.5%) | 465 (1.7%) | 172 (1.2%) | 31 (0.4%) | 20 (1.7%) | 12 (1.3%) | 112 (6.5%) | 26 (1.6%) | 21 (1.0%) | 85 (1.2%) |
| **Anxiety (GAD-2, 0-6. High = more anxious)** | | | | | | | | | | |
| Mean (SD) | 1.0 (1.4) | 2.2 (1.9) | 2.0 (1.9) | 1.4 (1.7) | 1.6 (1.8) | 1.2 (1.6) | 2.5 (1.9) | 1.3 (1.7) | 1.7 (1.8) | 2.3 (2.0) |
| Above threshold (≥ 3) | 18,271 (9.9%) | 8,216 (30.7%) | 3,973 (26.9%) | 1,211 (16.1%) | 242 (20.0%) | 122 (13.5%) | 585 (34.1%) | 270 (16.8%) | 465 (21.6%) | 2,230 (32.8%) |
| Below threshold (<3) | 160,176 (86.6%) | 18,060 (67.6%) | 10,649 (72.0%) | 6,269 (83.5%) | 947 (78.5%) | 768 (85.0%) | 1,018 (59.3%) | 1,310 (81.4%) | 1,675 (77.7%) | 4,495 (66.1%) |
| Missing | 6,609 (3.6%) | 453 (1.7%) | 174 (1.2%) | 26 (0.3%) | 18 (1.5%) | 13 (1.4%) | 114 (6.6%) | 29 (1.8%) | 17 (0.8%) | 80 (1.2%) |
| **Accidents or near-miss due to sleepiness in the past year** | | | | | | | | | | |
| Never | 172,653 (93.3%) | 24,229 (90.6%) | 13,419 (90.7%) | 7,057 (94.0%) | 1,107 (91.7%) | 849 (94.0%) | 1,394 (81.2%) | 1,511 (93.9%) | 1,947 (90.3%) | 6,186 (90.9%) |
| Once | 3,679 (2.0%) | 850 (3.2%) | 525 (3.5%) | 203 (2.7%) | 52 (4.3%) | 26 (2.9%) | 108 (6.3%) | 37 (2.3%) | 81 (3.8%) | 220 (3.2%) |

| | | | | | | | | | | |
|---|---|---|---|---|---|---|---|---|---|---|
| 2-5 times | 1,114 (0.6%) | 520 (1.9%) | 295 (2.0%) | 98 (1.3%) | 13 (1.1%) | 17 (1.9%) | 44 (2.6%) | 19 (1.2%) | 53 (2.5%) | 131 (1.9%) |
| 5-10 times | 137 (0.1%) | 78 (0.3%) | 57 (0.4%) | 14 (0.2%) | 4 (0.3%) | 0 (0.0%) | 16 (0.9%) | 5 (0.3%) | 11 (0.5%) | 26 (0.4%) |
| More than 10 times | 91 (0.0%) | 53 (0.2%) | 37 (0.3%) | 11 (0.1%) | 3 (0.2%) | 0 (0.0%) | 3 (0.2%) | 3 (0.2%) | 5 (0.2%) | 20 (0.3%) |
| Missing | 7,382 (4.0%) | 999 (3.7%) | 463 (3.1%) | 123 (1.6%) | 28 (2.3%) | 11 (1.2%) | 152 (8.9%) | 34 (2.1%) | 60 (2.8%) | 222 (3.3%) |

**Abbreviation:** FFS, Flinders Fatigue Scale; BC-CCI, British Columbia Cognitive Complaints Inventory; PHQ-4 (PHQ-2 and GAD-2), Patient Health Questionnaire 4-item (Patient Health Questionnaire 2-item, Generalized Anxiety Disorder 2-item)

**eTable 7.** Family history of sleep disorders by operationally defined sleep disorders

| | Those who reported positive family history, n (%) |
|---|---|
| **Chronic Insomnia Disorder** | |
| Case (n = 26,729) | 8,432 (31.5%) |
| Non-case (n = 153,762) | 28,925 (18.8%) |
| **Obstructive sleep apnoea** | |
| Case (n = 14,796) | 1,991 (13.5%) |
| Non-case (n = 165,379) | 11,391 (6.9%) |
| **Sleepwalking** | |
| Case (n = 1,609) | 152 (9.4%) |
| Non-case (n = 176,300) | 11,792 (6.7%) |
| **Nightmare Disorder** | |
| Case (n = 6,805) | 812 (11.9%) |
| Non-case (n = 168,993) | 9,195 (5.4%) |
| **Restless legs syndrome** | |
| Case (n = 7,506) | 2,558 (34.1%) |
| Non-case (n = 168,127) | 18,871 (11.2%) |
